# Polysomnographic parameters in schizoaffective disorder: a systematic review and meta-analysis

**DOI:** 10.64898/2026.04.06.26350239

**Authors:** Dario Morra, Gianluca Ficca, Giuseppe Barbato

## Abstract

A systematic review and meta-analysis of sleep studies in schizoaffective disorder were conducted using published articles researched in major databases within the period from inception to December 1, 2025. The sleep parameters: total sleep time, sleep efficiency, sleep latency, wakefulness, REM time and percentage, REM latency, REM density, stage 1, 2, 3 and 4 sleep time and percentage, delta sleep time and percentage, of drug-free schizoaffective patients were analyzed and, where available, compared with case-control data of healthy controls, depressed unipolar patients and schizophrenic patients. Forty studies were identified in the systematic review. Nine case-control studies with 67 schizoaffective patients, 88 schizophrenic patients, 79 healthy controls and 131 depressed patients were included in the meta-analyses. The primary outcome was the standard mean difference. Data were fitted with a random-effects model. Publication bias assessment was checked by Egger’s Regression and funnel plot asymmetry. Patients with schizoaffective disorder showed reduced total sleep time, increased sleep latency and wakefulness, along with reduced REM time and shortened REM latency, reduced stage 4 sleep time and percentage compared to healthy controls. Patients with schizoaffective disorder differed from depressed patients only for increased sleep latency, while they did not show any difference compared to patients with schizophrenia. SZA showed a non-significant trend (p=0.08) towards increased REM density compared to SCZ, suggesting the need to better clarify the role of REM density in mood and psychotic disorders.

## 1. Introduction

### 1.1 Schizoaffective disorder

Schizoaffective disorder (SZA) is a chronic condition which, according to current DSM-V criteria, can be diagnosed when a mood disorder (depression, mania or both) is present for the majority of the total active and residual course of schizophrenia (SCZ), from the onset of psychotic symptoms up until the current diagnosis (Jäger et al., 2011; Malaspina et al., 2013; Peterson et al., 2019). SZA is estimated to have a lifetime prevalence of 0.3%-1.1% (Sovner & McHugh, 1976; Rosenthal et al., 1980; Jäger et al., 2011). Age of onset of SZA is 25-35, intermediate between that of SCZ and mood disorders (Cheniaux et al., 2008; Rink et al., 2016; Miller & Black, 2019), although it might be broader (Abrams et al., 2008). Women are twice as likely to be diagnosed with SZA than men, as for mood disorders (Angst et al., 1980; Marneros et al., 1990; Scully et al., 2004; Malhi et al., 2008; Rink et al., 2016; Miller & Black, 2019). Mixed states in SZA have been reported to be as frequent as mixed states in BD and probably linked to a worse outcome (Marneros et al., 2004). Lifetime suicide risk in SZA is estimated to be 5%, although the presence of depressive symptoms might increase it (Hor & Taylor, 2010; Miller et al., 2019). SZA is mostly regarded as having a more favourable course than that of patients with SCZ but worse than that of patients with BD (Harrow et al., 2000; Marneros et al., 2004; Pinna et al., 2014; Miller & Black, 2019). The historical perspective proposed in 1920 by Kraepelin suggested that SCZ and BD were the two main diagnostic categories, with the psychoses associated with the former category having a poorer outcome than those associated with mood disorders (Angst et al., 2002). The dichotomy proposed by Kraepelin, however did not acknowledge those cases in which patients experience psychotic symptoms but also have periods of concomitant mood disorders. The term “schizoaffective psychosis” was first used by Jacob Kasanin in 1933 (Kasanin, 1933); therefore the “schizoaffective subtype” of “schizophrenic reaction” is present in DSM since its first 1952 edition (Malaspina et al., 2013). SZA started to be regarded as a separate diagnostic entity in DSM-III, while following editions added subtypes (depressed, bipolar, mixed) and eventually recognized SZA as a lifetime diagnosis starting from DSM-V (Malaspina et al., 2013). ICD-11 differs from DSM by dropping the longitudinal approach to the diagnosis of SZA, considering it difficult to ascertain the accurate temporal relationship between psychotic and mood episodes, proposing the coexistence yet independence of the two classes of symptoms as a main criterion (Peterson et al., 2019).

SZA has been generally regarded as having poor diagnostic reliability (Sovner & McHugh, 1976; Rosenthal et al., 1980; Cheniaux et al., 2008; Malhi et al., 2008; Jäger et al., 2011; Malaspina et al., 2013, Santelmann et al., 2015; Peterson et al., 2019), making SZA a potentially misdiagnosed condition (Malaspina et al., 2013, Wilson et al., 2014; Paul et al., 2021; Wy & Saadabadi, 2025). The question of whether SZA is an actual separate clinical entity or a subtype of SCZ or an affective disorder has been frequently discussed in literature, contributing to the controversial nosography classification of SZA (Meltzer, 1984; Tsuang & Simpson, 1984; Lapierre, 1994; Jäger et al., 2004; Lake & Hurwitz, 2007; Cheniaux et al., 2008; Cheniaux et al., 2008; Kantrowitz & Citrome, 2011; Wilson et al., 2014; Santelmann et al., 2015; Rink et al., 2016; Miller & Black, 2019; Pavlichenko et al., 2024).

SZA is mainly treated with antipsychotics as for SCZ (Jäger et al., 2011; Kantrowitz & Citrome, 2011; Munoz-Negro et al., 2019; Wy & Saadabadi, 2025), with paliperidone being the only FDA approved medication for SZA (Canuso et al., 2010; Canuso et al., 2010). Patients with SZA, nonetheless, receive additional prescription of benzodiazepines, antidepressants and mood stabilizers (Jäger et al., 2011; Pinna et al., 2014; Wy & Saadabadi, 2025) and more frequently compared to patients with SCZ (Pinna et al., 2014).

Electroconvulsive therapy (ECT) is also effective in treating treatment-resistant SZA (Miller & Black, 2019; Wy & Saadabadi, 2025). Cognitive (Madre et al., 2016; Gotra et al., 2020; Dehelean et al., 2021) and gray matter abnormalities (Radonic et al., 2011, Ivleva et al., 2012; Ivleva et al., 2013, Amann et al., 2016) have been found in SZA, often resembling more those in SCZ than those in BD (Miller & Black, 2019; Gotra et al., 2020) or being intermediate between SCZ and BD (Lynham et al., 2022). In addition, recent fMRI (Hager et al., 2017), functional connectivity (Du et al., 2020) and immune system (Mazza et al., 2019) investigations have been conducted to find more specific markers of SZA. SZA patients do not show a different ventricle-to-brain ratio compared to SCZ or BD (Rieder et al., 1983; Meltzer, 1984). Heritability of SZA seems to be similar to that of SCZ and bipolar disorder (BD) (Cardno et al., 1999). Some studies found that SZA is genetically linked to both SCZ and BD and might constitute an intermediate form of these two conditions (Coryell & Zimmerman, 1988; Lapensee, 1992; Laursen et al., 2005), with evidence that genes involved in glutamatergic abnormalities might play a key role in the onset of SZA, SCZ and BD (Fallin et al., 2005). In a recent study on swedish population, SZA has been found to carry high genetic risk for both SCZ and BD while being clearly separable from psychotic BD, suggesting that SZA might not be, from a genetic perspective, a subtype of SZ, BD or MD (Kendler et al., 2025). As already suggested in early studies (Meltzer, 1984), SZA might be an intermediate condition in a putative polygenic continuum model of psychoses in which patients with major psychoses share genetic influences (Marneros, 2010; Guloksuz & van Os, 2018; Gama Marques & Ouakinin, 2021).

### 1.2 Sleep studies of schizoaffective disorder

Polysomnography (PSG) has been also used to assess both sleep architecture and sleep continuity alterations and understand their relationship with psychiatric conditions. Insomnia (Palmese et al., 2011; Kaskie et al., 2017; Laskemoen et al., 2019), obstructive sleep apnea (or at risk for it) (Alam et al., 2012; Annamalai et al., 2015; Kaskie et al., 2017) and circadian rhythm disorders (Poulin et al., 2010; Kaskie et al., 2017) are sleep disturbances reported in SZA, although these studies mostly mixed a minority of SZA patients in larger SCZ samples. Insomnia has been found to be associated with obesity (Palmese et al., 2011) and suicidal ideation and suicidal attempt risk in SZA patients (Miller et al., 2019), while having sleep disturbances was associated with more severe negative symptoms and lower functioning (Laskemoen et al., 2019). A recent study using a modern algorithm-based automatic sleep spindles detection method reported different sleep spindle phenotypes between SZA and SCZ; moreover, both slow and fast sleep spindles density were increased in those patients with a history of manic or hypomanic symptoms, such as those diagnosed with SZA or BD (Petit et al., 2022).

Sleep architecture has been investigated, although not extensively, in samples entirely made of SZA patients and compared these with healthy controls (HC), SCZ or affective disorders patients. The first study to date compared SZA to latent and acute SCZ patients: SZA showed increased sleep latency, decreased sleep efficiency and decreased delta sleep time compared to latent SCZ. SZA also showed shortened REM latency and increased REM density compared to both latent and acute SCZ. REM time in SZA was increased compared to acute SCZ (Reich et al., 1975). A subsequent study compared SZA to major depressive disorder (MDD) with psychotic features: SZA showed increased REM time and increased delta sleep time. SZA and psychotic MDD seemed to be very similar to each other especially for the reduced REM latency and the increased REM density, although SZA showed increased sleep latency while psychotic MDD showed longer wakefulness time (Kupfer & Foster, 1975). The lack of differences between SZA and psychotic MDD sleep was observed again in a subsequent study (Kupfer et al., 1979). SZA sleep latency was also found to be increased compared to MDD and SCZ (Zarcone & Benson, 1995). Compared to HC, SZA have shown increased sleep latency (Benson et al., 1980; Benson & Zarcone, 1985; Zarcone & Benson, 1995) and wakefulness (Benson et al., 1980), whereas total sleep time (Benson & Zarcone, 1985; Zarcone & Benson, 1995), Stage 4 sleep time and percentage (Benson et al., 1980; Benson & Zarcone, 1985), delta sleep time (Benson et al., 1980) and sleep efficiency (Zarcone & Benson, 1995) have been shown to be reduced. Also, SZA REM time was found to be decreased compared to that of HC (Benson & Zarcone, 1985). REM latency was not found to differ across SZA, SCZ and MDD (Zarcone, 1987) and between SZA and psychotic and non-psychotic MDD (Coble et al., 1981).

The present review and meta-analyses were addressed to further define sleep alteration in SZA patients and to clarify its degree of similarity with SCZ or MDD sleep. Presence of specific alteration could also characterize an SZA biological/neurophysiological background, that could help to differentiate SZA from overlapping diagnosis. Total sleep time (TST), sleep onset latency (SOL), sleep efficiency (SE), wake after sleep onset (WASO), REM time (REMT), REM percentage (%REM), REM latency (REML), REM density (REMD), stage 1, 2, 3 and 4 sleep time (ST1, ST2, ST3, ST4) and percentage (%ST1, %ST2, %ST3, %ST4), delta sleep time (DELTAT) and percentage (%DELTA) in drug-free SZA patients were analyzed and, where available, compared with case-control data of HC, MDD and SCZ patients.

## 2. Methods

The literature search was conducted querying PubMed, Web of Science (WoS) and PsycINFO databases from inception to 2025. The main keywords combination used to ensure the identification of all the relevant studies was: “(sleep or sleep*) AND schizo*”, although different other keywords were combined to perform a thorough research: “PSG or polysomnogra*”, “EEG OR electroencephalogra*”, “REM OR REMS OR rapid-eye-movement OR rapid-eye-movements”, “paradox OR paradox*”, “print OR print*”, “D-phase OR dream OR dream*”, “night OR night*”, “monit*”, “architect*”. Specific attention was devoted to identifying studies regarding the effect of drugs on schizoaffective patients, therefore further keywords also based on both molecule and brand name were implemented in the search, such as: “atypical”, “typical” and “antipsych*”, “paliperidone”, “risperidone”, “Invega”, “Risperdal” and so on. These queries returned 3345 studies from PubMed, 4105 studies from WoS and 65 studies from PsycINFO. Out of these 7515 studies, 4120 resulted in duplicates present on multiple databases. The remaining 3395 studies were entirely read to assess their usefulness: 3202 studies have eventually been excluded due to lack of sleep data provided. 13 studies have been excluded due to presenting duplicate samples already entirely presented in a previous study by the same author or co-author. 144 studies have been excluded due to lack of SZA patients. 36 studies were assessed as eligible and all of them were retrieved. 40 studies were included in the study: 36 studies were included from database research and 4 other eligible studies were manually researched and found. 9 studies were assessed as eligible for the meta-analysis, while the remaining 31 studies were assessed as eligible for the systematic review only. The eligibility decision was made after a full-text article reading by the authors (DM, GF and GB), both agreeing on the inclusion or exclusion of each study. The meta-analysis was conducted in order to investigate sleep parameters differences between the following groups: drug-free SZA and HC (Table 1), drug-free SZA and drug-free MDD (Table 2), drug-free SZA and drug-free SCZ (Table 3). Case-control studies included in the systematic review only are reported in Table 4 (with the relative reason for exclusion). All the other non-case-control studies included in the systematic review only are reported in Table 5. Figure 1 presents the PRISMA diagram of the systematic review.

**Figure 1.**
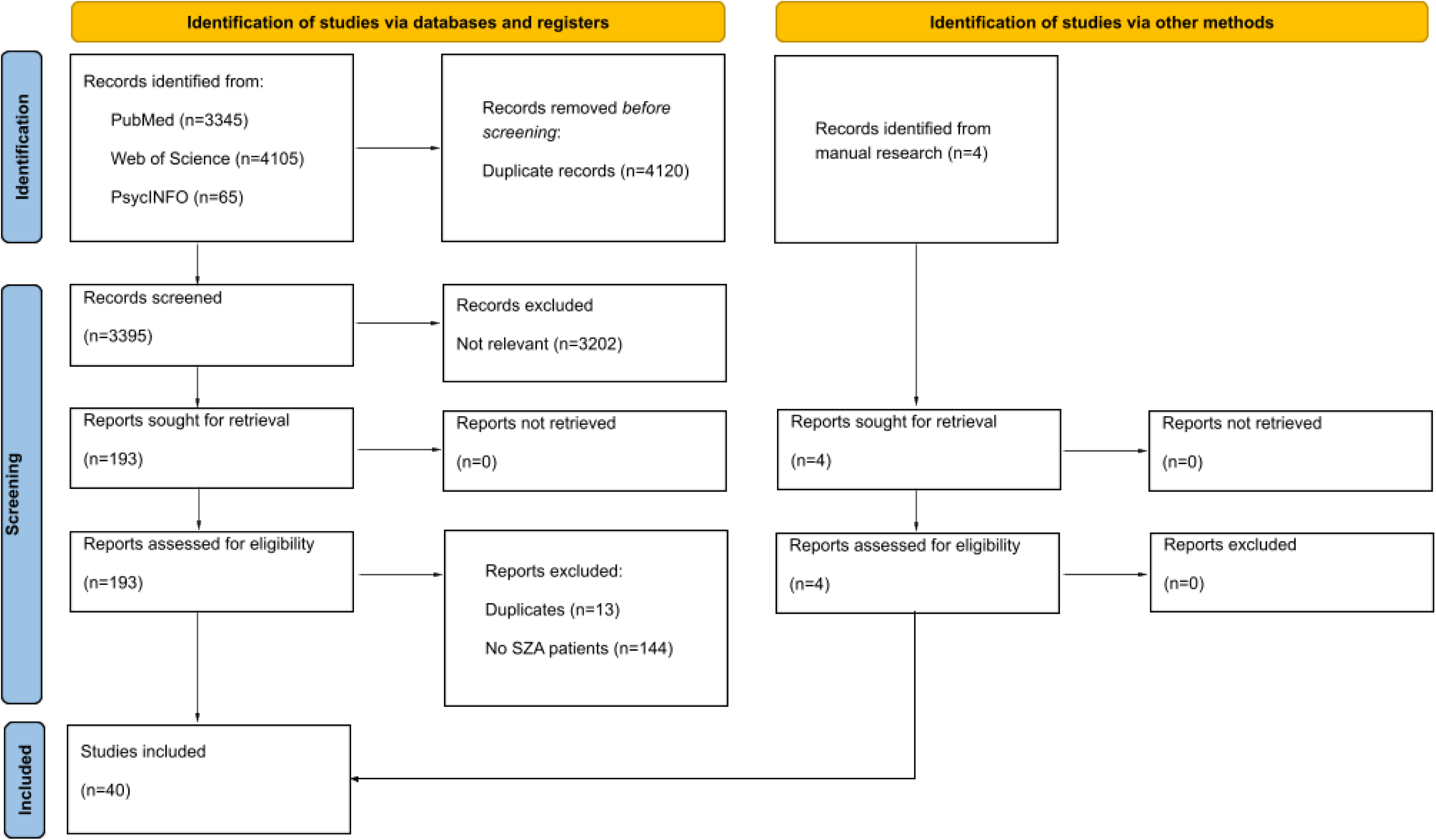
PRISMA diagram.

**Table 1.**
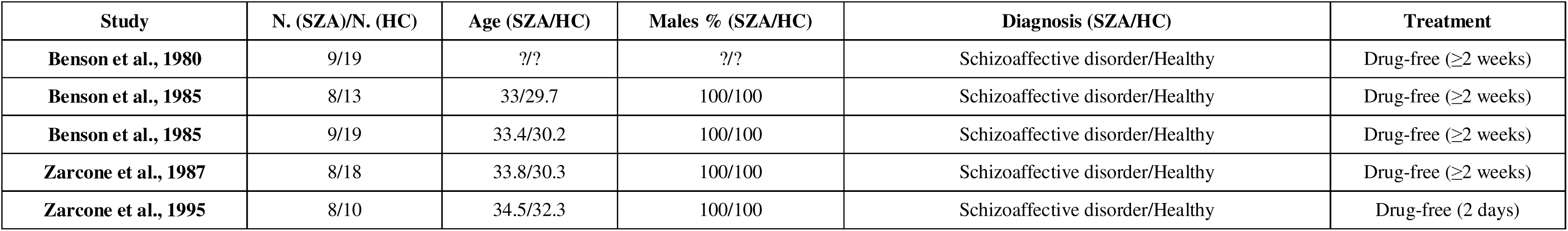
Drug-free SZA vs HC studies included in the meta-analysis.

**Table 2.**
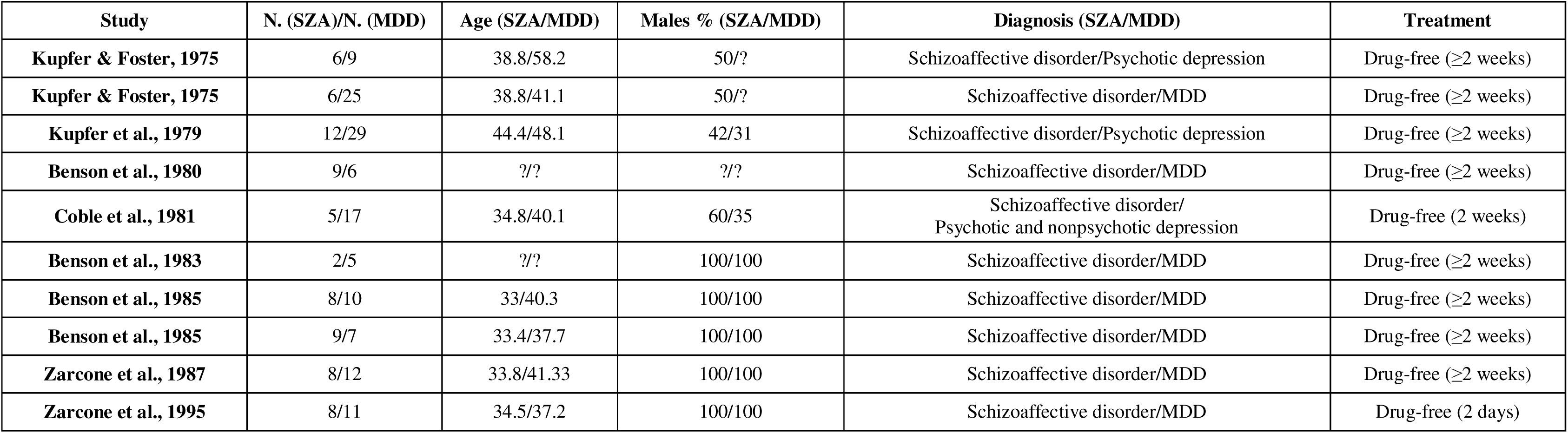
Drug-free SZA vs drug-free MDD studies included in the meta-analysis.

| Study | N. (SZA)/N. (MDD) | Age (SZA/MDD) | Males % (SZA/MDD) | Diagnosis (SZA/MDD) | Treatment |
| --- | --- | --- | --- | --- | --- |
| **Kupfer & Foster, 1975** | 6/9 | 38.8/58.2 | 50/? | Schizoaffective disorder/Psychotic depression | Drug-free ($\geq 2$ weeks) |
| **Kupfer & Foster, 1975** | 6/25 | 38.8/41.1 | 50/? | Schizoaffective disorder/MDD | Drug-free ($\geq 2$ weeks) |
| **Kupfer et al., 1979** | 12/29 | 44.4/48.1 | 42/31 | Schizoaffective disorder/Psychotic depression | Drug-free ($\geq 2$ weeks) |
| **Benson et al., 1980** | 9/6 | ?/? | ?/? | Schizoaffective disorder/MDD | Drug-free ($\geq 2$ weeks) |
| **Coble et al., 1981** | 5/17 | 34.8/40.1 | 60/35 | Schizoaffective disorder/<br>Psychotic and nonpsychotic depression | Drug-free (2 weeks) |
| **Benson et al., 1983** | 2/5 | ?/? | 100/100 | Schizoaffective disorder/MDD | Drug-free ($\geq 2$ weeks) |
| **Benson et al., 1985** | 8/10 | 33/40.3 | 100/100 | Schizoaffective disorder/MDD | Drug-free ($\geq 2$ weeks) |
| **Benson et al., 1985** | 9/7 | 33.4/37.7 | 100/100 | Schizoaffective disorder/MDD | Drug-free ($\geq 2$ weeks) |
| **Zarcone et al., 1987** | 8/12 | 33.8/41.33 | 100/100 | Schizoaffective disorder/MDD | Drug-free ($\geq 2$ weeks) |
| **Zarcone et al., 1995** | 8/11 | 34.5/37.2 | 100/100 | Schizoaffective disorder/MDD | Drug-free (2 days) |

**Table 3.** Drug-free SZA vs drug-free SCZ studies included in the meta-analysis.

| Study | N. (SZA)/N. (SCZ) | Age (SZA/SCZ) | Males % (SZA/SCZ) | Diagnosis (SZA/SCZ) | Treatment |
| --- | --- | --- | --- | --- | --- |
| **Reich et al., 1975** | 3/14 | 38.8/20.8 | 50/57 | Schizoaffective disorder/Schizophrenia (acute) | Drug-free ($\geq 2$ weeks) |
| **Reich et al., 1975** | 3/9 | 38.8/19 | 50/11 | Schizoaffective disorder/Schizophrenia (latent) | Drug-free ($\geq 2$ weeks) |
| **Benson et al., 1980** | 9/9 | ?/? | ?/? | Schizoaffective disorder/Schizophrenia | Drug-free ($\geq 2$ weeks) |
| **Benson et al., 1983** | 2/7 | ?/? | 100/100 | Schizoaffective disorder/Schizophrenia | Drug-free ($\geq 2$ weeks) |
| **Benson et al., 1985** | 8/11 | 33/27 | 100/100 | Schizoaffective disorder/Schizophrenia | Drug-free ($\geq 2$ weeks) |
| **Benson et al., 1985** | 9/8 | 33.4/26.6 | 100/100 | Schizoaffective disorder/Schizophrenia | Drug-free ($\geq 2$ weeks) |
| **Zarcone et al., 1987** | 8/12 | 33.8/26.2 | 100/100 | Schizoaffective disorder/Schizophrenia | Drug-free ($\geq 2$ weeks) |
| **Zarcone et al., 1995** | 8/18 | 34.5/33.1 | 100/100 | Schizoaffective disorder/Schizophrenia | Drug-free (2 days) |

**Table 4.** Case-control studies included in the systematic review only.

| **Study (excluded)** | **N. (SZA)/N. (CTRL)** | **Age (SZA/CTRL)** | **Males % (SZA/CTRL)** | **Diagnosis (SZA/CTRL)** | **Treatment** | **Reason of exclusion** |
| --- | --- | --- | --- | --- | --- | --- |
| **Kupfer et al., 1971** | 4 | 36.48 | 25 | 1 Schizophrenia<br>1 Schizoaffective disorder<br>2 Depressive neurosis | Drug-free ($\geq 2$ weeks) →<br>Chlorpromazine | Mixed sample |
| **Gillin et al., 1977** | 10 | 25 | 60 | 4 Schizoaffective disorder<br>2 Acute schizophrenia<br>1 Chronic schizophrenia<br>1 Schizoid personality disorder<br>2 Bipolar disorder | Drug-free ($\geq 2$ weeks) →<br>Pimozide | Mixed sample |
| **Gillin et al., 1978** | 7 | 23 | 43 | 3 Chronic schizophrenia<br>1 Acute schizophrenia<br>2 Schizoaffective disorder<br>1 Schizoid personality disorder + OCD | Drug-free ($\geq 1$ month) →<br>Pimozide | Mixed sample |
| **Keshavan et al., 1996** | 16 | 29.9 | 50 | 11 Schizophrenia<br>5 Schizoaffective disorder | Drug-free ($\geq 2$ weeks) →<br>Drug-free (31%)<br>Neuroleptics (69%)<br>+ Antidepressants<br>+ Mood stabilizers | Mixed medication<br>status/sample |
| **Keshavan et al., 1996** | 15 | 29.7 | ? | Schizophrenia<br>5 Schizoaffective disorder | Drug-free ($\geq 2$ weeks) →<br>Haloperidol (67%)<br>Fluphenazine (13%)<br>Trifluoperazine (6.7%) | Mixed sample |
|  |  |  |  |  | Clozapine (6.7%)<br>Risperidone (6.7%)<br>+ Anticholinergics (47%) |  |
| **Benson et al., 1996** | 14/15 | 33.6/30.5 | 100/100 | 11 Schizophrenia (presumably chronic)<br>3 Schizoaffective disorder/Healthy | Drug-free (2 days) | Mixed sample |
| **Nishino et al., 1998** | 14/14 | 33.3/34.4 | 100/100 | 12 Schizophrenia<br>2 Schizoaffective disorder/Healthy | Drug-free (2 days) | Mixed sample |
| **Keshavan et al., 1998** | 30/30 | 30.9/30.9 | 57/57 | 19 Schizophrenia<br>8 Schizoaffective disorder<br>3 Schizophreniform disorder/Healthy | Drug-free (Average 97.5 weeks;<br>Median 40 weeks) | Mixed sample |
| **Yamashita et al., 1999** | 1 | 34 | 0 | Schizoaffective disorder<br>(with haloperidol-induced akathisia) | Haloperidol + Chlorpromazine<br>+ Biperiden + BZD<br>+ Mood stabilizer<br>→ BZD + Mood stabilizer | Single-case study |
| **Rotenberg et al., 2000** | 20/10 | 45.8/43.3 | 65/60 | 19 Schizophrenia<br>1 Schizoaffective disorder/Healthy | Drug-free (≥2 weeks) | Mixed sample |
| **Wetter et al., 2002** | 1 | 31 | 0 | Schizoaffective disorder<br>(with risperidone-induced<br>restless leg syndrome) | Risperidone<br>+ Mood stabilizers<br>→ Quetiapine<br>+ Mood stabilizers | Single-case study |
| **Armitage et al., 2004** | 14 | 40.9 | 29 | 3 Schizoaffective disorder<br>11 Bipolar I disorder | Carbamazepine, BZD,<br>mood stabilizers<br>→ + Clozapine | Mixed sample |
| **Kluge et al., 2014** | 15 | 32.8 | 47 | Schizophrenia<br>1 Schizoaffective disorder | Drug-free (≥2 days) -><br>Olanzapine | Mixed sample |
| **Kluge et al., 2014** | 15 | 36.7 | 33 | Schizophrenia<br>2 Schizoaffective disorder<br>1 Schizophreniform disorder | Drug-free (≥2 days) -><br>Clozapine | Mixed sample |
| **Göder et al., 2015** | 16/16 | 29.4/28.3 | 56/44 | 14 Chronic schizophrenia<br>2 Schizoaffective disorder/Healthy | Antipsychotics (2° Gen., 94%)<br>Antipsychotics (1° Gen., 6%)<br>+ Antidepressants (13%)<br>+ BZD (6%)<br>+ Anticholinergics (6%)<br>+ Promethazine (6%) | Mixed sample |
| **Kaskie et al., 2019** | 27/23 | 23.2/24.7 | 68/72 | 16 Schizophrenia<br>2 Schizoaffective disorder<br>4 Unspecified psychosis<br>3 MDD with psychotic features<br>2 Bipolar disorder/Healthy | Antipsychotics (2° Gen., 41%)<br>Antipsychotics (3° Gen., 3.7%)<br>+ Antidepressants (83%)<br>+ Mood stabilizers (25%)<br>+ BZD (17%) | Mixed medication<br>status/sample |
|  |  |  |  |  | 15 drug-naïve |  |
| **Gerstenberg et al., 2020** | 12/24 | 16.6/16.5 | 58/46 | 9 Schizophrenia<br>2 Schizoaffective Disorder<br>1 Brief Psychotic Disorder/Healthy | Antipsychotics (3° Gen., 58.3%)<br>Antipsychotics (2°+3° Gen., 25.5%)<br>Antipsychotics (2° Gen., 16.2%)<br>+ BZD (25%)<br>+ Anticholinergics (16.7%) | Mixed sample |
| **Petit et al., 2022** | 12/988 | 58.2/59.9 | 42/58 | Schizoaffective disorder/Healthy | Medicated (25%)<br>Drug-free (75%) | Mixed medication status |
| **Denis et al., 2024** | 17/27 | 21/24 | 56/53 | 7 Schizophrenia<br>5 Schizoaffective disorder<br>4 Bipolar disorder<br>1 Unspecified psychosis/Healthy | Antipsychotics (2° Gen., 35%)<br>Antipsychotics (3° Gen., 29%)<br>Antipsychotics (1° Gen., 12%)<br>+ Mood stabilizers (47%)<br>+ Antidepressants (24%)<br>+ Anticholinergics (18%)<br>+ BZD (6%)<br>4 drug-free | Mixed medication status/sample |

**Table 5.** Non-case-control studies included in the systematic review only.

| Study | N. SZA/Sex/(Age) | Diagnosis | Medication status |
| --- | --- | --- | --- |
| **Hauri et al., 1973** | 2F (28.5) | Schizoaffective disorder | Drug-free (≥1 week) |
| **Gillin et al., 1981** | 3M, 3F (31) | 4 Chronic schizophrenia<br>2 Schizoaffective disorder | Drug-free (≥1 week) |
| **Glassman et al., 1986** | 1M (39) | Schizoaffective disorder | Lithium carbonate, chlorpromazine, triazolam, benztropine<br>(medication-induced somnambulism with REM suppression) |
| **Lahmeyer et al., 1988** | 2F (29) | Schizoaffective disorder | Drug-free (2 weeks) |
| **Keshavan et al., 1991** | 7M, 5F (31.2) | 8 Schizophrenia<br>3 Schizoaffective disorder<br>1 Schizotypal disorder | Drug-free (≥2 weeks)<br>2 drug-naïve |
| **Keshavan et al., 1992** | 14M, 5F (30.5) | 17 Schizophrenia<br>2 Schizoaffective disorder | Drug-free (≥2 weeks)<br>6 drug-naïve |
| **Douglass et al., 1993** | 6? (?) | **Schizoaffective disorder**<br>**Schizophrenia** | Drug-free (≥2 weeks) |
| **Douglass et al., 1993** | 3? (?) | Schizophrenia<br>Schizoaffective disorder | Drug-free (≥2 weeks) |
|  |  | + narcolepsy |  |
| **Keshavan et al., 1994** | 8M, 11F (30.6) | 13 Schizophrenia<br>6 Schizoaffective disorder (suicidal behavior) | Drug-free ($\geq 2$ weeks) |
| **Keshavan et al., 1994** | 12M, 8F (32.3) | 16 Schizophrenia<br>4 Schizoaffective disorder (no suicidal behavior) | Drug-free ($\geq 2$ weeks) |
| **Keshavan et al., 1995** | 10M, 10F (30) | 8 Schizoaffective disorder<br>12 Schizophrenia | Drug-free ($\geq 4$ weeks)<br>12 drug-naïve |
| **Baandrup et al., 2016** | 5M, 5F (49) | 6 Schizophrenia<br>2 Schizoaffective disorder<br>2 Bipolar disorder | Antipsychotics<br>+ BZD<br>+ Antidepressants<br>+ Mood stabilizers<br>+ Anticholinergics |
| **Weinhold et al., 2022** | 9M, 9F (40.7) | 11 Schizophrenia<br>7 Schizoaffective disorder | Atypical antipsychotics<br>+ Antidepressants<br>+ Mood stabilizers |
|  |  |  | **Study summary (no clear sleep data)** |
| **Giedke et al., 1992** | 12M, 18F (48) | 4 Schizoaffective disorder<br>26 MDD | All patients on neuroleptics, antidepressants and BZD<br>Sleep deprivation significantly improved depressed mood |
| **Keshavan et al., 1992** | 11M, 5F (31.65) | 10 Schizophrenia<br>4 Schizoaffective disorder<br>1 Atypical psychosis<br>1 Psychotic affective disorder | Reduced NREM sleep<br>REM activity increased, REML reduced (drug-free $\geq 2$ days)<br>Neuroleptic treatment did not improve sleep |

### 2.1 Eligibility criteria

Studies were assessed as eligible for the meta-analysis based on the following inclusion criteria:

1. Case-control studies with at least one SZA patient sample and one HC sample.
2. Case-control studies with at least one SZA patient sample and one MDD patient sample.
3. Case-control studies with at least one SZA patient sample and one SCZ patient sample.
4. Patient samples in a study had been drug-free for the same amount of time.
5. Samples aged over 13 years.
6. Studies whose patients have been diagnosed by satisfying criteria described in the DSM and/or ICD manual. Research diagnostic criteria (RDC) have been widely used to diagnose SZA and were therefore regarded as eligible.
7. Studies whose patients have had their sleep recorded through polysomnography and scored by using AASM or Rechtschaffen & Kales guidelines. Studies in which sleep stages were scored using early guidelines (e.g. those described by Antrobus et al., Reynolds et al. or Itil et al.) have been regarded as eligible when the adopted scoring methods did not differer from the widely adopted later guidelines.

Studies were excluded based on the following criteria:

1. Studies with no control group.
2. Studies with a control group different than SZA, SCZ, MDD or HC.
3. Studies with samples containing a quota of subjects with diagnoses different from SZA exceeding 5% of the whole sample (e.g. obsessive-compulsive disorder, bipolar disorder, autism spectrum disorder).
4. Studies with a SZA patient sample containing a quota of patients with a different medication status compared to most of the sample and exceeding 5% of the whole sample (e.g. 90% drug-free, 10% treated with at least one antipsychotic).
5. Studies with a SZA patient sample with comorbidities (e.g. narcolepsy, obstructive sleep apnea-hypopnea).
6. Single-case studies.
7. Studies describing sleep changes in SCZ patients without reporting clear sleep data.

The excluded studies were included in the systematic review.

The quality of each study included in either the systematic review or the meta-analysis was assessed using the Newcastle-Ottawa Scale; no study was awarded a score lower than 6.

### 2.2 Data extraction

Data extracted from each study were: sample size, age, gender, diagnosis, medication status, type of medication (if reported), mean and standard deviation (SD) of of TST, LAT, SE, WASO, REMT, %REM, REML, REMD, ST1, ST2, ST3, ST4, %ST1, %ST2, %ST3, %ST4, DELTAT, %DELTA. Some studies did not report SD for their sleep data; therefore SD has been estimated as follows. In each meta-analysis (e.g. REM time, SZA vs HC), the average SD was estimated by calculating the weighted average (based on the number of participants of each study) of all the available SDs. To the studies participating in the very same meta-analysis and whose sleep parameters were reported without SD, this estimated, weighted average SD was attributed. This estimation method for SD is regarded as reliable (Higgins et al., 2024) and is the only estimation process regarding missing data conducted in this study. In a limited number of studies some sleep parameters were not reported clearly but could be extractable by calculating them from other parameters (e.g., REMT = TST/100 x %REM). All these cases of data extraction have been properly highlighted in the supplementary material Table S1, where meta-analysis and systematic review data is displayed in extended form.

### 2.3 Statistical analysis

Jamovi 2.6.44 (desktop Windows x64 solid version) was used to conduct the meta-analysis. A minimum of three data sets from studies was required to perform a meta-analysis for each parameter. The analysis was carried out using the standardized mean difference as the outcome measure. A random-effects model was fitted to the data. The amount of heterogeneity (i.e., tau2), was estimated using the restricted maximum-likelihood estimator. In addition to the estimate of tau2, the Q-test for heterogeneity and the I2 statistic are reported. In case any amount of heterogeneity is detected (i.e., tau2 > 0, regardless of the results of the Q-test), a prediction interval for the true outcomes is also provided. Studentized residuals and Cook’s distances are used to examine whether studies may be outliers and/or influential in the context of the model. Studies with a studentized residual larger than the 100 x (1 - 0.05/(2 X k)) th percentile of a standard normal distribution are considered potential outliers (i.e., using a Bonferroni correction with two-sided alpha = 0.05 for k studies included in the meta-analysis). Studies with a Cook’s distance larger than the median plus six times the interquartile range of the Cook’s distances are considered to be influential. The rank correlation test and the regression test, using the standard error of the observed outcomes as predictor, are used to check for funnel plot asymmetry.

## 3. Results

### 3.1 Study characteristics

A total of 9 studies with 67 SZA patients, 88 SCZ patients, 79 HC and 131 MDD patients were included in the meta-analysis, with some of these studies participating in more than one sleep parameter analysis.

Further 31 studies with 113 SZA patients, 263 SCZ, 1031 HC and 31 MDD patients were included in the systematic review.

Results are summarized in Table 6. Publication bias and funnel plot asymmetry (through rank correlation and regression test) are analyzed in Table 7; outlier and overly influential studies are reported in the very same table. Results for the meta-analyses regarding sleep in drug-free SZA versus HC, drug-free MDD and drug-free SCZ are summarized through forest plots in Figure 2, 3 and 4 respectively.

**Figure 2.**
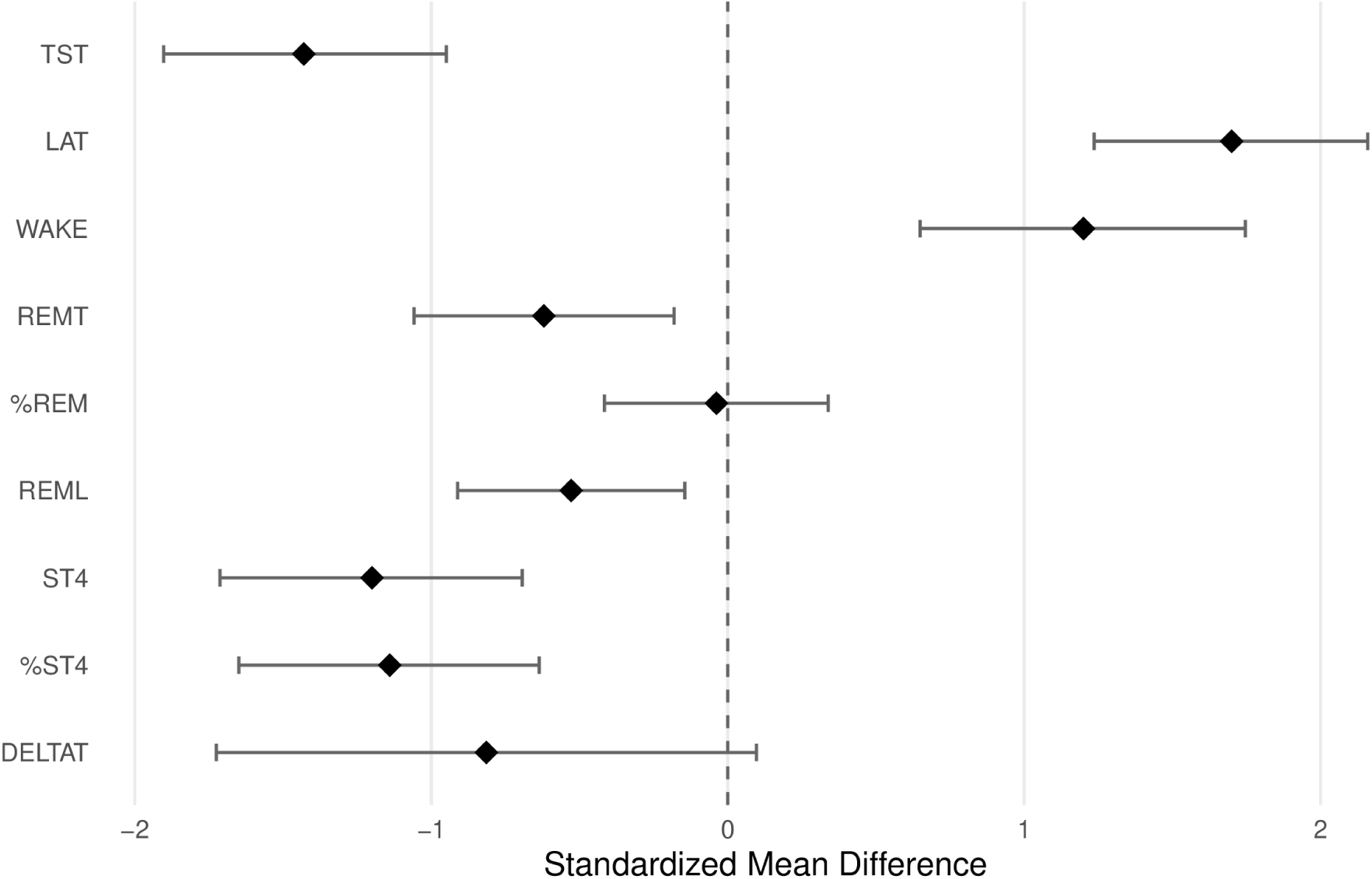
Sleep parameters in drug-free SZA vs HC (forest plot)

**Figure 3.**
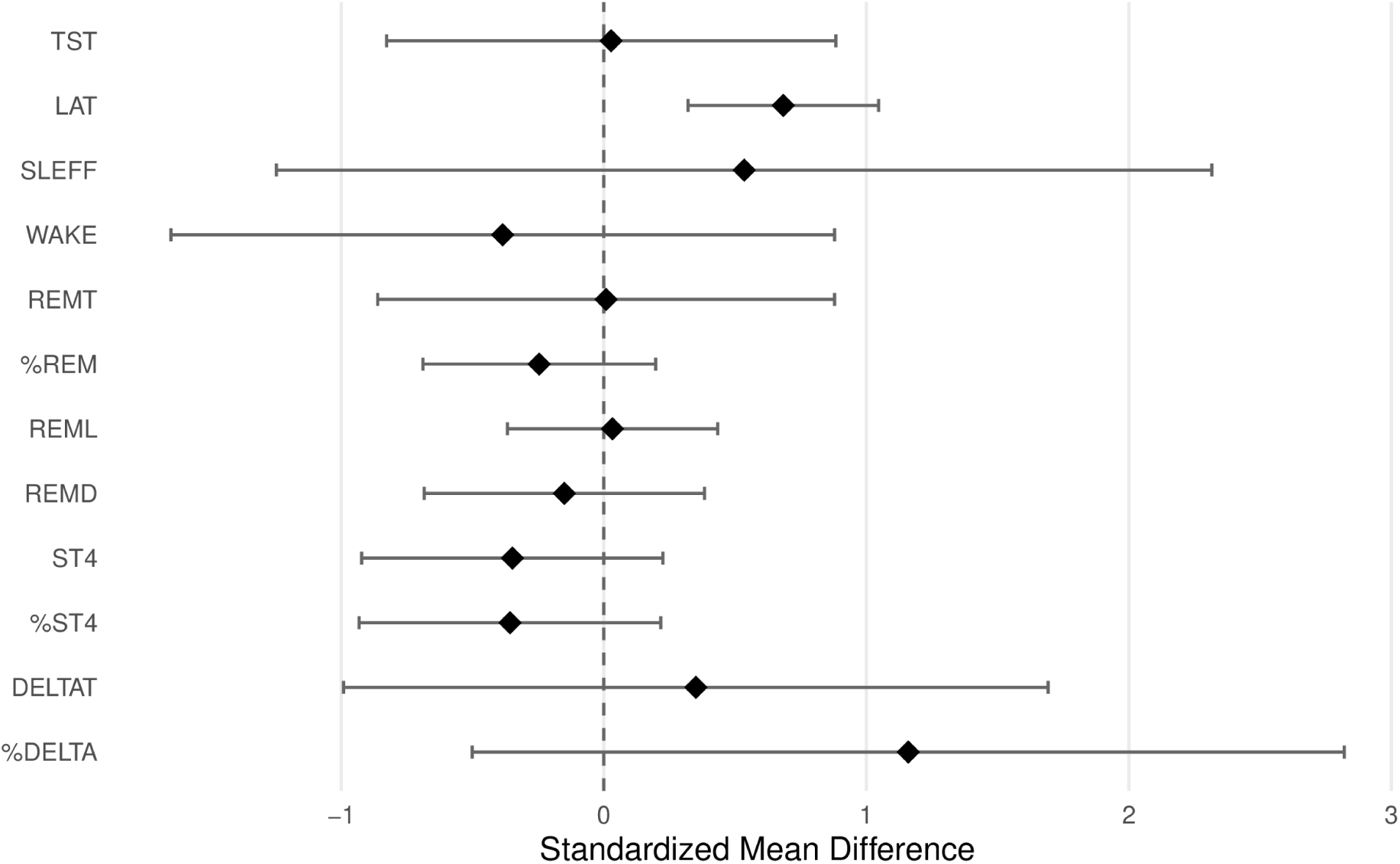
Sleep parameters in drug-free SZA vs drug-free MDD (forest plot)

**Figure 4.**
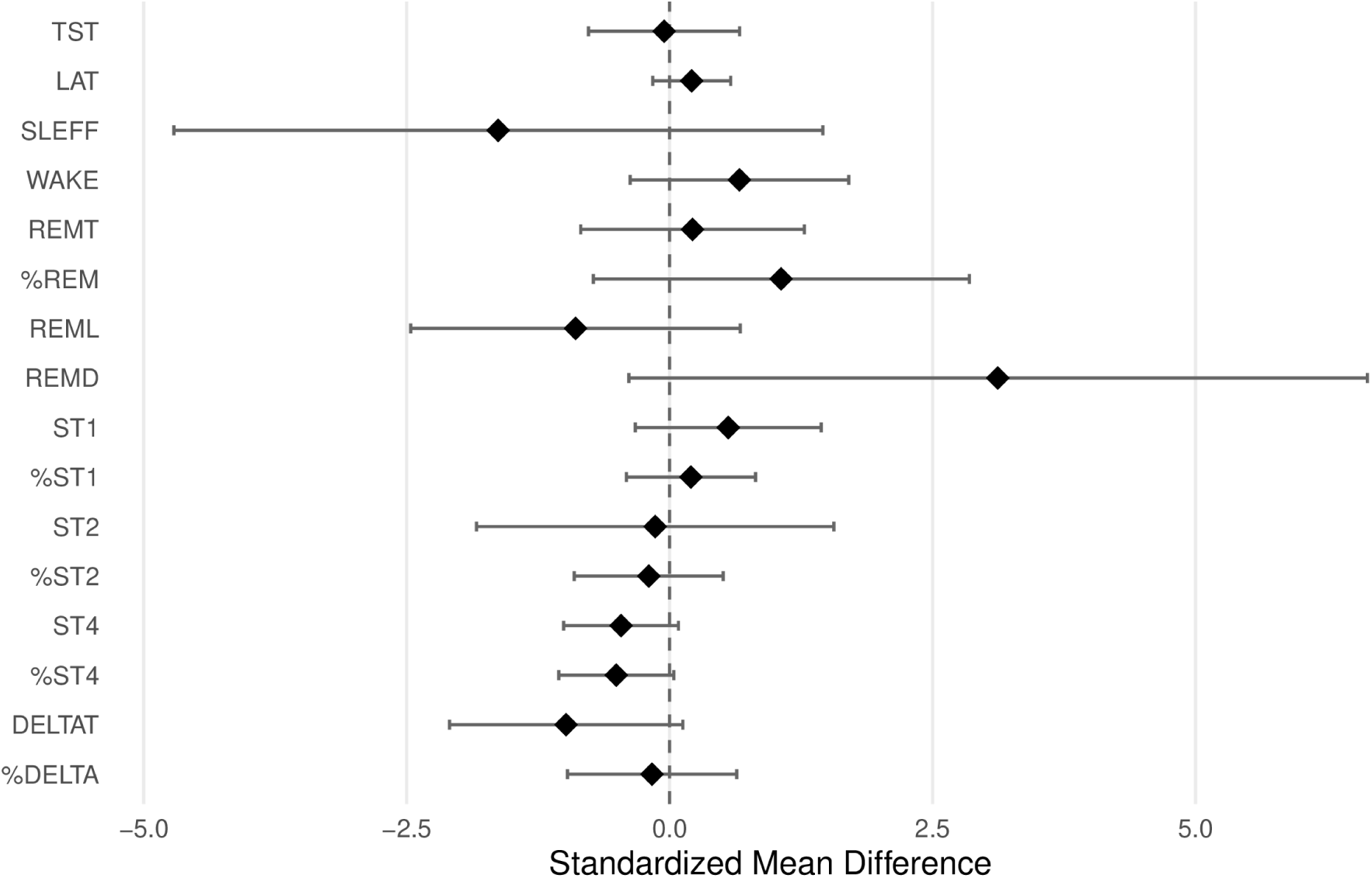
Sleep parameters in drug-free SZA vs drug-free SCZ (forest plot)

**Table 6.** Meta-analysis results.

|  | Meta-analysis |  |  |  |  | Heterogeneity |  |  |
| --- | --- | --- | --- | --- | --- | --- | --- | --- |
| **SZA vs HC** | **N. of studies** | **N. of subjects** | **Mean ± SD** | **RE Model** | **p-value** | **Q** | **I<sup>2</sup>** | **p-value** |
| **Total sleep time** | 4 | 33 vs 60 | 371.29 ± 72.7 vs 443.29 ± 28.72 | -1.43 (-1.903 to -0.949) | **p<0.001** | 2.797 | 0% | p=0.424 |
| **Sleep latency** | 5 | 42 vs 79 | 51.78 ± 39.47 vs 11 ± 8.09 | 1.70 (1.236 to 2.159) | **p<0.001** | 4.379 | 10.43% | p=0.357 |
| **Wakefulness** | 3 | 25 vs 47 | 46.17 ± 42.34 vs 12.83 ± 10.94 | 1.20 (0.649 to 1.746) | **p<0.001** | 2.262 | 6.62% | p=0.323 |
| **REM time** | 4 | 33 vs 60 | 79.7 ± 29.22 vs 96.85 ± 20.05 | -0.620 (-1.058 to -0.181) | **p=0.006** | 1.495 | 0% | p=0.683 |
| **%REM** | 5 | 42 vs 79 | 21.65 ± 5.98 vs 22.3 ± 5.02 | -0.038 (-0.416 to 0.339) | p=0.843 | 1.722 | 0% | p=0.787 |
| **REM latency** | 5 | 42 vs 60 | 66.35 ± 28.33 vs 86.26 ± 34.76 | -0.528 (-0.911 to -0.145) | **p=0.007** | 1.369 | 0% | p=0.849 |
| **Stage 4 sleep** | 3 | 26 vs 51 | 7.65 ± 16.17 vs 47.41 ± 37.00 | -1.20 (-1.713 to -0.693) | **p<0.001** | 0.166 | 0% | p=0.920 |
| **%Stage 4 sleep** | 3 | 26 vs 51 | 1.88 ± 4.63 vs 10.53 ± 8.30 | -1.14 (-1.649 to -0.636) | **p<0.001** | 0.429 | 0% | p=0.807 |
| **Delta sleep** | 3 | 25 vs 47 | 43.16 ± 32.25 vs 74.56 ± 30.90 | -0.814 (-1.725 to 0.097) | p=0.080 | 6.107 | 67.66% | p=0.047 |
| **SZA vs MDD** | **N. of studies** | **N. of subjects** | **Mean ± SD** | **RE Model** | **p-value** | **Q** | **I<sup>2</sup>** | **p-value** |
| **Total sleep time** | 7 | 53 vs 83 | 345.56 ± 67.65 vs 344.43 ± 58.48 | 0.028 (-0.827 to 0.884) | p=0.948 | 21.237 | 80.04% | p=0.002 |
| **Sleep latency** | 7 | 56 vs 80 | 57.82 ± 43.48 vs 37.68 ± 33.80 | 0.684 (0.321 to 1.047) | **p<0.001** | 4.931 | 0% | p=0.553 |
| **Sleep efficiency** | 3 | 26 vs 49 | 68.52 ± 15.42 vs 67.19 ± 17.64 | 0.535 (-1.247 to 2.316) | p=0.556 | 14.418 | 90.44% | p<0.001 |
| **Wakefulness** | 5 | 43 vs 67 | 40.08 ± 37.69 vs 42.41 ± 36.98 | -0.385 (-1.649 to 0.879) | p=0.550 | 19.199 | 88.48% | p<0.001 |
| **REM time** | 7 | 53 vs 83 | 72.89 ± 25.6 vs 78.23 ± 35.85 | 0.009 (-0.861 to 0.879) | p=0.984 | 20.393 | 80.67% | p=0.002 |
| **%REM** | 8 | 62 vs 89 | 21.08 ± 5.65 vs 22.71 ± 6.71 | -0.246 (-0.689 to 0.198) | p=0.277 | 11.842 | 37.83% | p=0.106 |
| **REM latency** | 10 | 67 vs 131 | 54.39 ± 25.57 vs 45.52 ± 22.90 | 0.033 (-0.367 to 0.434) | p=0.870 | 14.28 | 35.00% | p=0.113 |
| **REM density** | 3 | 20 vs 43 | 1.77 ± 0.71 vs 1.84 ± 0.95 | -0.150 (-0.684 to 0.384) | p=0.582 | 0.152 | 0% | p=0.927 |
| **Stage 4 sleep** | 3 | 26 vs 23 | 7.65 ± 16.17 vs 14.78 ± 25.23 | -0.348 (-0.922 to 0.225) | p=0.234 | 1.051 | 0% | p=0.591 |
| **%Stage 4 sleep** | 3 | 26 vs 23 | 1.88 ± 4.63 vs 3.60 ± 5.59 | -0.357 (-0.932 to 0.217) | p=0.223 | 1.281 | 0% | p=0.527 |
| **Delta sleep** | 6 | 49 vs 92 | 26.74 ± 15.60 vs 21.26 ± 12.90 | 0.351 (-0.991 to 1.693) | p=0.609 | 30.314 | 91.28% | p<0.001 |
| **%Delta sleep** | 3 | 26 vs 49 | 5.42 ± 3.28 vs 2.22 ± 2.67 | 1.16 (-0.501 to 2.821) | p=0.171 | 12.554 | 88.30% | p=0.002 |
| **SZA vs SCZ** | **N. of studies** | **N. of subjects** | **Mean ± SD** | **RE Model** | **p-value** | **Q** | **I<sup>2</sup>** | **p-value** |
| **Total sleep time** | 7 | 41 vs 79 | 368.62 ± 65.88 vs 369.82 ± 48.07 | -0.052 (-0.770 to 0.666) | p=0.887 | 15.949 | 66.73% | p=0.014 |
| **Sleep latency** | 6 | 44 vs 65 | 58.06 ± 36.12 vs 57.30 ± 36.78 | 0.210 (-0.160 to 0.581) | p=0.266 | 20.661 | 0% | p=0.004 |
| **Sleep efficiency** | 3 | 14 vs 41 | 71.80 ± 14.40 vs 78.63 ± 7.47 | -1.63 (-4.714 to 1.458) | p=0.301 | 14.999 | 93.61% | p<0.001 |
| **Wakefulness** | 5 | 31 vs 62 | 40.08 ± 32.34 vs 21.16 ± 14.97 | 0.664 (-0.375 to 1.703) | p=0.210 | 14.377 | 77.93% | p=0.006 |
| **REM time** | 7 | 41 vs 79 | 80.4 ± 25.6 vs 83.17 ± 18.53 | 0.218 (-0.845 to 1.281) | p=0.688 | 24.28 | 84.39% | p<0.001 |
| **%REM** | 8 | 50 vs 88 | 21.89 ± 5.36 vs 19.74 ± 2.86 | 1.06 (-0.724 to 2.850) | p=0.244 | 61.892 | 94.47% | p<0.001 |
| **REM latency** | 8 | 50 vs 88 | 62.27 ± 24.85 vs 67.15 ± 19.65 | -0.894 (-2.461 to 0.672) | p=0.263 | 41.256 | 93.60% | p<0.001 |
| **REM density** | 3 | 8 vs 30 | 2.31 ± 0.20 vs 1.82 ± 0.10 | 3.12 (-0.388 to 6.635) | p=0.081 | 21.365 | 89.27% | p<0.001 |
| **Stage 1 sleep** | 3 | 14 vs 41 | 43.48 ± 8.9 ± 37.74 ± 4.69 | 0.558 (-0.326 to 1.441) | p=0.216 | 3.517 | 44.12% | p=0.172 |
| **%Stage 1 sleep** | 3 | 14 vs 41 | 12.63 ± 5.6 vs 10.76 ± 4.70 | 0.203 (-0.411 to 0.817) | p=0.516 | 0.161 | 0% | p=0.923 |
| **Stage 2 sleep** | 4 | 23 vs 50 | 194.74 ± 18.30 vs 201.45 ± 11.41 | -0.137 (-1.836 to 1.561) | p=0.874 | 26.218 | 88.52% | p<0.001 |
| **%Stage 2 sleep** | 3 | 14 vs 41 | 56.02 ± 6.20 vs 58.83 ± 6.30 | -0.198 (-0.907 to 0.510) | p=0.583 | 2.372 | 19.39% | p=0.305 |
| **Stage 4 sleep** | 3 | 26 vs 28 | 7.65 ± 16.17 vs 17.38 ± 26.08 | -0.461 (-1.007 to 0.084) | p=0.097 | 0.862 | 0% | p=0.650 |
| **%Stage 4 sleep** | 3 | 26 vs 28 | 1.88 ± 4.63 vs 4.68 ± 6.55 | -0.507 (-1.054 to 0.040) | p=0.069 | 1.101 | 0% | p=0.577 |
| **Delta sleep** | 5 | 31 vs 62 | 38.33 ± 20.36 vs 42.39 ± 15.50 | -0.984 (-2.093 to 0.126) | p=0.082 | 16.348 | 80.35% | p=0.003 |
| **%Delta sleep** | 3 | 14 vs 41 | 9.91 ± 8.19 ± 10.25 ± 7.20 | -0.167 (-0.972 to 0.638) | p=0.685 | 2.916 | 35.41% | p=0.233 |

**Table 7.**
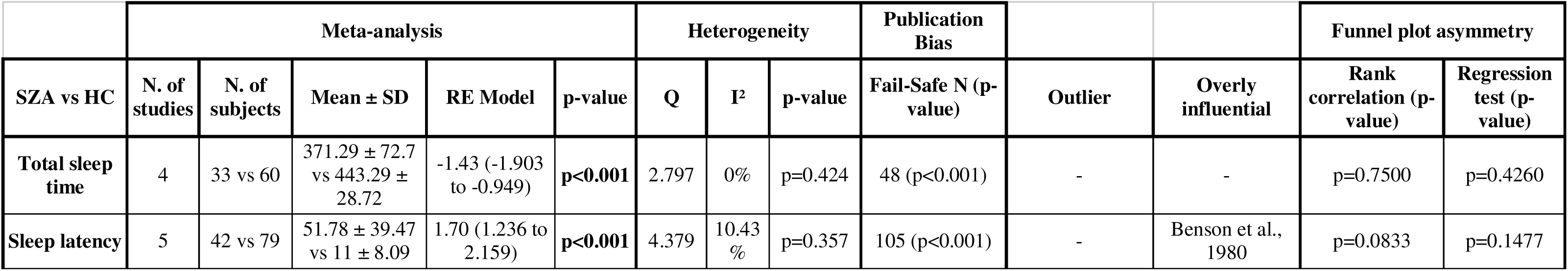
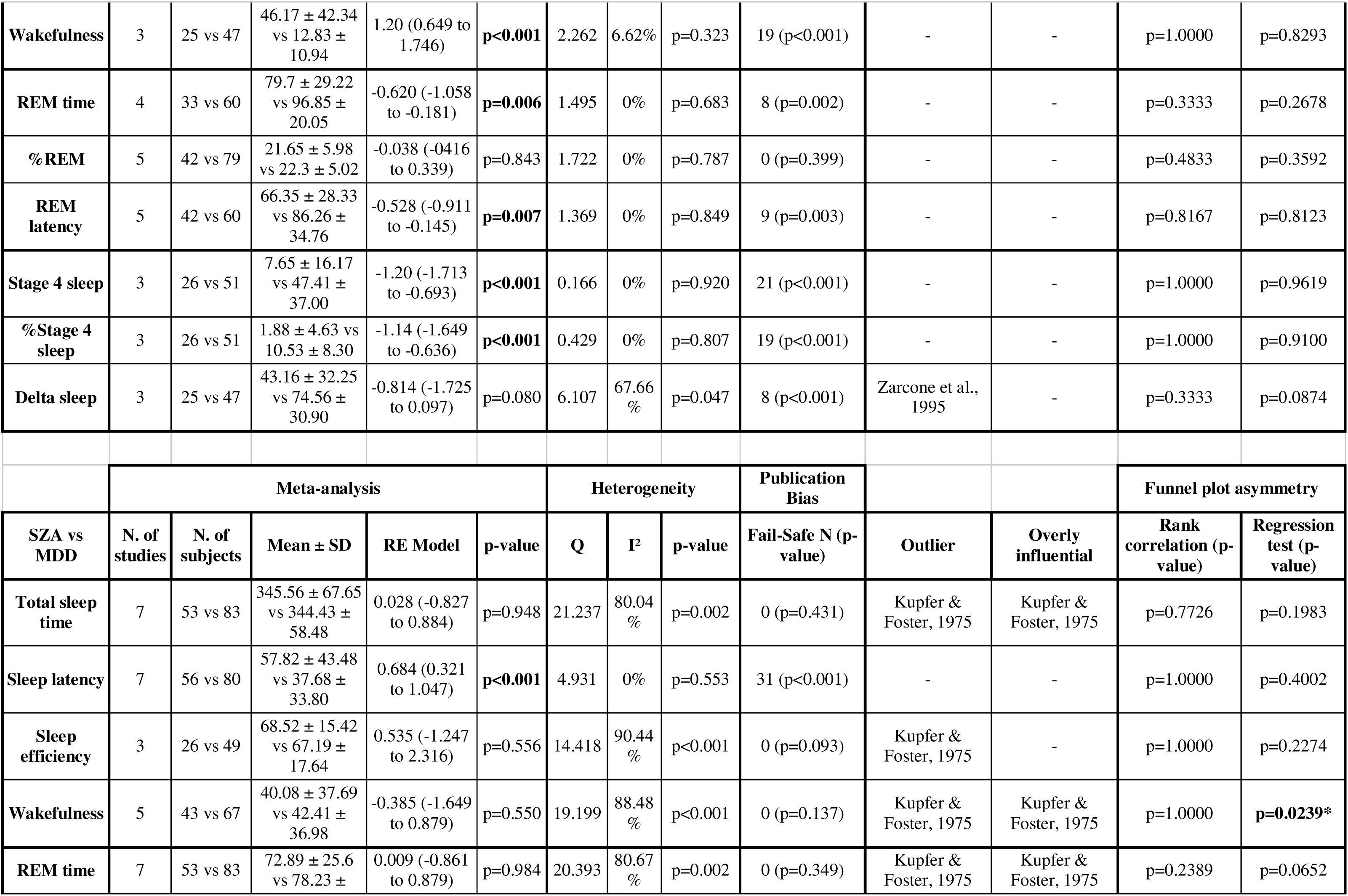
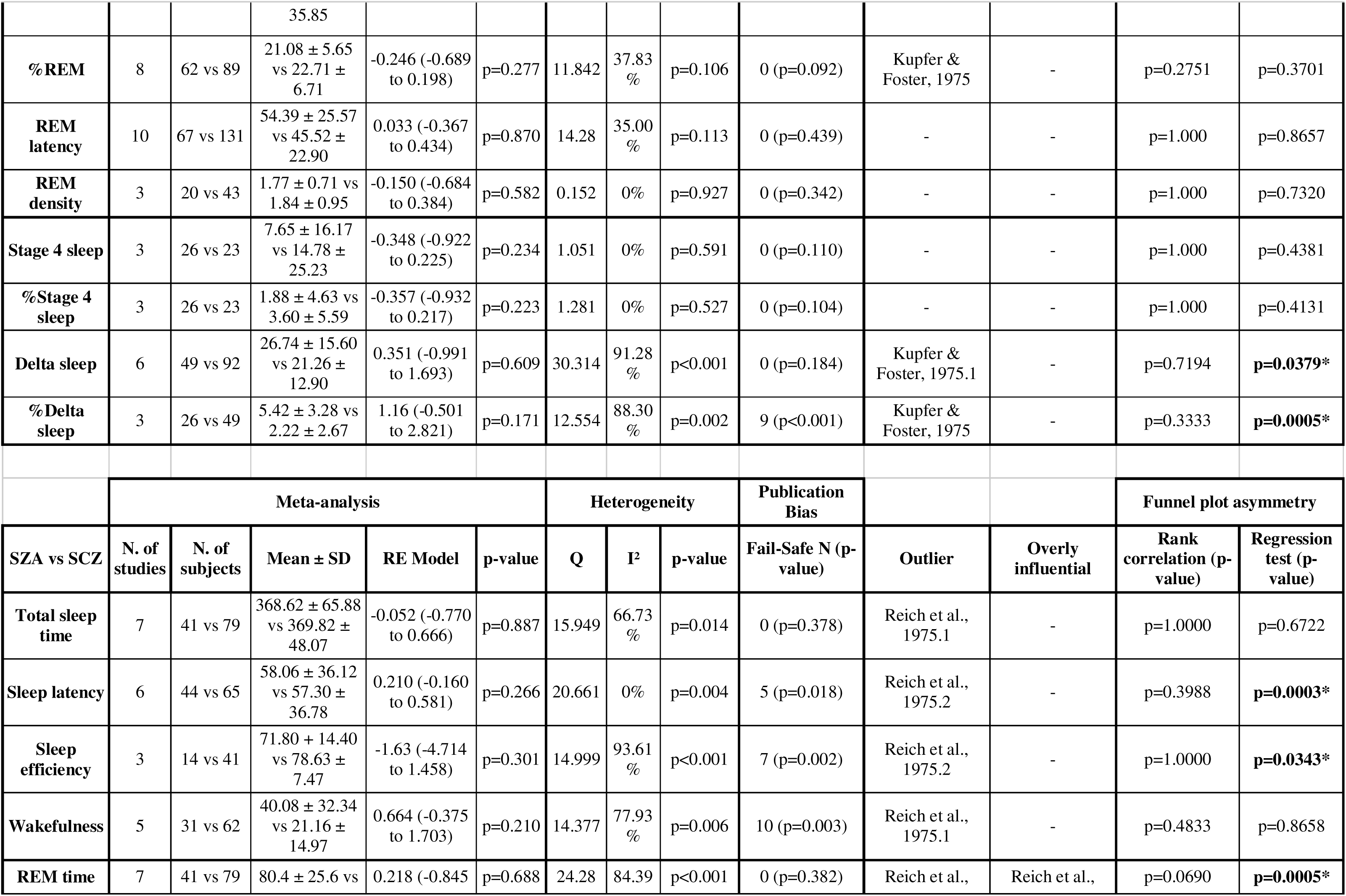
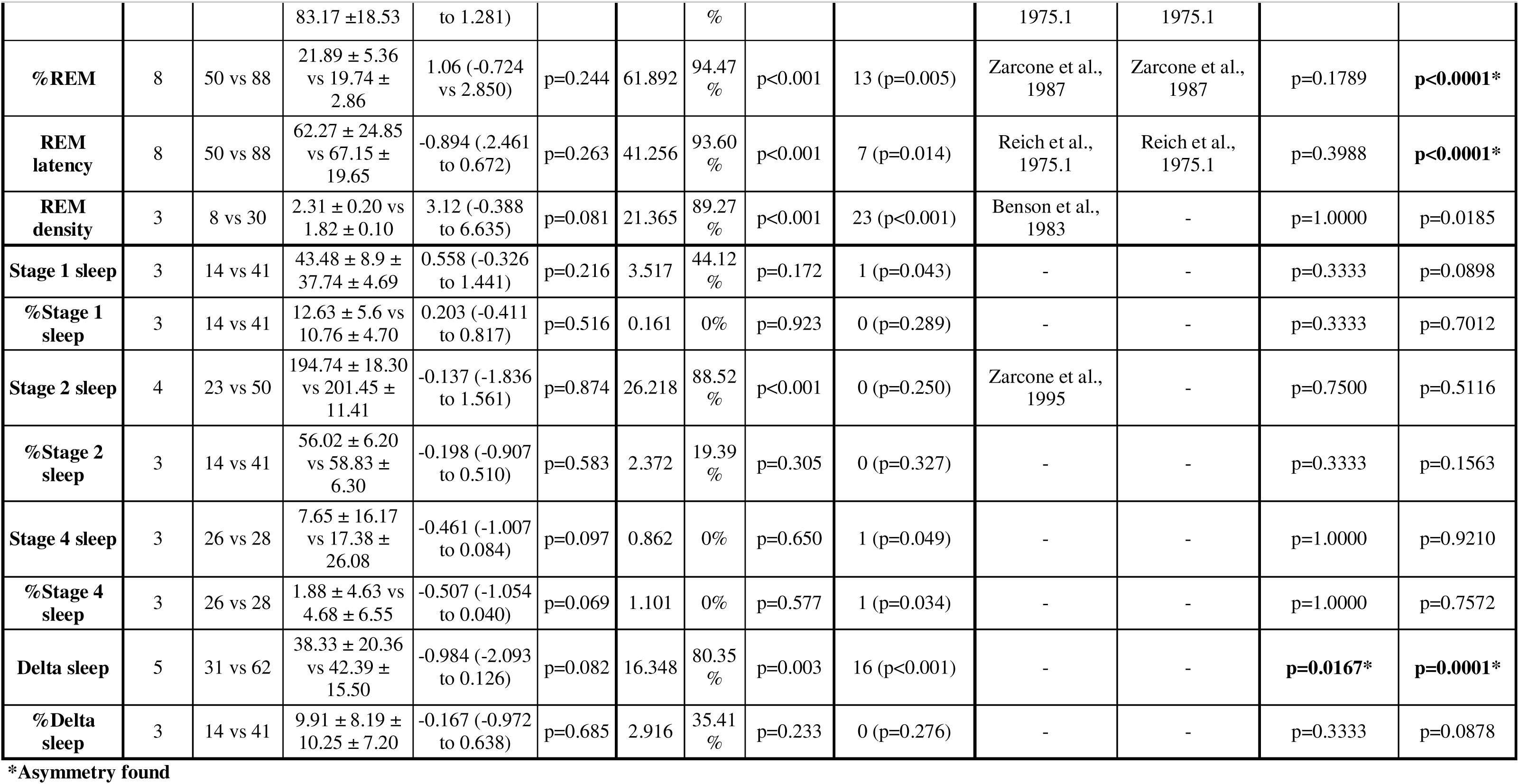
Publication bias and funnel plot asymmetry (rank correlation and regression test) analysis.

|  | Meta-analysis |  |  |  |  | Heterogeneity |  |  | Publication Bias |  |  | Funnel plot asymmetry |  |
| --- | --- | --- | --- | --- | --- | --- | --- | --- | --- | --- | --- | --- | --- |
| SZA vs HC | N. of studies | N. of subjects | Mean ± SD | RE Model | p-value | Q | I<sup>2</sup> | p-value | Fail-Safe N (p-value) | Outlier | Overly influential | Rank correlation (p-value) | Regression test (p-value) |
| **Total sleep time** | 4 | 33 vs 60 | 371.29 ± 72.7 vs 443.29 ± 28.72 | -1.43 (-1.903 to -0.949) | **p<0.001** | 2.797 | 0% | p=0.424 | 48 (p<0.001) | - | - | p=0.7500 | p=0.4260 |
| **Sleep latency** | 5 | 42 vs 79 | 51.78 ± 39.47 vs 11 ± 8.09 | 1.70 (1.236 to 2.159) | **p<0.001** | 4.379 | 10.43 % | p=0.357 | 105 (p<0.001) | - | Benson et al., 1980 | p=0.0833 | p=0.1477 |
| Wakefulness | 3 | 25 vs 47 | 46.17 ± 42.34<br>vs 12.83 ± 10.94 | 1.20 (0.649 to 1.746) | **p<0.001** | 2.262 | 6.62% | p=0.323 | 19 (p<0.001) | - | - | p=1.0000 | p=0.8293 |
| REM time | 4 | 33 vs 60 | 79.7 ± 29.22<br>vs 96.85 ± 20.05 | -0.620 (-1.058 to -0.181) | **p=0.006** | 1.495 | 0% | p=0.683 | 8 (p=0.002) | - | - | p=0.3333 | p=0.2678 |
| %REM | 5 | 42 vs 79 | 21.65 ± 5.98<br>vs 22.3 ± 5.02 | -0.038 (-0.416 to 0.339) | p=0.843 | 1.722 | 0% | p=0.787 | 0 (p=0.399) | - | - | p=0.4833 | p=0.3592 |
| REM latency | 5 | 42 vs 60 | 66.35 ± 28.33<br>vs 86.26 ± 34.76 | -0.528 (-0.911 to -0.145) | **p=0.007** | 1.369 | 0% | p=0.849 | 9 (p=0.003) | - | - | p=0.8167 | p=0.8123 |
| Stage 4 sleep | 3 | 26 vs 51 | 7.65 ± 16.17<br>vs 47.41 ± 37.00 | -1.20 (-1.713 to -0.693) | **p<0.001** | 0.166 | 0% | p=0.920 | 21 (p<0.001) | - | - | p=1.0000 | p=0.9619 |
| %Stage 4 sleep | 3 | 26 vs 51 | 1.88 ± 4.63 vs 10.53 ± 8.30 | -1.14 (-1.649 to -0.636) | **p<0.001** | 0.429 | 0% | p=0.807 | 19 (p<0.001) | - | - | p=1.0000 | p=0.9100 |
| Delta sleep | 3 | 25 vs 47 | 43.16 ± 32.25<br>vs 74.56 ± 30.90 | -0.814 (-1.725 to 0.097) | p=0.080 | 6.107 | 67.66 % | p=0.047 | 8 (p<0.001) | Zarcone et al., 1995 | - | p=0.3333 | p=0.0874 |
|  | Meta-analysis |  |  |  |  | Heterogeneity |  |  | Publication Bias |  |  | Funnel plot asymmetry |  |
| SZA vs MDD | N. of studies | N. of subjects | Mean ± SD | RE Model | p-value | Q | I<sup>2</sup> | p-value | Fail-Safe N (p-value) | Outlier | Overly influential | Rank correlation (p-value) | Regression test (p-value) |
| Total sleep time | 7 | 53 vs 83 | 345.56 ± 67.65<br>vs 344.43 ± 58.48 | 0.028 (-0.827 to 0.884) | p=0.948 | 21.237 | 80.04 % | p=0.002 | 0 (p=0.431) | Kupfer & Foster, 1975 | Kupfer & Foster, 1975 | p=0.7726 | p=0.1983 |
| Sleep latency | 7 | 56 vs 80 | 57.82 ± 43.48<br>vs 37.68 ± 33.80 | 0.684 (0.321 to 1.047) | **p<0.001** | 4.931 | 0% | p=0.553 | 31 (p<0.001) | - | - | p=1.0000 | p=0.4002 |
| Sleep efficiency | 3 | 26 vs 49 | 68.52 ± 15.42<br>vs 67.19 ± 17.64 | 0.535 (-1.247 to 2.316) | p=0.556 | 14.418 | 90.44 % | p<0.001 | 0 (p=0.093) | Kupfer & Foster, 1975 | - | p=1.0000 | p=0.2274 |
| Wakefulness | 5 | 43 vs 67 | 40.08 ± 37.69<br>vs 42.41 ± 36.98 | -0.385 (-1.649 to 0.879) | p=0.550 | 19.199 | 88.48 % | p<0.001 | 0 (p=0.137) | Kupfer & Foster, 1975 | Kupfer & Foster, 1975 | p=1.0000 | **p=0.0239\*** |
| REM time | 7 | 53 vs 83 | 72.89 ± 25.6<br>vs 78.23 ± | 0.009 (-0.861 to 0.879) | p=0.984 | 20.393 | 80.67 % | p=0.002 | 0 (p=0.349) | Kupfer & Foster, 1975 | Kupfer & Foster, 1975 | p=0.2389 | p=0.0652 |
|  |  |  | 35.85 |  |  |  |  |  |  |  |  |  |  |
| **%REM** | 8 | 62 vs 89 | 21.08 ± 5.65<br>vs 22.71 ± 6.71 | -0.246 (-0.689<br>to 0.198) | p=0.277 | 11.842 | 37.83<br>% | p=0.106 | 0 (p=0.092) | Kupfer &<br>Foster, 1975 | - | p=0.2751 | p=0.3701 |
| **REM**<br>**latency** | 10 | 67 vs 131 | 54.39 ± 25.57<br>vs 45.52 ± 22.90 | 0.033 (-0.367<br>to 0.434) | p=0.870 | 14.28 | 35.00<br>% | p=0.113 | 0 (p=0.439) | - | - | p=1.000 | p=0.8657 |
| **REM**<br>**density** | 3 | 20 vs 43 | 1.77 ± 0.71 vs<br>1.84 ± 0.95 | -0.150 (-0.684<br>to 0.384) | p=0.582 | 0.152 | 0% | p=0.927 | 0 (p=0.342) | - | - | p=1.000 | p=0.7320 |
| **Stage 4 sleep** | 3 | 26 vs 23 | 7.65 ± 16.17<br>vs 14.78 ± 25.23 | -0.348 (-0.922<br>to 0.225) | p=0.234 | 1.051 | 0% | p=0.591 | 0 (p=0.110) | - | - | p=1.000 | p=0.4381 |
| **%Stage 4**<br>**sleep** | 3 | 26 vs 23 | 1.88 ± 4.63 vs<br>3.60 ± 5.59 | -0.357 (-0.932<br>to 0.217) | p=0.223 | 1.281 | 0% | p=0.527 | 0 (p=0.104) | - | - | p=1.000 | p=0.4131 |
| **Delta sleep** | 6 | 49 vs 92 | 26.74 ± 15.60<br>vs 21.26 ± 12.90 | 0.351 (-0.991<br>to 1.693) | p=0.609 | 30.314 | 91.28<br>% | p<0.001 | 0 (p=0.184) | Kupfer &<br>Foster, 1975.1 | - | p=0.7194 | **p=0.0379\*** |
| **%Delta**<br>**sleep** | 3 | 26 vs 49 | 5.42 ± 3.28 vs<br>2.22 ± 2.67 | 1.16 (-0.501<br>to 2.821) | p=0.171 | 12.554 | 88.30<br>% | p=0.002 | 9 (p<0.001) | Kupfer &<br>Foster, 1975 | - | p=0.3333 | **p=0.0005\*** |
|  | **Meta-analysis** |  |  |  |  | **Heterogeneity** |  |  | **Publication**<br>**Bias** |  |  | **Funnel plot asymmetry** |  |
| **SZA vs SCZ** | **N. of**<br>**studies** | **N. of**<br>**subjects** | **Mean ± SD** | **RE Model** | **p-value** | **Q** | **I<sup>2</sup>** | **p-value** | **Fail-Safe N (p-**<br>**value)** | **Outlier** | **Overly**<br>**influential** | **Rank**<br>**correlation (p-**<br>**value)** | **Regression**<br>**test (p-**<br>**value)** |
| **Total sleep**<br>**time** | 7 | 41 vs 79 | 368.62 ± 65.88<br>vs 369.82 ± 48.07 | -0.052 (-0.770<br>to 0.666) | p=0.887 | 15.949 | 66.73<br>% | p=0.014 | 0 (p=0.378) | Reich et al.,<br>1975.1 | - | p=1.0000 | p=0.6722 |
| **Sleep latency** | 6 | 44 vs 65 | 58.06 ± 36.12<br>vs 57.30 ± 36.78 | 0.210 (-0.160<br>to 0.581) | p=0.266 | 20.661 | 0% | p=0.004 | 5 (p=0.018) | Reich et al.,<br>1975.2 | - | p=0.3988 | **p=0.0003\*** |
| **Sleep**<br>**efficiency** | 3 | 14 vs 41 | 71.80 ± 14.40<br>vs 78.63 ± 7.47 | -1.63 (-4.714<br>to 1.458) | p=0.301 | 14.999 | 93.61<br>% | p<0.001 | 7 (p=0.002) | Reich et al.,<br>1975.2 | - | p=1.0000 | **p=0.0343\*** |
| **Wakefulness** | 5 | 31 vs 62 | 40.08 ± 32.34<br>vs 21.16 ± 14.97 | 0.664 (-0.375<br>to 1.703) | p=0.210 | 14.377 | 77.93<br>% | p=0.006 | 10 (p=0.003) | Reich et al.,<br>1975.1 | - | p=0.4833 | p=0.8658 |
| **REM time** | 7 | 41 vs 79 | 80.4 ± 25.6 vs | 0.218 (-0.845 | p=0.688 | 24.28 | 84.39 | p<0.001 | 0 (p=0.382) | Reich et al., | Reich et al., | p=0.0690 | **p=0.0005\*** |
|  |  |  | 83.17 ±18.53 | to 1.281) |  |  | % |  |  | 1975.1 | 1975.1 |  |  |
| **%REM** | 8 | 50 vs 88 | 21.89 ± 5.36<br>vs 19.74 ±<br>2.86 | 1.06 (-0.724<br>vs 2.850) | p=0.244 | 61.892 | 94.47<br>% | p<0.001 | 13 (p=0.005) | Zarcone et al.,<br>1987 | Zarcone et al.,<br>1987 | p=0.1789 | **p<0.0001\*** |
| **REM**<br>**latency** | 8 | 50 vs 88 | 62.27 ± 24.85<br>vs 67.15 ±<br>19.65 | -0.894 (.2461<br>to 0.672) | p=0.263 | 41.256 | 93.60<br>% | p<0.001 | 7 (p=0.014) | Reich et al.,<br>1975.1 | Reich et al.,<br>1975.1 | p=0.3988 | **p<0.0001\*** |
| **REM**<br>**density** | 3 | 8 vs 30 | 2.31 ± 0.20 vs<br>1.82 ± 0.10 | 3.12 (-0.388<br>to 6.635) | p=0.081 | 21.365 | 89.27<br>% | p<0.001 | 23 (p<0.001) | Benson et al.,<br>1983 | - | p=1.0000 | p=0.0185 |
| **Stage 1 sleep** | 3 | 14 vs 41 | 43.48 ± 8.9 ±<br>37.74 ± 4.69 | 0.558 (-0.326<br>to 1.441) | p=0.216 | 3.517 | 44.12<br>% | p=0.172 | 1 (p=0.043) | - | - | p=0.3333 | p=0.0898 |
| **%Stage 1**<br>**sleep** | 3 | 14 vs 41 | 12.63 ± 5.6 vs<br>10.76 ± 4.70 | 0.203 (-0.411<br>to 0.817) | p=0.516 | 0.161 | 0% | p=0.923 | 0 (p=0.289) | - | - | p=0.3333 | p=0.7012 |
| **Stage 2 sleep** | 4 | 23 vs 50 | 194.74 ± 18.30<br>vs 201.45 ±<br>11.41 | -0.137 (-1.836<br>to 1.561) | p=0.874 | 26.218 | 88.52<br>% | p<0.001 | 0 (p=0.250) | Zarcone et al.,<br>1995 | - | p=0.7500 | p=0.5116 |
| **%Stage 2**<br>**sleep** | 3 | 14 vs 41 | 56.02 ± 6.20<br>vs 58.83 ±<br>6.30 | -0.198 (-0.907<br>to 0.510) | p=0.583 | 2.372 | 19.39<br>% | p=0.305 | 0 (p=0.327) | - | - | p=0.3333 | p=0.1563 |
| **Stage 4 sleep** | 3 | 26 vs 28 | 7.65 ± 16.17<br>vs 17.38 ±<br>26.08 | -0.461 (-1.007<br>to 0.084) | p=0.097 | 0.862 | 0% | p=0.650 | 1 (p=0.049) | - | - | p=1.0000 | p=0.9210 |
| **%Stage 4**<br>**sleep** | 3 | 26 vs 28 | 1.88 ± 4.63 vs<br>4.68 ± 6.55 | -0.507 (-1.054<br>to 0.040) | p=0.069 | 1.101 | 0% | p=0.577 | 1 (p=0.034) | - | - | p=1.0000 | p=0.7572 |
| **Delta sleep** | 5 | 31 vs 62 | 38.33 ± 20.36<br>vs 42.39 ±<br>15.50 | -0.984 (-2.093<br>to 0.126) | p=0.082 | 16.348 | 80.35<br>% | p=0.003 | 16 (p<0.001) | - | - | **p=0.0167\*** | **p=0.0001\*** |
| **%Delta**<br>**sleep** | 3 | 14 vs 41 | 9.91 ± 8.19 ±<br>10.25 ± 7.20 | -0.167 (-0.972<br>to 0.638) | p=0.685 | 2.916 | 35.41<br>% | p=0.233 | 0 (p=0.276) | - | - | p=0.3333 | p=0.0878 |

\*Asymmetry found

### 3.2 Total sleep time

Drug-free SZA showed reduced TST (SMD=-1.43; p<0.001) compared to HC. TST of drug-free SZA did not differ from that of drug-free MDD (SMD=0.0286; p=0.948) and drug-free SCZ (SMD=-0.0519; p=0.887).

### 3.3 Sleep latency

Drug-free SZA showed prolonged LAT compared to both HC (SMD=1.70; p<0.001) and drug-free MDD (SMD=0.684; p<0.0001). LAT of drug-free SZA did not differ from that of drug-free SCZ (SMD=0.210; p=0.887).

### 3.4 Sleep efficiency

Drug-free SZA SE did not differ from that of drug-free MDD (SMD=0.535; p=0.556) and drug-free SCZ (SMD=-1.63; p=0.301). It was not possible to compare drug-free SZA and HC SE due to lack of data.

### 3.5 Wakefulness time during sleep

Drug-free SZA showed increased WASO compared to HC (SMD=1.20; p<0.001). WASO of drug-free SZA did not differ from that of drug-free MDD (SMD=-0.385; p=0.550) and drug-free SCZ (SMD=0.664; p=0.210).

### 3.6 REM time

Drug-free SZA showed reduced REMT compared to HC (SMD=-0.620; p=0.006). REMT of drug-free SZA did not differ from that of drug-free MDD (SMD=0.009; p=0.984) and drug-free SCZ (SMD=0.218; p=0.688).

### 3.7 REM percentage

Drug-free SZA %REM did not differ from that of HC (SMD=-0.0382; p=0.843), drug-free MDD (SMD=-0.246; p=0.277) and drug-free SCZ (SMD=1.06; p=0.244).

### 3.8 REM latency

Drug-free SZA showed shortened REML compared to HC (SMD=-0.528; p=0.007). REML of drug-free SZA did not differ from that of drug-free MDD (SMD=0.0335; p=0.870) and drug-free SCZ (SMD=-0.894; p=0.263).

### 3.9 REM density

Drug-free SZA REMD did not differ from that of drug-free MDD (SMD=-0.150; p=0.582) and drug-free SCZ (SMD=3.12; p=0.081). It was not possible to compare drug-free SZA and HC REMD due to lack of data.

### 3.10 Stage 1 sleep time

Drug-free SZA ST1 did not differ from that of drug-free SCZ (SMD=0.0558; p=0.216). It was not possible to compare drug-free SZA ST1 to that of HC and drug-free MDD due to lack of data.

### 3.11 Stage 1 sleep percentage

Drug-free SZA %ST1 did not differ from that of drug-free SCZ (SMD=0.203; p=0.516). It was not possible to compare drug-free SZA %ST1 to that of HC and drug-free MDD due to lack of data.

### 3.12 Stage 2 sleep time

Drug-free SZA ST2 did not differ from that of drug-free SCZ (SMD=-0.137; p=0.874). It was not possible to compare drug-free SZA ST2 to that of HC and drug-free MDD due to lack of data.

### 3.13 Stage 2 sleep percentage

Drug-free SZA %ST2 did not differ from that of drug-free SCZ (SMD=-0.198; p=0.583). It was not possible to compare drug-free SZA %ST2 to that of HC and drug-free MDD due to lack of data.

### 3.14 Stage 3 sleep time

It was not possible to perform a meta-analysis for ST3 in any case-control due to lack of data.

### 3.15 Stage 3 sleep percentage

It was not possible to perform a meta-analysis for %ST3 in any case-control due to lack of data.

### 3.16 Stage 4 sleep time

Drug-free SZA showed reduced ST4 compared to HC (SMD=-1.20; p<0.001). ST4 of drug-free SZA did not differ from that of drug-free MDD (SMD=-0.348; p=0.234) and drug-free SCZ (SMD=-0.461; p=0.097).

### 3.17 Stage 4 sleep percentage

Drug-free SZA showed reduced %ST4 compared to HC (SMD=-1.14; p<0.001). %ST4 of drug-free SZA did not differ from that of drug-free MDD (SMD=-0.357; p=0.223) and drug-free SCZ (SMD=-0.507; p=0.069).

### 3.18 Delta sleep time

Drug-free SZA DELTAT did not differ from that of HC (SMD=-0.814; p=0.080), drug-free MDD (SMD=0.351; p=0.609) and drug-free SCZ (SMD=-0.984; p=0.082).

### 3.19 Delta sleep percentage

Drug-free SZA %DELTA did not differ from that of drug-free MDD (SMD=1.16; p=0.171) and drug-free SCZ (SMD=-0.167; p=0.685). It was not possible to compare drug-free SZA %DELTA to that of HC due to lack of data.

## 4. Discussion

### 4.1 Reviews and meta-analyses

Literature on sleep including samples with only SZA patients is limited, with most of the studies including SZA patients together with SCZ patients; therefore, it is difficult to clearly understand which sleep disturbances and abnormalities occur in SZA and their prevalence, as it is difficult to understand whether SZA sleep resembles more that of SCZ or that of affective disorders. The present work is the only systematic review and meta-analysis to date regarding sleep in SZA. Very few literature reviews on SZA have been performed, with each of them suggesting the necessity of more studies focused on these patients’ sleep (Clayton, 1982, Meltzer, 1984); however, case-control sleep studies providing clear, drug-free SZA patients sample can be found only in the 1975-1995 span.

A previous meta-analysis performed by the same authors regarding REM sleep in SCZ identified several SZA case-control studies included in this study; however, these were eventually excluded from the meta-analysis (Morra & Barbato, 2025).

The debate surrounding whether SZA exists as a separate nosography entity or not, as previously reported, might help understand the loss of interest towards this investigation. Our meta-analysis found that SZA show decreased TST, increased LAT and WASO compared to HC, confirming the existence of sleep fragmentation in these patients. LAT seems to be longer in SZA compared to that of MDD as well. REMT and REML are respectively reduced and shortened in SZA compared to HC; similarly, ST4 and %ST4 are sensibly reduced in SZA compared to HC. No other difference could be found when comparing SZA sleep to that of SCZ or MDD. Although our meta-analysis did not find a statistically significant difference, DELTAT might be reduced in SZA compared to HC (p=0.08). Moreover, REMD might be increased in SZA compared to SCZ (p=0.08), strengthening the idea that REMD might be strictly connected to emotional processing (Del Giudice et al., 2026) and be similarly increased in SZA and MDD beyond HC and non-affective psychoses levels (Kupfer & Foster, 1975). REML was previously found to clearly differentiate SZA from SCZ, being more like that of MDD and BD (Reich et al., 1975; Kupfer & Foster, 1975) and inversely correlating with affective symptoms severity (Reich et al., 1975). This correlation between REML and depressive symptoms in particular has been successively suggested to be not helpful in differentiating SZA, SCZ and MDD being depressive symptoms largely present in all these patients (Zarcone et al., 1987). Petit et al. argued that it might be even not possible to differentiate SCZ, SZA and BD type I and II patients by sleep parameters, for they estimated a sleep disturbances prevalence of 30-45% even in the healthy population (Petit et al., 2022).

According to our results, sleep variables do not allow to clearly characterize SZA patients from either SCZ and MDD patients.

### 4.2 Limitations and future perspectives

Our meta-analysis indeed presented results that often carried high heterogeneity, a limitation unfortunately common when studying the psychiatric population.

The majority of the meta-analyses performed in this study was based on a number of datasets between three (the minimum amount necessary to perform a meta-analysis as stated in the methods section) and five: this reflects the shortage of eligible studies to perform such analysis. The meta-analyis regarding REML in SZA vs MDD was the only that could boast the highest number of datasets included, ten. Several meta-analyses, such as that regarding %DELTA in SCZ vs HC or those concerning the first two stages of sleep in SCZ vs MDD could not be performed due to the datasets available being less than three. Moreover, it was not possible in any case to perform a single meta-analysis regarding sleep stage 3.

These limitations amount to a partial perspective on sleep in SZA where parameters that might contribute to a better understanding of the pathophysiology of SZA are lacking. Although not statistically significant, our meta-analysis suggested that REMD in SZA might actually be increased like that observed in MDD and higher than that in SCZ. The latter, in fact, has recently been reported not to differ from HC when examined in drug-naive patients (Morra & Barbato, 2025) The hypothesis that REMD is particularly elevated in mood disorders (Kupfer & Heninger, 1972; Sitaram et al., 1982; Kheirkhah et al., 2025) and may therefore be linked to emotional processing (Del Giudice et al., 2025) requires validation in larger samples.

As already mentioned, a recent study provided genetic evidence which strengthened the idea that SZA might be a distinct nosography entity (Kendler et al., 2025), as nonetheless currently proposed by diagnostic manuals such as DSM-V-TR and ICD-11. This evidence might encourage ongoing research (Petit et al., 2022; O’Regan et al., 2025) towards a more comprehensive understanding of the pathophysiology behind SZA including that belonging to the sleep domain. This investigation might also benefit from the use of modern digital sleep scoring devices such as actigraphy or in digital phenotyping (Pieters et al., 2023; Bagautdinova et al., 2023; Tanskanen et al., 2024).

## Data Availability

All data produced in the present work are contained in the manuscript and in the supplementary material attached.

## Abbreviations

BD: Bipolar disorder
DELTAT: Delta sleep time
%DELTA: Delta sleep percentage
EEG: Electroencephalography
HC: Healthy control
MDD: Major depressive disorder
NREM: Non-REM sleep
PRISMA: Preferred reporting items for systematic reviews and meta-analyses
PSG: Polysomnography
RDC: Research diagnostic criteria
REM: Rapid-eye-movement
REMD: Rapid-eye-movement density
REML: Rapid-eye-movement latency
REMT: Rapid-eye-movement time
%REM: Rapid-eye-movement percentage
SCZ: Schizophrenia
SD: Standard deviation
SE: Sleep efficiency
SOL: Sleep onset latency
ST1: Stage 1 sleep time
ST2: Stage 2 sleep time
ST4: Stage 4 sleep time
%ST1: Stage 1 sleep percentage
%ST2: Stage 2 sleep percentage
%ST4: Stage 4 sleep percentage
SWS: Slow-wave sleep
SZA: Schizoaffective disorder
TST: Total sleep time
WASO: Wake after sleep onset
WoS: Web of Science

## References

Abrams, D. J., Rojas, D. C., & Arciniegas, D. B. (2008). Is schizoaffective disorder a distinct categorical diagnosis? A critical review of the literature. Neuropsychiatric Disease and Treatment, 4(6), 1089–1109. 10.2147/ndt.s4120

Alam, A., Chengappa, K. N. R., & Ghinassi, F. (2012). Screening for obstructive sleep apnea among individuals with severe mental illness at a primary care clinic. General Hospital Psychiatry, 34(6), 660–664. 10.1016/j.genhosppsych.2012.06.015

Amann, B. L., Canales-Rodríguez, E. J., Madre, M., Radua, J., Monte, G., Alonso-Lana, S., Landin-Romero, R., Moreno-Alcázar, A., Bonnin, C. M., Sarró, S., Ortiz-Gil, J., Gomar, J. J., Moro, N., Fernandez-Corcuera, P., Goikolea, J. M., Blanch, J., Salvador, R., Vieta, E., McKenna, P. J., & Pomarol-Clotet, E. (2016). Brain structural changes in schizoaffective disorder compared to schizophrenia and bipolar disorder. Acta Psychiatrica Scandinavica, 133(1), 23–33. 10.1111/acps.12440

Angst, J. (2002). Historical aspects of the dichotomy between manic-depressive disorders and schizophrenia. Schizophrenia Research, 57(1), 5–13. 10.1016/s0920-9964(02)00328-6

Angst, J., Felder, W., & Lohmeyer, B. (1980). Course of schizoaffective psychoses: Results of a followup study. Schizophrenia Bulletin, 6(4), 579–585. 10.1093/schbul/6.4.579

Annamalai, A., Palmese, L. B., Chwastiak, L. A., Srihari, V. H., & Tek, C. (2015). High rates of obstructive sleep apnea symptoms among patients with schizophrenia. Psychosomatics, 56(1), 59–66. 10.1016/j.psym.2014.02.009

Bagautdinova, J., Mayeli, A., Wilson, J. D., Donati, F. L., Colacot, R. M., Meyer, N., Fusar-Poli, P., & Ferrarelli, F. (2023). Sleep Abnormalities in Different Clinical Stages of Psychosis: A Systematic Review and Meta-analysis. JAMA Psychiatry, 80(3), 202–210. 10.1001/jamapsychiatry.2022.4599

Benson, K. L., Berger, P. A., & Zarcone, V. P. (1980). Sleep stage measures of psychopathology using Research Diagnostic Criteria. Sleep Research, 9, 161.

Benson, K. L., & Zarcone, V. P. (1985). Testing the REM sleep phasic event intrusion hypothesis of schizophrenia. Psychiatry Research, 15(3), 163–173. 10.1016/0165-1781(85)90073-3

Benson, K. L., Zarcone, V. P., Faull, K. F., Barchas, J. D., & Berger, P. A. (1983). REM sleep eye movement activity and CSF concentrations of 5-hydroxyindoleacetic acid in psychiatric patients. Psychiatry Research, 8(1), 73–78. 10.1016/0165-1781(83)90141-5

Canuso, C. M., Lindenmayer, J.-P., Kosik-Gonzalez, C., Turkoz, I., Carothers, J., Bossie, C. A., & Schooler, N. R. (2010). A randomized, double-blind, placebo-controlled study of 2 dose ranges of paliperidone extended-release in the treatment of subjects with schizoaffective disorder. The Journal of Clinical Psychiatry, 71(5), 587–598. 10.4088/JCP.09m05564yel

Canuso, C. M., Schooler, N., Carothers, J., Turkoz, I., Kosik-Gonzalez, C., Bossie, C. A., Walling, D., & Lindenmayer, J.-P. (2010). Paliperidone extended-release in schizoaffective disorder: A randomized, controlled study comparing a flexible dose with placebo in patients treated with and without antidepressants and/or mood stabilizers. Journal of Clinical Psychopharmacology, 30(5), 487–495. 10.1097/JCP.0b013e3181eeb600

Cardno, A. G., Marshall, E. J., Coid, B., Macdonald, A. M., Ribchester, T. R., Davies, N. J., Venturi, P., Jones, L. A., Lewis, S. W., Sham, P. C., Gottesman, I. I., Farmer, A. E., McGuffin, P., Reveley, A. M., & Murray, R. M. (1999). Heritability estimates for psychotic disorders: The Maudsley twin psychosis series. Archives of General Psychiatry, 56(2), 162–168. 10.1001/archpsyc.56.2.162

Cheniaux, E., Landeira-Fernandez, J., Lessa Telles, L., Lessa, J. L. M., Dias, A., Duncan, T., & Versiani, M. (2008). Does schizoaffective disorder really exist? A systematic review of the studies that compared schizoaffective disorder with schizophrenia or mood disorders. Journal of Affective Disorders, 106(3), 209–217. 10.1016/j.jad.2007.07.009

Cheniaux, E., Landeira-Fernandez, J., & Versiani, M. (2009). The diagnoses of schizophrenia, schizoaffective disorder, bipolar disorder and unipolar depression: Interrater reliability and congruence between DSM-IV and ICD-10. Psychopathology, 42(5), 293–298. 10.1159/000228838

Clayton, P. J. (1982). Schizoaffective disorders. The Journal of Nervous and Mental Disease, 170(11), 646–650. 10.1097/00005053-198211000-00002

Coble, P. A., Kupfer, D. J., & Shaw, D. H. (1981). Distribution of REM latency in depression. Biological Psychiatry, 16(5), 453–466.

Coryell, W., & Zimmerman, M. (1988). The heritability of schizophrenia and schizoaffective disorder. A family study. Archives of General Psychiatry, 45(4), 323–327. 10.1001/archpsyc.1988.01800280033005

Dehelean, L., Romosan, A. M., Bucatos, B. O., Papava, I., Balint, R., Bortun, A. M. C., Toma, M. M., Bungau, S., & Romosan, R. S. (2021). Social and Neurocognitive Deficits in Remitted Patients with Schizophrenia, Schizoaffective and Bipolar Disorder. *Healthcare (Basel*, Switzerland), 9(4), 365. 10.3390/healthcare9040365

del Giudice, R., Stefanelli, R., Donati, F., Nisticò, V., Casetta, C., Castelnovo, A., Zangani, C., Sanguineti, C., Sala, M., Zambrelli, E., Canevini, M. P., & D’Agostino, A. (2026). Elevated gamma activity in left frontotemporal regions preceding sleep signals emotional arousal in Bipolar Disorders: Insights from a high-density EEG investigation. Journal of Affective Disorders, 394, 120517. 10.1016/j.jad.2025.120517

Du, Y., Hao, H., Wang, S., Pearlson, G. D., & Calhoun, V. D. (2020). Identifying commonality and specificity across psychosis sub-groups via classification based on features from dynamic connectivity analysis. NeuroImage. Clinical, 27, 102284. 10.1016/j.nicl.2020.102284

Fallin, M. D., Lasseter, V. K., Avramopoulos, D., Nicodemus, K. K., Wolyniec, P. S., McGrath, J. A., Steel, G., Nestadt, G., Liang, K.-Y., Huganir, R. L., Valle, D., & Pulver, A. E. (2005). Bipolar I disorder and schizophrenia: A 440-single-nucleotide polymorphism screen of 64 candidate genes among Ashkenazi Jewish case-parent trios. American Journal of Human Genetics, 77(6), 918–936. 10.1086/497703

Gama Marques, J., & Ouakinin, S. (2021). Schizophrenia-schizoaffective-bipolar spectra: An epistemological perspective. CNS Spectrums, 26(3), 197–201. 10.1017/S1092852919001408

Gotra, M. Y., Hill, S. K., Gershon, E. S., Tamminga, C. A., Ivleva, E. I., Pearlson, G. D., Keshavan, M. S., Clementz, B. A., McDowell, J. E., Buckley, P. F., Sweeney, J. A., & Keedy, S. K. (2020). Distinguishing patterns of impairment on inhibitory control and general cognitive ability among bipolar with and without psychosis, schizophrenia, and schizoaffective disorder. Schizophrenia Research, 223, 148–157. 10.1016/j.schres.2020.06.033

Guloksuz, S., & van Os, J. (2018). The slow death of the concept of schizophrenia and the painful birth of the psychosis spectrum. Psychological Medicine, 48(2), 229–244. 10.1017/S0033291717001775

Hager, B., Yang, A. C., Brady, R., Meda, S., Clementz, B., Pearlson, G. D., Sweeney, J. A., Tamminga, C., & Keshavan, M. (2017). Neural complexity as a potential translational biomarker for psychosis. Journal of Affective Disorders, 216, 89–99. 10.1016/j.jad.2016.10.016

Harrow, M., Grossman, L. S., Herbener, E. S., & Davies, E. W. (2000). Ten-year outcome: Patients with schizoaffective disorders, schizophrenia, affective disorders and mood-incongruent psychotic symptoms. The British Journal of Psychiatry: The Journal of Mental Science, 177, 421–426. 10.1192/bjp.177.5.421

Higgins JPT, Thomas J, Chandler J, Cumpston M, Li T, Page MJ, Welch VA, editors. (2024). *C*ochrane handbook for systematic reviews of interventions. Cochrane.

Hor, K., & Taylor, M. (2010). Suicide and schizophrenia: A systematic review of rates and risk factors. Journal of Psychopharmacology (Oxford, England), 24(4 Suppl), 81–90. 10.1177/1359786810385490

Ivleva, E. I., Bidesi, A. S., Keshavan, M. S., Pearlson, G. D., Meda, S. A., Dodig, D., Moates, A. F., Lu, H., Francis, A. N., Tandon, N., Schretlen, D. J., Sweeney, J. A., Clementz, B. A., & Tamminga, C. A. (2013). Gray matter volume as an intermediate phenotype for psychosis: Bipolar-Schizophrenia Network on Intermediate Phenotypes (B-SNIP). The American Journal of Psychiatry, 170(11), 1285–1296. 10.1176/appi.ajp.2013.13010126

Ivleva, E. I., Bidesi, A. S., Thomas, B. P., Meda, S. A., Francis, A., Moates, A. F., Witte, B., Keshavan, M. S., & Tamminga, C. A. (2012). Brain gray matter phenotypes across the psychosis dimension. Psychiatry Research, 204(1), 13–24. 10.1016/j.pscychresns.2012.05.001

Jäger, M., Bottlender, R., Strauss, A., & Möller, H.-J. (2004). Fifteen-year follow-up of ICD-10 schizoaffective disorders compared with schizophrenia and affective disorders. Acta Psychiatrica Scandinavica, 109(1), 30–37. 10.1111/j.0001-690x.2004.00208.x

Jäger, M., Haack, S., Becker, T., & Frasch, K. (2011). Schizoaffective disorder—An ongoing challenge for psychiatric nosology. European Psychiatry: The Journal of the Association of European Psychiatrists, 26(3), 159–165. 10.1016/j.eurpsy.2010.03.010

Kantrowitz, J. T., & Citrome, L. (2011). Schizoaffective disorder: A review of current research themes and pharmacological management. CNS Drugs, 25(4), 317–331. 10.2165/11587630-000000000-00000

Kasanin, J. (1994). The acute schizoaffective psychoses. 1933. The American Journal of Psychiatry, 151(6 Suppl), 144–154. 10.1176/ajp.151.6.144

Kaskie, R. E., Graziano, B., & Ferrarelli, F. (2017). Schizophrenia and sleep disorders: Links, risks, and management challenges. Nature and Science of Sleep, 9, 227–239. 10.2147/NSS.S121076

Kendler, K. S., Ohlsson, H., Sundquist, J., & Sundquist, K. (2025). Profiles of Genetic Risks for Psychotic Disorders. JAMA Psychiatry, 82(9), 926–933. 10.1001/jamapsychiatry.2025.1289

Kheirkhah, M., Duncan, W. C., Yuan, Q., Wang, P. R., Jamalabadi, H., Leistritz, L., Walter, M., Goldman, D., Zarate, C. A., & Hejazi, N. S. (2025). REM density predicts rapid antidepressant response to ketamine in individuals with treatment-resistant depression. Neuropsychopharmacology, 50(6), 941–946. 10.1038/s41386-025-02066-7

Kupfer, D. J., Broudy, D., Spiker, D. G., Neil, J. F., & Coble, P. A. (1979). EEG sleep and affective psychoses: I. Schizoaffective disorders. Psychiatry Research, 1(2), 173–178. 10.1016/0165-1781(79)90058-1

Kupfer, D. J., & Foster, F. G. (1975). The sleep of psychotic patients: Does it all look alike? Research Publications - Association for Research in Nervous and Mental Disease, 54, 143–164.

Kupfer, D. J., & Heninger, G. R. (1972). REM activity as a correlate of mood changes throughout the night. Electroencephalographic sleep patterns in a patient with a 48-hour cyclic mood disorder. Archives of General Psychiatry, 27(3), 368–373. 10.1001/archpsyc.1972.01750270072011

Lake, C. R., & Hurwitz, N. (2007). Schizoaffective disorder merges schizophrenia and bipolar disorders as one disease—There is no schizoaffective disorder. Current Opinion in Psychiatry, 20(4), 365–379. 10.1097/YCO.0b013e3281a305ab

Lapensée, M. A. (1992). A review of schizoaffective disorder: I. Current concepts. Canadian Journal of Psychiatry. Revue Canadienne De Psychiatrie, 37(5), 335–346. 10.1177/070674379203700507

Lapierre, Y. D. (1994). Schizophrenia and manic-depression: Separate illnesses or a continuum? Canadian Journal of Psychiatry. Revue Canadienne De Psychiatrie, 39(9 Suppl 2), S59–64.

Laskemoen, J. F., Simonsen, C., Büchmann, C., Barrett, E. A., Bjella, T., Lagerberg, T. V., Vedal, T. J., Andreassen, O. A., Melle, I., & Aas, M. (2019). Sleep disturbances in schizophrenia spectrum and bipolar disorders—A transdiagnostic perspective. Comprehensive Psychiatry, 91, 6–12. 10.1016/j.comppsych.2019.02.006

Laursen, T. M., Labouriau, R., Licht, R. W., Bertelsen, A., Munk-Olsen, T., & Mortensen, P. B. (2005). Family history of psychiatric illness as a risk factor for schizoaffective disorder: A Danish register-based cohort study. Archives of General Psychiatry, 62(8), 841–848. 10.1001/archpsyc.62.8.841

Lynham, A. J., Cleaver, S. L., Jones, I. R., & Walters, J. T. R. (2022). A meta-analysis comparing cognitive function across the mood/psychosis diagnostic spectrum. Psychological Medicine, 52(2), 323–331. 10.1017/S0033291720002020

Madre, M., Canales-Rodríguez, E. J., Ortiz-Gil, J., Murru, A., Torrent, C., Bramon, E., Perez, V., Orth, M., Brambilla, P., Vieta, E., & Amann, B. L. (2016). Neuropsychological and neuroimaging underpinnings of schizoaffective disorder: A systematic review. Acta Psychiatrica Scandinavica, 134(1), 16–30. 10.1111/acps.12564

Malaspina, D., Owen, M. J., Heckers, S., Tandon, R., Bustillo, J., Schultz, S., Barch, D. M., Gaebel, W., Gur, R. E., Tsuang, M., Van Os, J., & Carpenter, W. (2013). Schizoaffective Disorder in the DSM-5. Schizophrenia Research, 150(1), 21–25. 10.1016/j.schres.2013.04.026

Malhi, G. S., Green, M., Fagiolini, A., Peselow, E. D., & Kumari, V. (2008). Schizoaffective disorder: Diagnostic issues and future recommendations. Bipolar Disorders, 10(1 Pt 2), 215–230. 10.1111/j.1399-5618.2007.00564.x

Marneros, A. (2010). The psychotic «continuum». Psychiatrike = Psychiatriki, 21(4), 275–276.

Marneros, A., Deister, A., & Rohde, A. (1990). Psychopathological and social status of patients with affective, schizophrenic and schizoaffective disorders after long-term course. Acta Psychiatrica Scandinavica, 82(5), 352–358. 10.1111/j.1600-0447.1990.tb01400.x

Marneros, A., Röttig, S., Wenzel, A., Blöink, R., & Brieger, P. (2004). Affective and schizoaffective mixed states. European Archives of Psychiatry and Clinical Neuroscience, 254(2), 76–81. 10.1007/s00406-004-0462-9

Mazza, M. G., Capellazzi, M., Tagliabue, I., Lucchi, S., Rossetti, A., & Clerici, M. (2019). Neutrophil-lymphocyte, monocyte-lymphocyte and platelet-lymphocyte ratio in schizoaffective disorder compared to schizophrenia. General Hospital Psychiatry, 61, 86–87. 10.1016/j.genhosppsych.2019.06.013

Meltzer, H. Y. (1984). Schizoaffective disorder: Is the news of its nonexistence premature? Editor’s introduction. Schizophrenia Bulletin, 10(1), 11–13. 10.1093/schbul/10.1.11

Miller, B. J., Parker, C. B., Rapaport, M. H., Buckley, P. F., & McCall, W. V. (2019). Insomnia and suicidal ideation in nonaffective psychosis. Sleep, 42(2), zsy215. 10.1093/sleep/zsy215

Miller, J. N., & Black, D. W. (2019). Schizoaffective disorder: A review. Annals of Clinical Psychiatry: Official Journal of the American Academy of Clinical Psychiatrists, 31(1), 47–53.

Morra, D., & Barbato, G. (2025). REM sleep in schizophrenia: a systematic review and meta-analysis. Sleep medicine reviews, 83, 102134. 10.1016/j.smrv.2025.102134

Muñoz-Negro, J. E., Cuadrado, L., & Cervilla, J. A. (2019). Current Evidences on Psychopharmacology of Schizoaffective Disorder. Actas Espanolas De Psiquiatria, 47(5), 190–201.

O’Regan, D., Poole, N., Otaiku, A., Jackson, M. L., & Rosenzweig, I. (2025). Sleep Neurophysiology and Psychiatric Disorders: A Transdiagnostic Framework for Mechanistic and Therapeutic Insight. Current Treatment Options in Neurology, 27(1), 36. 10.1007/s11940-025-00847-5

Palmese, L. B., DeGeorge, P. C., Ratliff, J. C., Srihari, V. H., Wexler, B. E., Krystal, A. D., & Tek, C. (2011). Insomnia is frequent in schizophrenia and associated with night eating and obesity. Schizophrenia Research, 133(1–3), 238–243. 10.1016/j.schres.2011.07.030

Paul, T., Javed, S., Karam, A., Loh, H., & Ferrer, G. F. (2021). A Misdiagnosed Case of Schizoaffective Disorder With Bipolar Manifestations. Cureus, 13(7), e16686. 10.7759/cureus.16686

Pavlichenko, A., Petrova, N., & Stolyarov, A. (2024). The Modern Concept of Schizoaffective Disorder: A Narrative Review. Consortium Psychiatricum, 5(3), 42–55. 10.17816/CP15513

Peterson, D. L., Webb, C. A., Keeley, J. W., Gaebel, W., Zielasek, J., Rebello, T. J., Robles, R., Matsumoto, C., Kogan, C. S., Kulygina, M., Farooq, S., Green, M. F., Falkai, P., Hasan, A., Galderisi, S., Larach, V., Krasnov, V., & Reed, G. M. (2019). The reliability and clinical utility of ICD-11 schizoaffective disorder: A field trial. Schizophrenia Research, 208, 235–241. 10.1016/j.schres.2019.02.011

Petit, J.-M., Strippoli, M.-P. F., Stephan, A., Ranjbar, S., Haba-Rubio, J., Solelhac, G., Heinzer, R., Preisig, M., Siclari, F., & Do, K. Q. (2022). Sleep spindles in people with schizophrenia, schizoaffective disorders or bipolar disorders: A pilot study in a general population-based cohort. BMC Psychiatry, 22(1), 758. 10.1186/s12888-022-04423-y

Pieters, L. E., Deenik, J., de Vet, S., Delespaul, P., & van Harten, P. N. (2023). Combining actigraphy and experience sampling to assess physical activity and sleep in patients with psychosis: A feasibility study. Frontiers in Psychiatry, 14, 1107812. 10.3389/fpsyt.2023.1107812

Pinna, F., Sanna, L., Perra, V., Pisu Randaccio, R., Diana, E., Carpiniello, B., & Cagliari Recovery Study Group. (2014). Long-term outcome of schizoaffective disorder. Are there any differences with respect to schizophrenia? Rivista Di Psichiatria, 49(1), 41–49. 10.1708/1407.15624

Poulin, J., Chouinard, S., Pampoulova, T., Lecomte, Y., Stip, E., & Godbout, R. (2010). Sleep habits in middle-aged, non-hospitalized men and women with schizophrenia: A comparison with healthy controls. Psychiatry Research, 179(3), 274–278. 10.1016/j.psychres.2009.08.009

Radonić, E., Rados, M., Kalember, P., Bajs-Janović, M., Folnegović-Smalc, V., & Henigsberg, N. (2011). Comparison of hippocampal volumes in schizophrenia, schizoaffective and bipolar disorder. Collegium Antropologicum, 35 *Suppl 1*, 249–252.

Reich, L., Weiss, B. L., Coble, P., McPartland, R., & Kupfer, D. J. (1975). Sleep disturbance in schizophrenia. A revisit. Archives of General Psychiatry, 32(1), 51–55. 10.1001/archpsyc.1975.01760190053006

Rieder, R. O., Mann, L. S., Weinberger, D. R., van Kammen, D. P., & Post, R. M. (1983). Computed tomographic scans in patients with schizophrenia, schizoaffective, and bipolar affective disorder. Archives of General Psychiatry, 40(7), 735–739. 10.1001/archpsyc.1983.01790060033004

Rink, L., Pagel, T., Franklin, J., & Baethge, C. (2016). Characteristics and heterogeneity of schizoaffective disorder compared with unipolar depression and schizophrenia—A systematic literature review and meta-analysis. Journal of Affective Disorders, 191, 8–14. 10.1016/j.jad.2015.10.045

Rosenthal, N. E., Rosenthal, L. N., Stallone, F., Dunner, D. L., & Fieve, R. R. (1980). Toward the validation of RDC schizoaffective disorder. Archives of General Psychiatry, 37(7), 804–810. 10.1001/archpsyc.1980.01780200082009

Santelmann, H., Franklin, J., Bußhoff, J., & Baethge, C. (2015). Test-retest reliability of schizoaffective disorder compared with schizophrenia, bipolar disorder, and unipolar depression—A systematic review and meta-analysis. Bipolar Disorders, 17(7), 753–768. 10.1111/bdi.12340

Scully, P. J., Owens, J. M., Kinsella, A., & Waddington, J. L. (2004). Schizophrenia, schizoaffective and bipolar disorder within an epidemiologically complete, homogeneous population in rural Ireland: Small area variation in rate. Schizophrenia Research, 67(2–3), 143–155. 10.1016/S0920-9964(03)00194-4

Sitaram, N., Nurnberger, J. I., Gershon, E. S., & Gillin, J. C. (1982). Cholinergic regulation of mood and REM sleep: Potential model and marker of vulnerability to affective disorder. The American Journal of Psychiatry, 139(5), 571–576. 10.1176/ajp.139.5.571

Sovner, R. D., & McHugh, P. R. (1976). Bipolar course in schizo-affective illness. Biological Psychiatry, 11(2), 195–204.

Tanskanen, T. E., Wegelius, A., Härkönen, T., Gummerus, E.-M., Stenberg, J.-H., Selinheimo, S. I. K., Alakuijala, A., Tenhunen, M., Paajanen, T., Järnefelt, H., Kajaste, S., Blom, K., Kieseppä, T., Tuisku, K., & Paunio, T. (2024). Cognitive behavioural therapy for insomnia (CBT-I) in schizophrenia and schizoaffective disorder: Protocol for a randomised controlled trial. BMJ Open, 14(6), e076129. 10.1136/bmjopen-2023-076129

Tsuang, M. T., & Simpson, J. C. (1984). Schizoaffective disorder: Concept and reality. Schizophrenia Bulletin, 10(1), 14–25. 10.1093/schbul/10.1.14

Wilson, J. E., Nian, H., & Heckers, S. (2014). The schizoaffective disorder diagnosis: A conundrum in the clinical setting. European Archives of Psychiatry and Clinical Neuroscience, 264(1), 29–34. 10.1007/s00406-013-0410-7

Wy, T. J. P., & Saadabadi, A. (2025). Schizoaffective Disorder. In StatPearls. StatPearls Publishing. http://www.ncbi.nlm.nih.gov/books/NBK541012/

Zarcone, V. P., & Benson, K. L. (1995). Middle ear muscle activity (MEMA) in schizophrenia using a noninvasive technique. Sleep, 18(4), 266–271. 10.1093/sleep/18.4.266

Zarcone, V. P., Benson, K. L., & Berger, P. A. (1987). Abnormal rapid eye movement latencies in schizophrenia. Archives of General Psychiatry, 44(1), 45–48. 10.1001/archpsyc.1987.01800130047007

