## Supplementary Material S1 for "Polysomnographic parameters in schizoaffective disorder: a systematic review and meta-analysis"

| Study | N. (SZA)/N. (HC) | Age (SZA/HC) | Males % (SZA/HC) | Diagnosis (SZA/HC) | Treatment |
|---|---|---|---|---|---|
| **Benson et al., 1980** | 9/19 | ?/? | ?/? | Schizoaffective disorder/Healthy | Drug-free (≥2 weeks) |
| **Benson et al., 1985** | 8/13 | 33/29.7 | 100/100 | Schizoaffective disorder/Healthy | Drug-free (≥2 weeks) |
| **Benson et al., 1985** | 9/19 | 33.4/30.2 | 100/100 | Schizoaffective disorder/Healthy | Drug-free (≥2 weeks) |
| **Zarcone et al., 1987** | 8/18 | 33.8/30.3 | 100/100 | Schizoaffective disorder/Healthy | Drug-free (≥2 weeks) |
| **Zarcone et al., 1995** | 8/10 | 34.5/32.3 | 100/100 | Schizoaffective disorder/Healthy | Drug-free (2 days) |

| Study | N. SZA | N. HC | REMT (SZA) | SD | REMT (HC) | SD | %REM (SZA) | SD | %REM (HC) | SD | REML (SZA) | SD | REML (HC) | SD | REMD (SZA) | SD | REMD (HC) | SD |
|---|---|---|---|---|---|---|---|---|---|---|---|---|---|---|---|---|---|---|
| **Benson et al., 1980** | 9 | 19 | | | | | 23.66 | | 22.38 | | 75.98 | | 96.66 | | | | | |
| **Benson et al., 1985** | 8 | 13 | 68.65 | 27.48 | 95.30 | 28.20 | 18.35 | 7.65 | 22.40 | 7.45 | 74.20 | 41.45 | 89.90 | 40.50 | | | | |
| **Benson et al., 1985** | 9 | 19 | 92.30 | 38.80 | 98.90 | 21.00 | 22.70 | 7.85 | 22.70 | 5.40 | 73.80 | 24.65 | 82.20 | 32.90 | | | | |
| **Zarcone et al., 1987** | 8 | 18 | 91.16 | 20.19 | 101.62 | 13.17 | 22.98 | 4.20 | 22.57 | 3.16 | 53.46 | 14.90 | 88.60 | 42.79 | | | | |
| **Zarcone et al., 1995** | 8 | 10 | 65.10 | | 86.40 | | 20.20 | 4.00 | 20.80 | 4.50 | 52.20 | 32.80 | 65.30 | 16.40 | | | | |

| Study | N. (SZA)/N. (MDD) | Age (SZA/MDD) | Males % (SZA/MDD) | Diagnosis (SZA/MDD) | Treatment |
|---|---|---|---|---|---|
| **Kupfer & Foster, 1975** | 6/9 | 38.8/58.2 | 50/? | Schizoaffective disorder/Psychotic depression | Drug-free (≥2 weeks) |
| **Kupfer & Foster, 1975** | 6/25 | 38.8/41.1 | 50/? | Schizoaffective disorder/MDD | Drug-free (≥2 weeks) |
| **Kupfer et al., 1979** | 12/29 | 44.4/48.1 | 42/31 | Schizoaffective disorder/Psychotic depression | Drug-free (≥2 weeks) |
| **Benson et al., 1980** | 9/6 | ?/? | ?/? | Schizoaffective disorder/MDD | Drug-free (≥2 weeks) |
| **Coble et al., 1981** | 5/17 | 34.8/40.1 | 60/35 | Schizoaffective disorder/<br>Psychotic and nonpsychotic depression | Drug-free (2 weeks) |
| **Benson et al., 1983** | 2/5 | ?/? | 100/100 | Schizoaffective disorder/MDD | Drug-free (≥2 weeks) |
| **Benson et al., 1985** | 8/10 | 33/40.3 | 100/100 | Schizoaffective disorder/MDD | Drug-free (≥2 weeks) |
| **Benson et al., 1985** | 9/7 | 33.4/37.7 | 100/100 | Schizoaffective disorder/MDD | Drug-free (≥2 weeks) |
| **Zarcone et al., 1987** | 8/12 | 33.8/41.33 | 100/100 | Schizoaffective disorder/MDD | Drug-free (≥2 weeks) |
| **Zarcone et al., 1995** | 8/11 | 34.5/37.2 | 100/100 | Schizoaffective disorder/MDD | Drug-free (2 days) |

| Study | N. SZA | N. MDD | REMT (SZA) | SD | REMT (MDD) | SD | %REM (SZA) | SD | %REM (MDD) | SD | REML (SZA) | SD | REML (MDD) | SD | REMD (SZA) | SD | REMD (MDD) | SD |
|---|---|---|---|---|---|---|---|---|---|---|---|---|---|---|---|---|---|---|
| **Kupfer & Foster, 1975** | 6 | 9 | 81.00 | 10.50 | 51.30 | 7.00 | 23.20 | 1.90 | 19.60 | 2.40 | 31.70 | 5.70 | 36.00 | 7.20 | 2.33 | | 2.35 | |
| **Kupfer & Foster, 1975** | 6 | 25 | | | | | | | | | 31.70 | 5.70 | 40.91 | 6.05 | | | | |
| **Kupfer et al., 1979** | 12 | 29 | 47.22 | | 57.24 | | 17.70 | 6.60 | 21.10 | 5.30 | 23.41 | 27.90 | 41.20 | 33.40 | 1.34 | 0.71 | 1.55 | 0.95 |
| **Benson et al., 1980** | 9 | 6 | | | | | 23.66 | | 30.23 | | 75.98 | | 49.33 | | | | | |
| **Coble et al., 1981** | 5 | 17 | | | | | | | | | 49.87 | | 46.04 | | | | | |
| **Benson et al., 1983** | 2 | 5 | 90.20 | | 106.66 | | 22.96 | | 24.97 | | 68.25 | | 62.28 | | 2.62 | | 2.61 | |
| **Benson et al., 1985** | 8 | 10 | 68.65 | 27.48 | 89.40 | 53.70 | 18.35 | 7.65 | 19.90 | 11.95 | 74.20 | 41.45 | 63.60 | 17.80 | | | | |
| **Benson et al., 1985** | 9 | 7 | 92.30 | 38.80 | 109.60 | 52.70 | 22.70 | 7.85 | 26.30 | 14.30 | 73.80 | 24.65 | 44.30 | 65.70 | | | | |
| **Zarcone et al., 1987** | 8 | 12 | 91.16 | 20.19 | 105.12 | 32.78 | 22.98 | 4.20 | 25.28 | 7.10 | 53.46 | 14.90 | 48.77 | 24.46 | | | | |
| **Zarcone et al., 1995** | 8 | 11 | 65.10 | | 83.25 | | 20.20 | 4.00 | 21.80 | 3.90 | 52.20 | 32.80 | 45.50 | 22.10 | | | | |

| Study | N. (SZA)/N. (SCZ) | Age (SZA/SCZ) | Males % (SZA/SCZ) | Diagnosis (SZA/SCZ) | Treatment |
|---|---|---|---|---|---|
| **Reich et al., 1975** | 3/14 | 38.8/20.8 | 50/57 | Schizoaffective disorder/Schizophrenia (acute) | Drug-free (≥2 weeks) |
| **Reich et al., 1975** | 3/9 | 38.8/19 | 50/11 | Schizoaffective disorder/Schizophrenia (latent) | Drug-free (≥2 weeks) |
| **Benson et al., 1980** | 9/9 | ?/? | ?/? | Schizoaffective disorder/Schizophrenia | Drug-free (≥2 weeks) |
| **Benson et al., 1983** | 2/7 | ?/? | 100/100 | Schizoaffective disorder/Schizophrenia | Drug-free (≥2 weeks) |
| **Benson et al., 1985** | 8/11 | 33/27 | 100/100 | Schizoaffective disorder/Schizophrenia | Drug-free (≥2 weeks) |
| **Benson et al., 1985** | 9/8 | 33.4/26.6 | 100/100 | Schizoaffective disorder/Schizophrenia | Drug-free (≥2 weeks) |
| **Zarcone et al., 1987** | 8/12 | 33.8/26.2 | 100/100 | Schizoaffective disorder/Schizophrenia | Drug-free (≥2 weeks) |
| **Zarcone et al., 1995** | 8/18 | 34.5/33.1 | 100/100 | Schizoaffective disorder/Schizophrenia | Drug-free (2 days) |

| Study | N. SZA | N. SCZ | REMT (SZA) | SD | REMT (SCZ) | SD | %REM (SZA) | SD | %REM (SCZ) | SD | REML (SZA) | SD | REML (SCZ) | SD | REMD (SZA) | SD | REMD (SCZ) | SD |
|---|---|---|---|---|---|---|---|---|---|---|---|---|---|---|---|---|---|---|
| **Reich et al., 1975** | 3 | 14 | 81 | 10.5 | 54 | 5.6 | 23.20 | 1.90 | 17.40 | 1.60 | 31.70 | 5.70 | 97.90 | 11.00 | 2.20 | 0.20 | 1.60 | 0.10 |
| **Reich et al., 1975** | 3 | 9 | 81 | 10.5 | 74.8 | 5.6 | 23.20 | 1.90 | 20.00 | 1.40 | 31.70 | 5.70 | 81.00 | 12.00 | 2.20 | 0.20 | 1.50 | 0.10 |
| **Benson et al., 1980** | 9 | 9 | | | | | 23.66 | | 25.44 | | 75.98 | | 66.20 | | | | | |
| **Benson et al., 1983** | 2 | 7 | 90.20 | | 100.37 | | 22.96 | | 26.01 | | 68.25 | | 50.89 | | 2.62 | | 2.66 | |
| **Benson et al., 1985** | 8 | 11 | 68.65 | 27.48 | 98.00 | 40.30 | 18.35 | 7.65 | 25.85 | 5.70 | 74.20 | 41.45 | 62.90 | 41.00 | | | | |
| **Benson et al., 1985** | 9 | 8 | 92.30 | 38.80 | 101.70 | 19.45 | 22.70 | 7.85 | 24.45 | 4.78 | 73.80 | 24.65 | 75.60 | 32.83 | | | | |
| **Zarcone et al., 1987** | 8 | 12 | 91.16 | 20.19 | 99.62 | 22.73 | 22.99 | 4.20 | 4.04 | 0.26 | 53.46 | 14.90 | 49.68 | 15.57 | | | | |
| **Zarcone et al., 1995** | 8 | 18 | 65.10 | | 75.07 | | 20.20 | 4.00 | 20.80 | 3.70 | 52.20 | 32.80 | 53.60 | 14.00 | | | | |

| Study | N. SZA | REMT (SZA) | SD | N. HC | REMT (HC) | SD |
|---|---|---|---|---|---|---|
| Benson et al., 1985 | 8 | 68.65 | 27.48 | 13 | 95.30 | 28.20 |
| Benson et al., 1985 | 9 | 92.30 | 38.80 | 19 | 98.90 | 21.00 |
| Zarcone et al., 1987 | 8 | 91.16 | 20.19 | 18 | 101.62 | 13.17 |
| Zarcone et al., 1995 | 8 | 65.10 | 29.22 | 10 | 86.40 | 20.05 |
|  | 33 | 79.70 | 29.22 | 60 | 96.85 | 20.05 |

| Study | N. SZA | %REM (SZA) | SD | N. HC | %REM (HC) | SD |
|---|---|---|---|---|---|---|
| Benson et al., 1980 | 9 | 23.66 | 5.98 | 19 | 22.38 | 5.02 |
| Benson et al., 1985 | 8 | 18.35 | 7.65 | 13 | 22.40 | 7.45 |
| Benson et al., 1985 | 9 | 22.70 | 7.85 | 19 | 22.70 | 5.40 |
| Zarcone et al., 1987 | 8 | 22.98 | 4.20 | 18 | 22.57 | 3.16 |
| Zarcone et al., 1995 | 8 | 20.20 | 4.00 | 10 | 20.80 | 4.50 |
|  | 42 | 21.65 | 5.98 | 79 | 22.30 | 5.02 |

| Study | N. SZA | REML (SZA) | SD | N. HC | REML (HC) | SD |
|---|---|---|---|---|---|---|
| Benson et al., 1980 | 9 | 75.98 | 28.33 | 19 | 96.66 | 34.76 |
| Benson et al., 1985 | 8 | 74.20 | 41.45 | 13 | 89.90 | 40.50 |
| Benson et al., 1985 | 9 | 73.80 | 24.65 | 19 | 82.20 | 32.90 |
| Zarcone et al., 1987 | 8 | 53.46 | 14.90 | 18 | 88.60 | 42.79 |
| Zarcone et al., 1995 | 8 | 52.20 | 32.80 | 10 | 65.30 | 16.40 |
|  | 42 | 66.35 | 28.33 | 60 | 86.26 | 34.76 |

| Study | N. SZA | REMT (SZA) | SD | N. MDD | REMT (MDD) | SD |
|---|---|---|---|---|---|---|
| Kupfer & Foster, 1975 | 6 | 81.00 | 10.50 | 9 | 51.30 | 7.00 |
| Kupfer et al., 1979 | 12 | 47.22 | 25.60 | 29 | 57.24 | 35.85 |
| Benson et al., 1983 | 2 | 90.20 | 25.60 | 5 | 106.66 | 35.85 |
| Benson et al., 1985 | 8 | 68.65 | 27.48 | 10 | 89.40 | 53.70 |
| Benson et al., 1985 | 9 | 92.30 | 38.80 | 7 | 109.60 | 52.70 |
| Zarcone et al., 1987 | 8 | 91.16 | 20.19 | 12 | 105.12 | 32.78 |
| Zarcone et al., 1995 | 8 | 65.10 | 25.60 | 11 | 83.25 | 35.85 |
|  | 53 | 72.89 | 25.60 | 83 | 78.23 | 35.85 |

| Study | N. SZA | %REM (SZA) | SD | N. MDD | %REM (MDD) | SD |
|---|---|---|---|---|---|---|
| Kupfer & Foster, 1975 | 6 | 23.20 | 1.90 | 9 | 19.60 | 2.40 |
| Kupfer et al., 1979 | 12 | 17.70 | 6.60 | 29 | 21.10 | 5.30 |
| Benson et al., 1980 | 9 | 23.66 | 5.65 | 6 | 30.23 | 6.71 |
| Benson et al., 1983 | 2 | 22.96 | 5.65 | 5 | 24.97 | 6.71 |
| Benson et al., 1985 | 8 | 18.35 | 7.65 | 10 | 19.90 | 11.95 |
| Benson et al., 1985 | 9 | 22.70 | 7.85 | 7 | 26.30 | 14.30 |
| Zarcone et al., 1987 | 8 | 22.98 | 4.20 | 12 | 25.28 | 7.10 |
| Zarcone et al., 1995 | 8 | 20.20 | 4.00 | 11 | 21.80 | 3.90 |
|  | 62 | 21.08 | 5.65 | 89 | 22.71 | 6.71 |

| Study | N. SZA | REML (SZA) | SD | N. MDD | REML (MDD) | SD |
|---|---|---|---|---|---|---|
| Kupfer & Foster, 1975 | 3 | 31.70 | 5.70 | 9 | 36.00 | 7.20 |
| Kupfer & Foster, 1975 | 3 | 31.70 | 5.70 | 25 | 40.91 | 6.05 |
| Kupfer et al., 1979 | 12 | 23.41 | 27.90 | 29 | 41.20 | 33.40 |
| Benson et al., 1980 | 9 | 75.98 | 25.57 | 6 | 49.33 | 22.90 |
| Coble et al., 1981 | 5 | 49.87 | 25.57 | 17 | 46.04 | 22.90 |
| Benson et al., 1983 | 2 | 68.25 | 25.57 | 5 | 62.28 | 22.90 |
| Benson et al., 1985 | 8 | 74.20 | 41.45 | 10 | 63.60 | 17.80 |
| Benson et al., 1985 | 9 | 73.80 | 24.65 | 7 | 44.30 | 65.70 |
| Zarcone et al., 1987 | 8 | 53.46 | 14.90 | 12 | 48.77 | 24.46 |
| Zarcone et al., 1995 | 8 | 52.20 | 32.80 | 11 | 45.50 | 22.10 |
|  | 67 | 54.39 | 25.57 | 131 | 45.52 | 22.90 |

| Study | N. SZA | REMD (SZA) | SD | N. MDD | REMD (MDD) | SD |
|---|---|---|---|---|---|---|
| Kupfer & Foster, 1975 | 6 | 2.33 | 0.71 | 9 | 2.35 | 0.95 |
| Kupfer et al., 1979 | 12 | 1.34 | 0.71 | 29 | 1.55 | 0.95 |
| Benson et al., 1983 | 2 | 2.62 | 0.71 | 5 | 2.61 | 0.95 |
|  | 20 | 1.77 | 0.71 | 43 | 1.84 | 0.95 |

| Study | N. SZA | REMT (SZA) | SD | N. SCZ | REMT (SCZ) | SD |
|---|---|---|---|---|---|---|
| Benson et al., 1983 | 2 | 90.20 | 25.60 | 7 | 100.37 | 18.53 |
| Benson et al., 1985 | 8 | 68.65 | 27.48 | 11 | 98.00 | 40.30 |
| Benson et al., 1985 | 9 | 92.30 | 38.80 | 8 | 101.70 | 19.45 |
| Zarcone et al., 1987 | 8 | 91.16 | 20.19 | 12 | 99.62 | 22.73 |
| Zarcone et al., 1995 | 8 | 65.10 | 25.60 | 18 | 75.07 | 18.53 |
| Reich et al., 1975 | 3 | 81.00 | 10.50 | 14 | 54.00 | 5.60 |
| Reich et al., 1975 | 3 | 81.00 | 10.50 | 9 | 74.80 | 5.60 |
|  | 41 | 80.40 | 25.60 | 79 | 83.17 | 18.53 |

| Study | N. SZA | %REM (SZA) | SD | N. SCZ | %REM (SCZ) | SD |
|---|---|---|---|---|---|---|
| Benson et al., 1980 | 9 | 23.66 | 5.36 | 9 | 25.44 | 2.86 |
| Benson et al., 1983 | 2 | 22.96 | 5.36 | 7 | 26.01 | 2.86 |
| Benson et al., 1985 | 8 | 18.35 | 7.65 | 11 | 25.85 | 5.70 |
| Benson et al., 1985 | 9 | 22.70 | 7.85 | 8 | 24.45 | 4.78 |
| Zarcone et al., 1987 | 8 | 22.99 | 4.20 | 12 | 4.04 | 0.26 |
| Zarcone et al., 1995 | 8 | 20.20 | 4.00 | 18 | 20.80 | 3.70 |
| Reich et al., 1975 | 3 | 23.20 | 1.90 | 14 | 17.40 | 1.60 |
| Reich et al., 1975 | 3 | 23.20 | 1.90 | 9 | 20.00 | 1.40 |
|  | 50 | 21.89 | 5.36 | 88 | 19.74 | 2.86 |

| Study | N. SZA | REML (SZA) | SD | N. SCZ | REML (SCZ) | SD |
|---|---|---|---|---|---|---|
| Benson et al., 1980 | 9 | 75.98 | 24.85 | 9 | 66.20 | 19.65 |
| Benson et al., 1983 | 2 | 68.25 | 24.85 | 7 | 50.89 | 19.65 |
| Benson et al., 1985 | 8 | 74.20 | 41.45 | 11 | 62.90 | 41.00 |
| Benson et al., 1985 | 9 | 73.80 | 24.65 | 8 | 75.60 | 32.83 |
| Zarcone et al., 1987 | 8 | 53.46 | 14.90 | 12 | 49.68 | 15.57 |
| Zarcone et al., 1995 | 8 | 52.20 | 32.80 | 18 | 53.60 | 14.00 |
| Reich et al., 1975 | 3 | 31.70 | 5.70 | 14 | 97.90 | 11.00 |
| Reich et al., 1975 | 3 | 31.70 | 5.70 | 9 | 81.00 | 12.00 |
|  | 50 | 62.27 | 24.85 | 88 | 67.15 | 19.65 |

| Study | N. SZA | REMD (SZA) | SD | N. SCZ | REMD (SCZ) | SD |
|---|---|---|---|---|---|---|
| Benson et al., 1983 | 2 | 2.62 | 0.20 | 7 | 2.66 | 0.10 |
| Reich et al., 1975 | 3 | 2.20 | 0.20 | 14 | 1.60 | 0.10 |
| Reich et al., 1975 | 3 | 2.20 | 0.20 | 9 | 1.50 | 0.10 |
|  | 8 | 2.31 | 0.20 | 30 | 1.82 | 0.10 |

**Drug-free SZA vs HC**

**Drug-free SZA vs MDD**

**Drug-free SZA vs SCZ**

**Calculated from original data**

**SD not available in original data (averaged from similar available data)**

| Study | N. (SZA)/N. (HC) | Age (SZA/HC) | Males % (SZA/HC) | Diagnosis (SZA/HC) | Treatment |
|---|---|---|---|---|---|
| **Benson et al., 1980** | 9/19 | ?/? | ?/? | Schizoaffective disorder/Healthy | Drug-free (≥2 weeks) |
| **Benson et al., 1985** | 8/13 | 33/29.7 | 100/100 | Schizoaffective disorder/Healthy | Drug-free (≥2 weeks) |
| **Benson et al., 1985** | 9/19 | 33.4/30.2 | 100/100 | Schizoaffective disorder/Healthy | Drug-free (≥2 weeks) |
| **Zarcone et al., 1987** | 8/18 | 33.8/30.3 | 100/100 | Schizoaffective disorder/Healthy | Drug-free (≥2 weeks) |
| **Zarcone et al., 1995** | 8/10 | 34.5/32.3 | 100/100 | Schizoaffective disorder/Healthy | Drug-free (2 days) |

| Study | N. SZA | N. HC | ST1 (SZA) | SD | ST1 (HC) | SD | %ST1 (SZA) | SD | %ST1 (HC) | SD | ST2 (SZA) | SD | ST2 (HC) | SD | %ST2 (SZA) | SD | %ST2 (HC) | SD | ST3 (SZA) | SD | ST3 (HC) | SD | %ST3 (SZA) | SD | %ST3 (HC) | SD | ST4 (SZA) | SD | ST4 (HC) | SD | %ST4 (SZA) | SD | %ST4 (HC) | SD | DELTAT (SZA) | SD | DELTAT (HC) | SD | %DELTA (SZA) | SD | %DELTA (HC) | SD |
|---|---|---|---|---|---|---|---|---|---|---|---|---|---|---|---|---|---|---|---|---|---|---|---|---|---|---|---|---|---|---|---|---|---|---|---|---|---|---|---|---|---|---|
| **Benson et al., 1980** | 9 | 19 | | | | | | | | | 206.98 | | 224.25 | | | | | | 30.74 | | 35.97 | | | | | | 12.62 | | 49.34 | | 3.17 | | 11.02 | | 43.36 | | 85.31 | | | | | |
| **Benson et al., 1985** | 8 | 13 | | | | | | | | | | | | | | | | | | | | | | | | | 3.25 | 14.05 | 46.50 | 44.30 | 0.75 | 4.73 | 10.00 | 10.50 | | | | | | | | |
| **Benson et al., 1985** | 9 | 19 | | | | | | | | | | | | | | | | | | | | | | | | | 6.60 | 18.05 | 46.10 | 32.00 | 1.60 | 4.55 | 10.40 | 6.80 | | | | | | | | |
| **Zarcone et al., 1987** | 8 | 18 | | | | | | | | | | | | | | | | | | | | | | | | | | | | | | | | | 42.94 | 32.25 | 83.18 | 30.90 | | | | |
| **Zarcone et al., 1995** | 8 | 10 | 50.26 | | 80.17 | | 15.6 | 5.6 | 19.30 | 6.40 | 163.68 | | 209.78 | | 50.80 | 6.20 | 50.50 | 7.30 | | | | | | | | | | | | | | | | | 43.17 | | 38.63 | | 13.40 | 8.10 | 9.30 | 6.20 |

| Study | N. (SZA)/N. (MDD) | Age (SZA/MDD) | Males % (SZA/MDD) | Diagnosis (SZA/MDD) | Treatment |
|---|---|---|---|---|---|
| **Kupfer & Foster, 1975** | 6/9 | 38.8/58.2 | 50/? | Schizoaffective disorder/Psychotic depression | Drug-free (≥2 weeks) |
| **Kupfer & Foster, 1975** | 6/25 | 38.8/41.1 | 50/? | Schizoaffective disorder/MDD | Drug-free (≥2 weeks) |
| **Kupfer et al., 1979** | 12/29 | 44.4/48.1 | 42/31 | Schizoaffective disorder/Psychotic depression | Drug-free (≥2 weeks) |
| **Benson et al., 1980** | 9/6 | ?/? | ?/? | Schizoaffective disorder/ | Drug-free (≥2 weeks) |
| **Benson et al., 1985** | 8/10 | 33/40.3 | 100/100 | Schizoaffective disorder/MDD | Drug-free (≥2 weeks) |
| **Benson et al., 1985** | 9/7 | 33.4/37.7 | 100/100 | Schizoaffective disorder/MDD | Drug-free (≥2 weeks) |
| **Zarcone et al., 1987** | 8/12 | 33.8/41.33 | 100/100 | Schizoaffective disorder/MDD | Drug-free (≥2 weeks) |
| **Zarcone et al., 1995** | 8/11 | 34.5/37.2 | 100/100 | Schizoaffective disorder/MDD | Drug-free (2 days) |

| Study | N. SZA | N. MDD | ST1 (SZA) | SD | ST1 (MDD) | SD | %ST1 (SZA) | SD | %ST1 (MDD) | SD | ST2 (SZA) | SD | ST2 (MDD) | SD | %ST2 (SZA) | SD | %ST2 (MDD) | SD | ST3 (SZA) | SD | ST3 (MDD) | SD | %ST3 (SZA) | SD | %ST3 (MDD) | SD | ST4 (SZA) | SD | ST4 (MDD) | SD | %ST4 (SZA) | SD | %ST4 (MDD) | SD | DELTAT (SZA) | SD | DELTAT (MDD) | SD | %DELTA (SZA) | SD | %DELTA (MDD) | SD |
|---|---|---|---|---|---|---|---|---|---|---|---|---|---|---|---|---|---|---|---|---|---|---|---|---|---|---|---|---|---|---|---|---|---|---|---|---|---|---|---|---|---|---|
| **Kupfer & Foster, 1975** | 6 | 9 | | | | | | | | | | | | | | | | | | | | | | | | | | | | | | | | | 18.20 | 4.50 | 4.40 | 2.20 | 4.80 | 1.20 | 1.60 | 0.85 |
| **Kupfer & Foster, 1975** | 6 | 25 | | | | | | | | | | | | | | | | | | | | | | | | | | | | | | | | | 18.20 | 4.50 | 21.00 | 5.70 | | | | |
| **Kupfer et al., 1979** | 12 | 29 | 24.55 | | 33.37 | | 9.20 | 5.30 | 12.30 | 6.40 | 184.90 | | 172.00 | | 69.30 | 8.60 | 63.40 | 10.10 | | | | | | | | | | | | | | | | | 1.07 | | 0.81 | | 0.40 | 1.10 | 0.30 | 1.70 |
| **Benson et al., 1980** | 9 | 6 | | | | | | | | | 206.98 | | 172.88 | | | | | | 30.74 | | 45.11 | | | | | | 12.62 | | 19.92 | | 3.17 | | 4.72 | | 43.36 | | 65.03 | | | | | |
| **Benson et al., 1985** | 8 | 10 | | | | | | | | | | | | | | | | | | | | | | | | | 3.25 | 14.05 | 3.70 | 20.70 | 0.75 | 4.73 | 0.90 | 4.75 | | | | | | | | |
| **Benson et al., 1985** | 9 | 7 | | | | | | | | | | | | | | | | | | | | | | | | | 6.60 | 18.05 | 26.20 | 31.70 | 1.60 | 4.55 | 6.50 | 6.80 | | | | | | | | |
| **Zarcone et al., 1987** | 8 | 12 | | | | | | | | | | | | | | | | | | | | | | | | | | | | | | | | | 42.94 | 32.25 | 54.18 | 35.92 | | | | |
| **Zarcone et al., 1995** | 8 | 11 | 50.26 | | 61.49 | | 15.6 | 5.6 | 16.1 | 4.7 | 163.68 | | 207.37 | | 50.80 | 6.20 | 54.3 | 4.5 | | | | | | | | | | | | | | | | | 43.17 | | 29.79 | | 13.40 | 8.10 | 7.80 | 6.70 |

| Study | N. (SZA)/N. (SCZ) | Age (SZA/SCZ) | Males % (SZA/SCZ) | Diagnosis (SZA/SCZ) | Treatment |
|---|---|---|---|---|---|
| **Reich et al., 1975** | 3/14 | 38.8/20.8 | 50/57 | Schizoaffective disorder/Schizophrenia (acute) | Drug-free (≥2 weeks) |
| **Reich et al., 1975** | 3/9 | 38.8/19 | 50/11 | Schizoaffective disorder/Schizophrenia (latent) | Drug-free (≥2 weeks) |
| **Benson et al., 1980** | 9/9 | ?/? | ?/? | Schizoaffective disorder/ | Drug-free (≥2 weeks) |
| **Benson et al., 1985** | 8/11 | 33/27 | 100/100 | Schizoaffective disorder/Schizophrenia | Drug-free (≥2 weeks) |
| **Benson et al., 1985** | 9/8 | 33.4/26.6 | 100/100 | Schizoaffective disorder/Schizophrenia | Drug-free (≥2 weeks) |
| **Zarcone et al., 1987** | 8/12 | 33.8/26.2 | 100/100 | Schizoaffective disorder/Schizophrenia | Drug-free (≥2 weeks) |
| **Zarcone et al., 1995** | 8/18 | 34.5/33.1 | 100/100 | Schizoaffective disorder/Schizophrenia | Drug-free (2 days) |

| Study | N. SZA | N. SCZ | ST1 (SZA) | SD | ST1 (SCZ) | SD | %ST1 (SZA) | SD | %ST1 (SCZ) | SD | ST2 (SZA) | SD | ST2 (SCZ) | SD | %ST2 (SZA) | SD | %ST2 (SCZ) | SD | ST3 (SZA) | SD | ST3 (SCZ) | SD | %ST3 (SZA) | SD | %ST3 (SCZ) | SD | ST4 (SZA) | SD | ST4 (SCZ) | SD | %ST4 (SZA) | SD | %ST4 (SCZ) | SD | DELTAT (SZA) | SD | DELTAT (SCZ) | SD | %DELTA (SZA) | SD | %DELTA (SCZ) | SD |
|---|---|---|---|---|---|---|---|---|---|---|---|---|---|---|---|---|---|---|---|---|---|---|---|---|---|---|---|---|---|---|---|---|---|---|---|---|---|---|---|---|---|---|
| **Reich et al., 1975** | 3 | 14 | 34.20 | 8.90 | 26.40 | 4.30 | 8.68 | | 7.95 | | 217.80 | 18.30 | 194.70 | 11.80 | 62.98 | | 63.81 | | | | | | | | | | | | | | | | | | 18.20 | 4.50 | 28.90 | 5.70 | 5.26 | | 9.47 | |
| **Reich et al., 1975** | 3 | 9 | 34.20 | 8.90 | 29.80 | 5.30 | 8.68 | | 8.65 | | 217.80 | 18.30 | 220.80 | 10.80 | 62.98 | | 58.93 | | | | | | | | | | | | | | | | | | 18.20 | 4.50 | 43.30 | 7.70 | 5.26 | | 11.56 | |
| **Benson et al., 1980** | 9 | 9 | | | | | | | | | 206.98 | | 199.24 | | | | | | 30.74 | | 36.71 | | | | | | 12.62 | | 21.17 | | 3.17 | | 5.54 | | 43.36 | | 57.88 | | | | | |
| **Benson et al., 1985** | 8 | 11 | | | | | | | | | | | | | | | | | | | | | | | | | 3.25 | 14.05 | 18.00 | 19.30 | 0.75 | 4.73 | 4.98 | 4.20 | | | | | | | | |
| **Benson et al., 1985** | 9 | 8 | | | | | | | | | | | | | | | | | | | | | | | | | 6.60 | 18.05 | 12.25 | 35.40 | 1.60 | 4.55 | 3.30 | 9.78 | | | | | | | | |
| **Zarcone et al., 1987** | 8 | 12 | | | | | | | | | | | | | | | | | | | | | | | | | | | | | | | | | 42.94 | 32.25 | 54.22 | 32.79 | | | | |
| **Zarcone et al., 1995** | 8 | 18 | 50.26 | | 50.53 | | 15.6 | 5.6 | 14.00 | 4.70 | 163.68 | | 198.13 | | 50.80 | 6.20 | 54.90 | 6.30 | | | | | | | | | | | | | | | | | 43.17 | | 36.81 | | 13.40 | 8.10 | 10.20 | 7.20 |

| |
|---|
| Drug-free SZA vs HC |
| Drug-free SZA vs MDD |
| Drug-free SZA vs SCZ |
| Calculated from original data |
| SD not available in original data (averaged from similar available data) |

| Study | N. SZA | ST4 (SZA) | SD | N. HC | ST4 (HC) | SD |
|---|---|---|---|---|---|---|
| Benson et al., 1980 | 9 | 12.62 | 16.17 | 19 | 49.34 | 37.00 |
| Benson et al., 1985 | 8 | 3.25 | 14.05 | 13 | 46.50 | 44.30 |
| Benson et al., 1985 | 9 | 6.60 | 18.05 | 19 | 46.10 | 32.00 |
| | 26 | 7.65 | 16.17 | 51 | 47.41 | 37.00 |

| Study | N. SZA | %ST4 (SZA) | SD | N. HC | %ST4 (HC) | SD |
|---|---|---|---|---|---|---|
| Benson et al., 1980 | 9 | 3.17 | 4.63 | 19 | 11.02 | 8.30 |
| Benson et al., 1985 | 8 | 0.75 | 4.73 | 13 | 10.00 | 10.50 |
| Benson et al., 1985 | 9 | 1.60 | 4.55 | 19 | 10.40 | 6.80 |
| | 26 | 1.88 | 4.63 | 51 | 10.53 | 8.30 |

| Study | N. SZA | DELTAT (SZA) | SD | N. HC | DELTAT (HC) | SD |
|---|---|---|---|---|---|---|
| Benson et al., 1980 | 9 | 43.36 | 32.25 | 19 | 85.31 | 30.90 |
| Zarcone et al., 1987 | 8 | 42.94 | 32.25 | 18 | 83.18 | 30.90 |
| Zarcone et al., 1995 | 8 | 43.17 | 32.25 | 10 | 38.63 | 30.90 |
| | 25 | 43.16 | 32.25 | 47 | 74.56 | 30.90 |

| Study | N. SZA | ST4 (SZA) | SD | N. MDD | ST4 (MDD) | SD |
|---|---|---|---|---|---|---|
| Benson et al., 1980 | 9 | 12.62 | 16.17 | 6 | 19.92 | 25.23 |
| Benson et al., 1985 | 8 | 3.25 | 14.05 | 10 | 3.70 | 20.70 |
| Benson et al., 1985 | 9 | 6.60 | 18.05 | 7 | 26.20 | 31.70 |
| | 26 | 7.65 | 16.17 | 23 | 14.78 | 25.23 |

| Study | N. SZA | %ST4 (SZA) | SD | N. MDD | %ST4 (MDD) | SD |
|---|---|---|---|---|---|---|
| Benson et al., 1980 | 9 | 3.17 | 4.63 | 6 | 4.72 | 5.59 |
| Benson et al., 1985 | 8 | 0.75 | 4.73 | 10 | 0.90 | 4.75 |
| Benson et al., 1985 | 9 | 1.60 | 4.55 | 7 | 6.50 | 6.80 |
| | 26 | 1.88 | 4.63 | 23 | 3.60 | 5.59 |

| Study | N. SZA | DELTAT (SZA) | SD | N. MDD | DELTAT (MDD) | SD |
|---|---|---|---|---|---|---|
| Kupfer & Foster, 1975 | 6 | 18.20 | 4.50 | 9 | 4.40 | 2.20 |
| Kupfer & Foster, 1975 | 6 | 18.20 | 4.50 | 25 | 21.00 | 5.70 |
| Kupfer et al., 1979 | 12 | 1.07 | 15.60 | 29 | 0.81 | 12.90 |
| Benson et al., 1980 | 9 | 43.36 | 15.60 | 6 | 65.03 | 12.90 |
| Zarcone et al., 1987 | 8 | 42.94 | 32.25 | 12 | 54.18 | 35.92 |
| Zarcone et al., 1995 | 8 | 43.17 | 15.60 | 11 | 29.79 | 12.90 |
| | 49 | 26.74 | 15.60 | 92 | 21.26 | 12.90 |

| Study | N. SZA | %DELTA (SZA) | SD | N. MDD | %DELTA (MDD) | SD |
|---|---|---|---|---|---|---|
| Kupfer & Foster, 1975 | 6 | 4.80 | 1.20 | 9 | 1.60 | 0.85 |
| Kupfer et al., 1979 | 12 | 0.40 | 1.10 | 29 | 0.30 | 1.70 |
| Zarcone et al., 1995 | 8 | 13.40 | 8.10 | 11 | 7.80 | 6.70 |
| | 26 | 5.42 | 3.28 | 49 | 2.22 | 2.67 |

| Study | N. SZA | ST1 (SZA) | SD | N. SCZ | ST1 (SCZ) | SD |
|---|---|---|---|---|---|---|
| Reich et al., 1975 | 3 | 34.20 | 8.90 | 14 | 26.40 | 4.30 |
| Reich et al., 1975 | 3 | 34.20 | 8.90 | 9 | 29.80 | 5.30 |
| Zarcone et al., 1995 | 8 | 50.26 | 8.90 | 18 | 50.53 | 4.69 |
| | 14 | 43.38 | 8.90 | 41 | 37.74 | 4.69 |

| Study | N. SZA | %ST1 (SZA) | SD | N. SCZ | %ST1 (SCZ) | SD |
|---|---|---|---|---|---|---|
| Reich et al., 1975 | 3 | 8.68 | 5.6 | 14 | 7.95 | 4.70 |
| Reich et al., 1975 | 3 | 8.68 | 5.6 | 9 | 8.65 | 4.70 |
| Zarcone et al., 1995 | 8 | 15.60 | 5.6 | 18 | 14.00 | 4.70 |
| | 14 | 12.63 | 5.6 | 41 | 10.76 | 4.70 |

| Study | N. SZA | ST2 (SZA) | SD | N. SCZ | ST2 (SCZ) | SD |
|---|---|---|---|---|---|---|
| Reich et al., 1975 | 3 | 217.80 | 18.30 | 14 | 194.70 | 11.80 |
| Reich et al., 1975 | 3 | 217.80 | 18.30 | 9 | 220.80 | 10.80 |
| Benson et al., 1980 | 9 | 206.98 | 18.30 | 9 | 199.24 | 11.41 |
| Zarcone et al., 1995 | 8 | 163.68 | 18.30 | 18 | 198.13 | 11.41 |
| | 23 | 194.74 | 18.30 | 50 | 201.45 | 11.41 |

| Study | N. SZA | %ST2 (SZA) | SD | N. SCZ | %ST2 (SCZ) | SD |
|---|---|---|---|---|---|---|
| Reich et al., 1975 | 3 | 62.98 | 6.20 | 14 | 63.81 | 6.30 |
| Reich et al., 1975 | 3 | 62.98 | 6.20 | 9 | 58.93 | 6.30 |
| Zarcone et al., 1995 | 8 | 50.80 | 6.20 | 18 | 54.90 | 6.30 |
| | 14 | 56.02 | 6.20 | 41 | 58.83 | 6.30 |

| Study | N. SZA | ST4 (SZA) | SD | N. SCZ | ST4 (SCZ) | SD |
|---|---|---|---|---|---|---|
| Benson et al., 1980 | 9 | 12.62 | 16.17 | 9 | 21.17 | 26.08 |
| Benson et al., 1985 | 8 | 3.25 | 14.05 | 11 | 18.00 | 19.30 |
| Benson et al., 1985 | 9 | 6.60 | 18.05 | 8 | 12.25 | 35.40 |
| | 26 | 7.65 | 16.17 | 28 | 17.38 | 26.08 |

| Study | N. SZA | %ST4 (SZA) | SD | N. SCZ | %ST4 (SCZ) | SD |
|---|---|---|---|---|---|---|
| Benson et al., 1980 | 9 | 3.17 | 4.63 | 9 | 5.54 | 6.55 |
| Benson et al., 1985 | 8 | 0.75 | 4.73 | 11 | 4.98 | 4.20 |
| Benson et al., 1985 | 9 | 1.60 | 4.55 | 8 | 3.30 | 9.78 |
| | 26 | 1.88 | 4.63 | 28 | 4.68 | 6.55 |

| Study | N. SZA | DELTAT (SZA) | SD | N. SCZ | DELTAT (SCZ) | SD |
|---|---|---|---|---|---|---|
| Reich et al., 1975 | 3 | 18.20 | 4.50 | 14 | 28.90 | 5.70 |
| Reich et al., 1975 | 3 | 18.20 | 4.50 | 9 | 43.30 | 7.70 |
| Benson et al., 1980 | 9 | 43.36 | 20.36 | 9 | 57.88 | 15.50 |
| Zarcone et al., 1987 | 8 | 42.94 | 32.25 | 12 | 54.22 | 32.79 |
| Zarcone et al., 1995 | 8 | 43.17 | 20.36 | 18 | 36.81 | 15.50 |
| | 31 | 38.33 | 20.36 | 62 | 42.39 | 15.50 |

| Study | N. SZA | %DELTA (SZA) | SD | N. SCZ | %DELTA (SCZ) | SD |
|---|---|---|---|---|---|---|
| Reich et al., 1975 | 3 | 5.26 | 8.10 | 14 | 9.47 | 7.20 |
| Reich et al., 1975 | 3 | 5.26 | 8.10 | 9 | 11.56 | 7.20 |
| Zarcone et al., 1995 | 8 | 13.40 | 8.10 | 18 | 10.20 | 7.20 |
| | 14 | 9.91 | 8.10 | 41 | 10.25 | 7.20 |

| Study | N. (SZA)/N. (HC) | Age (SZA/HC) | Males % (SZA/HC) | Diagnosis (SZA/HC) | Treatment |
|---|---|---|---|---|---|
| **Benson et al., 1980** | 9/19 | ?/? | ?/? | Schizoaffective disorder/Healthy | Drug-free (≥2 weeks) |
| **Benson et al., 1985** | 8/13 | 33/29.7 | 100/100 | Schizoaffective disorder/Healthy | Drug-free (≥2 weeks) |
| **Benson et al., 1985** | 9/19 | 33.4/30.2 | 100/100 | Schizoaffective disorder/Healthy | Drug-free (≥2 weeks) |
| **Zarcone et al., 1987** | 8/18 | 33.8/30.3 | 100/100 | Schizoaffective disorder/Healthy | Drug-free (≥2 weeks) |
| **Zarcone et al., 1995** | 8/10 | 34.5/32.3 | 100/100 | Schizoaffective disorder/Healthy | Drug-free (2 days) |

| Study | N. SZA | N. HC | TST (SZA) | SD | TST (HC) | SD | SLAT (SZA) | SD | SLAT (HC) | SD | SLEFF (SZA) | SD | SLEFF (HC) | SD | WAKE (SZA) | SD | WAKE (HC) | SD |
|---|---|---|---|---|---|---|---|---|---|---|---|---|---|---|---|---|---|---|
| **Benson et al., 1980** | 9 | 19 | | | | | 36.77 | | 9.88 | | | | | | 41.62 | | 8.43 | 10.94 |
| **Benson et al., 1985** | 8 | 13 | 354.25 | 111.20 | 441.40 | 44.05 | 47.5 | 27.88 | 6.2 | 5.40 | | | | | | | | |
| **Benson et al., 1985** | 9 | 19 | 409.10 | 44.70 | 451.80 | 23.50 | 42.70 | 30.00 | 8.50 | 6.60 | | | | | | | | |
| **Zarcone et al., 1987** | 8 | 18 | 394.89 | 36.98 | 451.15 | 17.78 | 60.12 | 32.07 | 16.08 | 10.59 | | | | | 44.45 | 38.67 | 7.72 | 6.08 |
| **Zarcone et al., 1995** | 8 | 10 | 322.20 | 101.40 | 415.40 | 38.40 | 74.80 | 69.10 | 15.00 | 9.90 | 70.90 | 22.20 | 90.70 | 5.40 | 53.00 | 46.00 | 30.40 | 19.70 |

| Study | N. (SZA)/N. (MDD) | Age (SZA/MDD) | Males % (SZA/MDD) | Diagnosis (SZA/MDD) | Treatment |
|---|---|---|---|---|---|
| **Kupfer & Foster, 1975** | 6/9 | 38.8/58.2 | 50/? | Schizoaffective disorder/Psychotic depression | Drug-free (≥2 weeks) |
| **Kupfer et al., 1979** | 12/29 | 44.4/48.1 | 42/31 | Schizoaffective disorder/Psychotic depression | Drug-free (≥2 weeks) |
| **Benson et al., 1980** | 9/6 | ?/? | ?/? | Schizoaffective disorder/ | Drug-free (≥2 weeks) |
| **Benson et al., 1983** | 2/5 | ?/? | 100/100 | Schizoaffective disorder/MDD | Drug-free (≥2 weeks) |
| **Benson et al., 1985** | 8/10 | 33/40.3 | 100/100 | Schizoaffective disorder/MDD | Drug-free (≥2 weeks) |
| **Benson et al., 1985** | 9/7 | 33.4/37.7 | 100/100 | Schizoaffective disorder/MDD | Drug-free (≥2 weeks) |
| **Zarcone et al., 1987** | 8/12 | 33.8/41.33 | 100/100 | Schizoaffective disorder/MDD | Drug-free (≥2 weeks) |
| **Zarcone et al., 1995** | 8/11 | 34.5/37.2 | 100/100 | Schizoaffective disorder/MDD | Drug-free (2 days) |

| Study | N. SZA | N. MDD | TST (SZA) | SD | TST (MDD) | SD | SLAT (SZA) | SD | SLAT (MDD) | SD | SLEFF (SZA) | SD | SLEFF (MDD) | SD | WAKE (SZA) | SD | WAKE (MDD) | SD |
|---|---|---|---|---|---|---|---|---|---|---|---|---|---|---|---|---|---|---|
| **Kupfer & Foster, 1975** | 6 | 9 | 345.80 | 28.40 | 262.90 | 23.70 | | | | | 73.00 | 4.00 | 61.00 | 5.00 | 14.70 | 5.70 | 54.50 | 13.40 |
| **Kupfer et al., 1979** | 12 | 29 | 266.80 | 73.40 | 271.30 | 79.00 | 79.00 | 54.50 | 63.30 | 41.90 | 64.70 | 16.60 | 62.40 | 22.80 | 40.10 | 47.50 | 47.80 | 41.90 |
| **Benson et al., 1980** | 9 | 6 | | | | | 36.77 | | 32.78 | | | | | | 41.62 | | 19.33 | |
| **Benson et al., 1983** | 2 | 5 | 392.90 | | 427.22 | | 57.70 | | 24.36 | | | | | | | | | |
| **Benson et al., 1985** | 8 | 10 | 354.25 | 111.20 | 426.10 | 43.40 | 47.5 | 27.88 | 12.00 | 19.40 | | | | | | | | |
| **Benson et al., 1985** | 9 | 7 | 409.10 | 44.70 | 403.80 | 40.40 | 42.70 | 30.00 | 8.90 | 52.60 | | | | | | | | |
| **Zarcone et al., 1987** | 8 | 12 | 394.89 | 36.98 | 410.74 | 36.99 | 60.12 | 32.07 | 34.69 | 31.41 | | | | | 44.45 | 38.67 | 32.26 | 36.98 |
| **Zarcone et al., 1995** | 8 | 11 | 322.20 | 101.40 | 381.90 | 81.50 | 74.80 | 69.10 | 23.80 | 16.20 | 70.90 | 22.20 | 84.90 | 14.40 | 53.00 | 46.00 | 42.00 | 43.30 |

| Study | N. (SZA)/N. (SCZ) | Age (SZA/SCZ) | Males % (SZA/SCZ) | Diagnosis (SZA/SCZ) | Treatment |
|---|---|---|---|---|---|
| **Reich et al., 1975** | 3/14 | 38.8/20.8 | 50/57 | Schizoaffective disorder/Schizophrenia (acute) | Drug-free (≥2 weeks) |
| **Reich et al., 1975** | 3/9 | 38.8/19 | 50/11 | Schizoaffective disorder/Schizophrenia (latent) | Drug-free (≥2 weeks) |
| **Benson et al., 1980** | 9/9 | ?/? | ?/? | Schizoaffective disorder/Schizophrenia | Drug-free (≥2 weeks) |
| **Benson et al., 1983** | 2/7 | ?/? | 100/100 | Schizoaffective disorder/Schizophrenia | Drug-free (≥2 weeks) |
| **Benson et al., 1985** | 8/11 | 33/27 | 100/100 | Schizoaffective disorder/Schizophrenia | Drug-free (≥2 weeks) |
| **Benson et al., 1985** | 9/8 | 33.4/26.6 | 100/100 | Schizoaffective disorder/Schizophrenia | Drug-free (≥2 weeks) |
| **Zarcone et al., 1987** | 8/12 | 33.8/26.2 | 100/100 | Schizoaffective disorder/Schizophrenia | Drug-free (≥2 weeks) |
| **Zarcone et al., 1995** | 8/18 | 34.5/33.1 | 100/100 | Schizoaffective disorder/Schizophrenia | Drug-free (2 days) |

| Study | N. SZA | N. SCZ | TST (SZA) | SD | TST (SCZ) | SD | SLAT (SZA) | SD | SLAT (SCZ) | SD | SLEFF (SZA) | SD | SLEFF (SCZ) | SD | WAKE (SZA) | SD | WAKE (SCZ) | SD |
|---|---|---|---|---|---|---|---|---|---|---|---|---|---|---|---|---|---|---|
| **Reich et al., 1975** | 3 | 14 | 345.80 | 28.40 | 305.10 | 16.40 | 102.20 | 17.70 | 79.60 | 18.40 | 73.00 | 4.00 | 72.00 | 4.00 | 14.70 | 5.70 | 30.10 | 10.00 |
| **Reich et al., 1975** | 3 | 9 | 345.80 | 28.40 | 374.70 | 11.40 | 102.20 | 17.70 | 43.40 | 5.70 | 73.00 | 4.00 | 87.00 | 2.00 | 14.70 | 5.70 | 6.20 | 3.40 |
| **Benson et al., 1980** | 9 | 9 | | | | | 36.77 | | 38.22 | | | | | | 41.62 | | 12.64 | 20.77 |
| **Benson et al., 1983** | 2 | 7 | 392.90 | | 385.87 | | 57.70 | | 67.10 | | | | | | | | | |
| **Benson et al., 1985** | 8 | 11 | 354.25 | 111.20 | 413.30 | 87.60 | 47.5 | 27.88 | 33.20 | 63.00 | | | | | | | | |
| **Benson et al., 1985** | 9 | 8 | 409.10 | 44.70 | 399.95 | 61.37 | 42.70 | 30.00 | 47.15 | 23.83 | | | | | | | | |
| **Zarcone et al., 1987** | 8 | 12 | 394.89 | 36.98 | 385.77 | 50.50 | 60.12 | 32.07 | 63.99 | 30.58 | | | | | 44.45 | 38.67 | 16.26 | 9.77 |
| **Zarcone et al., 1995** | 8 | 18 | 322.20 | 101.40 | 360.90 | 58.10 | 74.80 | 69.10 | 67.40 | 60.50 | 70.90 | 22.20 | 79.60 | 12.90 | 53.00 | 46.00 | 29.20 | 28.10 |

| Study | N. SZA | TST (SZA) | SD | N. HC | TST (HC) | SD |
|---|---|---|---|---|---|---|
| Benson et al., 1985 | 8 | 354.25 | 111.20 | 13 | 441.40 | 44.05 |
| Benson et al., 1985 | 9 | 409.10 | 44.70 | 19 | 451.80 | 23.50 |
| Zarcone et al., 1987 | 8 | 394.89 | 36.98 | 18 | 451.15 | 17.78 |
| Zarcone et al., 1995 | 8 | 322.20 | 101.40 | 10 | 415.40 | 38.40 |
| | 33 | 371.29 | 72.70 | 60 | 443.29 | 28.72 |

| Study | N. SZA | SLAT (SZA) | SD | N. HC | SLAT (HC) | SD |
|---|---|---|---|---|---|---|
| Benson et al., 1980 | 9 | 36.77 | 39.47 | 19 | 9.88 | 5.40 |
| Benson et al., 1985 | 8 | 47.50 | 27.88 | 13 | 6.2 | 5.40 |
| Benson et al., 1985 | 9 | 42.70 | 30.00 | 19 | 8.50 | 6.60 |
| Zarcone et al., 1987 | 8 | 60.12 | 32.07 | 18 | 16.08 | 10.59 |
| Zarcone et al., 1995 | 8 | 74.80 | 69.10 | 10 | 15.00 | 9.90 |
| | 42 | 51.78 | 39.47 | 79 | 11.00 | 8.09 |

| Study | N. SZA | WAKE (SZA) | SD | N. HC | WAKE (HC) | SD |
|---|---|---|---|---|---|---|
| Benson et al., 1980 | 9 | 41.62 | 42.34 | 19 | 8.43 | 10.94 |
| Zarcone et al., 1987 | 8 | 44.45 | 38.67 | 18 | 7.72 | 6.08 |
| Zarcone et al., 1995 | 8 | 53.00 | 46.00 | 10 | 30.40 | 19.70 |
| | 25 | 46.17 | 42.34 | 47 | 12.83 | 10.94 |

| Study | N. SZA | TST (SZA) | SD | N. MDD | TST (MDD) | SD |
|---|---|---|---|---|---|---|
| Kupfer & Foster, 1975 | 6 | 345.80 | 28.40 | 9 | 262.90 | 23.70 |
| Kupfer et al., 1979 | 12 | 266.80 | 73.40 | 29 | 271.30 | 79.00 |
| Benson et al., 1983 | 2 | 392.90 | 67.65 | 5 | 427.22 | 58.48 |
| Benson et al., 1985 | 8 | 354.25 | 111.20 | 10 | 426.10 | 43.40 |
| Benson et al., 1985 | 9 | 409.10 | 44.70 | 7 | 403.80 | 40.40 |
| Zarcone et al., 1987 | 8 | 394.89 | 36.98 | 12 | 410.74 | 36.99 |
| Zarcone et al., 1995 | 8 | 322.20 | 101.40 | 11 | 381.90 | 81.50 |
| | 53 | 345.56 | 67.65 | 83 | 344.43 | 58.48 |

| Study | N. SZA | SLAT (SZA) | SD | N. MDD | SLAT (MDD) | SD |
|---|---|---|---|---|---|---|
| Kupfer et al., 1979 | 12 | 79.00 | 54.50 | 29 | 63.30 | 41.90 |
| Benson et al., 1980 | 9 | 36.77 | 43.48 | 6 | 32.78 | 33.80 |
| Benson et al., 1983 | 2 | 57.70 | 43.48 | 5 | 24.36 | 33.80 |
| Benson et al., 1985 | 8 | 47.50 | 27.88 | 10 | 12.00 | 19.40 |
| Benson et al., 1985 | 9 | 42.70 | 30.00 | 7 | 8.90 | 52.60 |
| Zarcone et al., 1987 | 8 | 60.12 | 32.07 | 12 | 34.69 | 31.41 |
| Zarcone et al., 1995 | 8 | 74.80 | 69.10 | 11 | 23.80 | 16.20 |
| | 56 | 57.82 | 43.48 | 80 | 37.68 | 33.80 |

| Study | N. SZA | WAKE (SZA) | SD | N. MDD | WAKE (MDD) | SD |
|---|---|---|---|---|---|---|
| Kupfer & Foster, 1975 | 6 | 14.70 | 5.70 | 9 | 54.50 | 13.40 |
| Kupfer et al., 1979 | 12 | 40.10 | 47.50 | 29 | 47.80 | 41.90 |
| Benson et al., 1980 | 9 | 41.62 | 37.69 | 6 | 19.33 | 36.98 |
| Zarcone et al., 1987 | 8 | 44.45 | 38.67 | 12 | 32.26 | 36.98 |
| Zarcone et al., 1995 | 8 | 53.00 | 46.00 | 11 | 42.00 | 43.30 |
| | 43 | 40.08 | 37.69 | 67 | 42.41 | 36.98 |

| Study | N. SZA | SLEFF (SZA) | SD | N. MDD | SLEFF (MDD) | SD |
|---|---|---|---|---|---|---|
| Kupfer & Foster, 1975 | 6 | 73.00 | 4.00 | 9 | 61.00 | 5.00 |
| Kupfer et al., 1979 | 12 | 64.70 | 16.60 | 29 | 62.40 | 22.80 |
| Zarcone et al., 1995 | 8 | 70.90 | 22.20 | 11 | 84.90 | 14.40 |
| | 26 | 68.52 | 15.42 | 49 | 67.19 | 17.64 |

| Study | N. SZA | TST (SZA) | SD | N. SCZ | TST (SCZ) | SD |
|---|---|---|---|---|---|---|
| Benson et al., 1983 | 2 | 392.90 | 65.88 | 7 | 385.87 | 48.07 |
| Benson et al., 1985 | 8 | 354.25 | 111.20 | 11 | 413.30 | 87.60 |
| Benson et al., 1985 | 9 | 409.10 | 44.70 | 8 | 399.95 | 61.37 |
| Zarcone et al., 1987 | 8 | 394.89 | 36.98 | 12 | 385.77 | 50.50 |
| Zarcone et al., 1995 | 8 | 322.20 | 101.40 | 18 | 360.90 | 58.10 |
| Reich et al., 1975 | 3 | 345.80 | 28.40 | 14 | 305.10 | 16.40 |
| Reich et al., 1975 | 3 | 345.80 | 28.40 | 9 | 374.70 | 11.40 |
| | 41 | 368.62 | 65.88 | 79 | 369.82 | 48.07 |

| Study | N. SZA | SLAT (SZA) | SD | N. SCZ | SLAT (SCZ) | SD |
|---|---|---|---|---|---|---|
| Reich et al., 1975 | 3 | 102.20 | 17.70 | 14 | 79.60 | 18.40 |
| Reich et al., 1975 | 3 | 102.20 | 17.70 | 9 | 43.40 | 5.70 |
| Benson et al., 1980 | 9 | 36.77 | 36.12 | 9 | 38.22 | 36.78 |
| Benson et al., 1983 | 2 | 57.70 | 36.12 | 7 | 67.10 | 36.78 |
| Benson et al., 1985 | 8 | 47.50 | 27.88 | 11 | 33.20 | 63.00 |
| Benson et al., 1985 | 9 | 42.70 | 30.00 | 8 | 47.15 | 23.83 |
| Zarcone et al., 1987 | 8 | 60.12 | 32.07 | 12 | 63.99 | 30.58 |
| Zarcone et al., 1995 | 8 | 74.80 | 69.10 | 18 | 67.40 | 60.50 |
| | 44 | 58.06 | 36.12 | 65 | 57.30 | 36.78 |

| Study | N. SZA | WAKE (SZA) | SD | N. SCZ | WAKE (SCZ) | SD |
|---|---|---|---|---|---|---|
| Benson et al., 1980 | 9 | 41.62 | 32.34 | 9 | 12.64 | 14.97 |
| Zarcone et al., 1987 | 8 | 44.45 | 38.67 | 12 | 16.26 | 9.77 |
| Zarcone et al., 1995 | 8 | 53.00 | 46.00 | 18 | 29.20 | 28.10 |
| Reich et al., 1975 | 3 | 14.70 | 5.70 | 14 | 30.10 | 10.00 |
| Reich et al., 1975 | 3 | 14.70 | 5.70 | 9 | 6.20 | 3.40 |
| | 31 | 40.08 | 32.34 | 62 | 21.16 | 14.97 |

| Study | N. SZA | SLEFF (SZA) | SD | N. SCZ | SLEFF (SCZ) | SD |
|---|---|---|---|---|---|---|
| Reich et al., 1975 | 3 | 73.00 | 4.00 | 14 | 72.00 | 4.00 |
| Reich et al., 1975 | 3 | 73.00 | 4.00 | 9 | 87.00 | 2.00 |
| Zarcone et al., 1995 | 8 | 70.90 | 22.20 | 18 | 79.60 | 12.90 |
| | 14 | 71.80 | 14.40 | 41 | 78.63 | 7.47 |

| |
|---|
| Drug-free SZA vs HC |
| Drug-free SZA vs MDD |
| Drug-free SZA vs SCZ |

| |
|---|
| Calculated from original data |
| SD not available in original data (averaged from similar available data) |

| Study (excluded) | N. (SZA)/N. (CTRL) | Age (SZA/CTRL) | Males % (SZA/CTRL) | Diagnosis (SZA/CTRL) | Treatment | Reason of exclusion |
|---|---|---|---|---|---|---|
| **Kupfer et al., 1971** | 4 | 36.48 | 25 | 1 Schizophrenia<br>**1 Schizoaffective disorder**<br>2 Depressive neurosis | Drug-free (≥2 weeks) →<br>Chlorpromazine | Mixed sample |
| **Gillin et al., 1977** | 10 | 25 | 60 | **4 Schizoaffective disorder**<br>2 Acute schizophrenia<br>1 Chronic schizophrenia<br>1 Schizoid personality disorder<br>2 Bipolar disorder | Drug-free (≥2 weeks) →<br>Pimozide | Mixed sample |
| **Gillin et al., 1978** | 7 | 23 | 43 | 3 Chronic schizophrenia<br>1 Acute schizophrenia<br>**2 Schizoaffective disorder**<br>1 Schizoid personality disorder + OCD | Drug-free (≥1 month) →<br>Pimozide | Mixed sample |
| **Keshavan et al., 1996** | 16 | 29.9 | 50 | 11 Schizophrenia<br>**5 Schizoaffective disorder** | Drug-free (≥2 weeks) →<br>**Drug-free (31%)**<br>Neuroleptics (69%)<br>+ Antidepressants<br>+ Mood stabilizers | Mixed medication status/sample |
| **Keshavan et al., 1996** | 15 | 29.7 | ? | Schizophrenia<br>**5 Schizoaffective disorder** | Drug-free (≥2 weeks) -><br>Haloperidol (67%)<br>Fluphenazine (13%)<br>Trifluoperazine (6.7%)<br>Clozapine (6.7%)<br>Risperidone (6.7%)<br>+ Anticholinergics (47%) | Mixed sample |
| **Benson et al., 1996** | 14/15 | 33.6/30.5 | 100/100 | 11 Schizophrenia (presumably chronic)<br>**3 Schizoaffective disorder**/Healthy | Drug-free (2 days) | Mixed sample |
| **Nishino et al., 1998** | 14/14 | 33.3/34.4 | 100/100 | 12 Schizophrenia<br>**2 Schizoaffective disorder**/Healthy | Drug-free (2 days) | Mixed sample |
| **Keshavan et al., 1998** | 30/30 | 30.9/30.9 | 57/57 | 19 Schizophrenia<br>**8 Schizoaffective disorder**<br>3 Schizophreniform disorder/Healthy | Drug-free (Average 97.5 weeks;<br>Median 40 weeks) | Mixed sample |
| **Yamashita et al., 1999** | 1 | 34 | 0 | **Schizoaffective disorder**<br>(with haloperidol-induced akathisia) | Haloperidol + Chlorpromazine<br>+ Biperiden + BZD<br>+ Mood stabilizer<br>→ BZD + Mood stabilizer | Single-case study |
| **Rotenberg et al., 2000** | 20/10 | 45.8/43.3 | 65/60 | 19 Schizophrenia<br>**1 Schizoaffective disorder**/Healthy | Drug-free (≥2 weeks) | Mixed sample |
| **Wetter et al., 2002** | 1 | 31 | 0 | **Schizoaffective disorder**<br>(with risperidone-induced<br>restless leg syndrome) | Risperidone<br>+ Mood stabilizers<br>→ Quetiapine<br>+ Mood stabilizers | Single-case study |
| **Armitage et al., 2004** | 14 | 40.9 | 29 | **3 Schizoaffective disorder**<br>11 Bipolar I disorder | Carbamazepine, BZD,<br>mood stabilizers<br>→ + Clozapine | Mixed sample |
| **Kluge et al., 2014** | 15 | 32.8 | 47 | Schizophrenia<br>**1 Schizoaffective disorder** | Drug-free (≥2 days) -><br>Olanzapine | Mixed sample |
| **Kluge et al., 2014** | 15 | 36.7 | 33 | Schizophrenia<br>**2 Schizoaffective disorder**<br>1 Schizophreniform disorder | Drug-free (≥2 days) -><br>Clozapine | Mixed sample |
| **Göder et al., 2015** | 16/16 | 29.4/28.3 | 56/44 | 14 Chronic schizophrenia<br>**2 Schizoaffective disorder**/Healthy | Antipsychotics (2° Gen., 94%)<br>Antipsychotics (1° Gen., 6%)<br>+ Antidepressants (13%)<br>+ BZD (6%)<br>+ Anticholinergics (6%)<br>+ Promethazine (6%) | Mixed sample |
| **Kaskie et al., 2019** | 27/23 | 23.2/24.7 | 68/72 | 16 Schizophrenia<br>**2 Schizoaffective disorder**<br>4 Unspecified psychosis<br>3 MDD with psychotic features<br>2 Bipolar disorder/Healthy | Antipsychotics (2° Gen., 41%)<br>Antipsychotics (3° Gen., 3.7%)<br>+ Antidepressants (83%)<br>+ Mood stabilizers (25%)<br>+ BZD (17%)<br><br>**15 drug-naive** | Mixed medication status/sample |
| **Gerstenberg et al., 2020** | 12/24 | 16.6/16.5 | 58/46 | 9 Schizophrenia<br>**2 Schizoaffective Disorder**<br>1 Brief Psychotic Disorder/Healthy | Antipsychotics (3° Gen., 58.3%)<br>Antipsychotics (2°+3° Gen., 25.5%)<br>Antipsychotics (2° Gen., 16.2%)<br>+ BZD (25%)<br>+ Anticholinergics (16.7%) | Mixed sample |
| **Petit et al., 2022** | 12/988 | 58.2/59.9 | 42/58 | **Schizoaffective disorder**/Healthy | Medicated (25%)<br>**Drug-free (75%)** | Mixed medication status |
| **Denis et al., 2024** | 17/27 | 21/24 | 56/53 | 7 Schizophrenia<br>**5 Schizoaffective disorder**<br>4 Bipolar disorder<br>1 Unspecified psychosis/Healthy | Antipsychotics (2° Gen., 35%)<br>Antipsychotics (3° Gen., 29%)<br>Antipsychotics (1° Gen., 12%)<br>+ Mood stabilizers (47%)<br>+ Antidepressants (24%)<br>+ Anticholinergics (18%)<br>+ BZD (6%)<br><br>**4 drug-free** | Mixed medication status/sample |

| Study (excluded) | N. SZA | N. CTRL | TST (SZA) | SD | TST (CTRL) | SD | LAT (SZA) | SD | LAT (CTRL) | SD | SLEFF (SZA) | SD | SLEFF (CTRL) | SD |
|---|---|---|---|---|---|---|---|---|---|---|---|---|---|---|
| **Kupfer et al., 1971** | 4 | 4 | 311.30 | 63.90 | 354.30 | 48.30 | 57.30 | 25.00 | 53.90 | 24.90 | | | | |
| **Gillin et al., 1977** | 10 | 10 | 352.00 | 46.00 | 404.00 | 52.00 | | | | | 80.70 | 7.30 | 87.30 | 7.50 |
| **Gillin et al., 1978** | 7 | 7 | 334.00 | 18.00 | 385.00 | 12.00 | | | | | 82.70 | 5.10 | 84.50 | 1.90 |
| **Keshavan et al., 1996** | 16 | 16 | 325.00 | 92.70 | 436.80 | 77.30 | 63.80 | 62.50 | 53.40 | 36.70 | 72.17 | | 83.61 | |
| **Keshavan et al., 1996** | 15 | 15 | 340.40 | 101.90 | 404.40 | 48.60 | 49.60 | 39.50 | 21.70 | 12.20 | 74.73 | | 90.01 | |
| **Benson et al., 1996** | 14 | 15 | 334.20 | 56.00 | 446.50 | 17.80 | 92.40 | 71.50 | 15.70 | 11.10 | 74.80 | 14.20 | 94.60 | 2.60 |
| **Nishino et al., 1998** | 14 | 14 | 365.90 | 57.80 | 425.80 | 28.30 | 71.60 | 62.80 | 17.30 | 12.40 | 78.90 | 13.40 | 91.40 | 4.10 |
| **Keshavan et al., 1998** | 30 | 30 | 347.60 | 69.80 | 429.40 | 44.00 | 51.80 | 52.10 | 10.60 | 7.10 | 77.90 | 13.90 | 94.40 | 3.30 |
| **Yamashita et al., 1999** | 1 | 1 | | | | | | | | | 86.80 | | 84.40 | |
| **Rotenberg et al., 2000** | 20 | 10 | 413.00 | 74.00 | 412.00 | 43.00 | 32.00 | 69.00 | 3.30 | 2.10 | 92.00 | 11.00 | 98.00 | 3.00 |
| **Wetter et al., 2002** | 1 | 1 | 412.50 | | 470.50 | | 71.50 | | 14.50 | | 85.30 | | 96.60 | |
| **Armitage et al., 2004** | 14 | 14 | 406.30 | 74.50 | 495.80 | 94.70 | 20.90 | 23.30 | 44.20 | 30.90 | 92.50 | 6.40 | 91.90 | 4.50 |
| **Kluge et al., 2014** | 15 | 15 | 350.90 | 75.40 | 459.60 | 24.80 | 42.20 | 30.20 | 19.40 | 20.40 | 83.00 | 14.00 | 97.00 | 1.00 |
| **Kluge et al., 2014** | 15 | 15 | 354.40 | 90.70 | 457.20 | 24.00 | 29.90 | 26.60 | 14.00 | 18.00 | 82.00 | 13.00 | 97.00 | 3.00 |
| **Göder et al., 2015** | 16 | 16 | 428.70 | 72.90 | 369.70 | 42.20 | 34.60 | 35.30 | 21.00 | 17.60 | 85.80 | 8.90 | 86.60 | 7.80 |
| **Kaskie et al., 2019** | 27 | 23 | 389.00 | 88.00 | 428.00 | 50.00 | 43.00 | 53.00 | 15.00 | 12.00 | | | | |
| **Gerstenberg et al., 2020** | 12 | 24 | | | | | 24.10 | 16.60 | 16.10 | 9.00 | 91.50 | 5.50 | 91.70 | 5.00 |
| **Petit et al., 2022** | 12 | 988 | 386.40 | 78.60 | 394.80 | 69.00 | 31.00 | | 16.00 | | 78.80 | | 83.90 | |
| **Dan et al., 2024** | 17 | 27 | 507.00 | 54.00 | 508.00 | 74.00 | 20.00 | 20.00 | 25.00 | 29.00 | 85.00 | 9.00 | 86.00 | 8.00 |

| Study (excluded) | WAKE (SZA) | SD | WAKE (CTRL) | SD | REMT (SZA) | SD | REMT (CTRL) | SD | %REM (SZA) | SD | %REM (CTRL) | SD | REML (SZA) | SD | REML (CTRL) | SD | REMD (SZA) | SD | REMD (CTRL) | SD |
|---|---|---|---|---|---|---|---|---|---|---|---|---|---|---|---|---|---|---|---|---|
| **Kupfer et al., 1971** | 54.80 | 37.90 | 21.10 | 17.40 | 59.80 | 23.90 | 65.30 | 17.00 | 19.10 | 3.80 | 18.50 | 4.10 | 57.90 | 22.60 | 60.30 | 41.10 | 1.61 | | 1.66 | |
| **Gillin et al., 1977** | 14.00 | 21.00 | 11.00 | 8.00 | 82.00 | 15.00 | 95.00 | 16.00 | 22.80 | 3.50 | 24.30 | 3.20 | 64.00 | 18.00 | 65.00 | 35.00 | 1.70 | 0.40 | 1.50 | 0.50 |
| **Gillin et al., 1978** | | | | | 80.00 | 6.00 | 92.00 | 7.00 | 23.00 | 1.60 | 21.10 | 1.80 | 46.00 | 13.00 | 64.00 | 13.00 | 1.80 | 0.20 | 1.80 | 0.10 |
| **Keshavan et al., 1996** | 61.50 | 84.00 | 32.20 | 19.30 | 76.20 | 35.30 | 98.90 | 35.30 | 22.30 | 8.30 | 22.20 | 5.60 | 61.20 | 24.40 | 84.60 | 44.20 | 1.29 | 0.45 | 1.57 | 0.44 |
| **Keshavan et al., 1996** | 65.60 | 85.50 | 17.80 | 15.50 | 77.60 | 39.00 | 91.90 | 23.70 | 21.60 | 8.00 | 22.70 | 4.90 | 56.00 | 26.50 | 76.20 | 36.80 | 1.40 | 0.70 | 1.50 | 0.80 |
| **Benson et al., 1996** | 28.00 | 26.30 | 7.20 | 6.70 | 63.83 | | 103.59 | | 19.10 | 3.20 | 23.20 | 3.20 | 62.60 | 21.10 | 94.40 | 44.70 | | | | |
| **Nishino et al., 1998** | 30.00 | 25.80 | 25.90 | 17.60 | 75.70 | 22.90 | 93.20 | 27.40 | 20.70 | 4.40 | 22.00 | 5.80 | 65.30 | 21.90 | 73.20 | 13.30 | | | | |
| **Keshavan et al., 1998** | 49.30 | 43.20 | 15.00 | 13.60 | 78.60 | 27.40 | 107.60 | 25.70 | 22.60 | 6.30 | 24.90 | 5.10 | 60.90 | 27.00 | 69.10 | 30.30 | 1.50 | 0.60 | 1.40 | 0.50 |
| **Yamashita et al., 1999** | | | | | | | | | 17.80 | | 20.60 | | | | | | | | | |
| **Rotenberg et al., 2000** | | | | | 93.75 | | 79.52 | | 22.70 | 8.00 | 19.30 | 3.70 | 93.40 | 6.10 | 89.40 | 33.70 | 5.10 | 3.55 | 4.07 | 2.54 |
| **Wetter et al., 2002** | 71.00 | | 13.00 | | 70.00 | | 66.50 | | 16.97 | | 14.13 | | | | | | | | | |
| **Armitage et al., 2004** | 24.78 | | 32.72 | | 51.60 | | 62.97 | | 12.70 | 5.70 | 12.70 | 5.20 | 115.90 | 41.90 | 118.70 | 49.20 | 2.50 | 1.10 | 2.90 | 0.80 |
| **Kluge et al., 2014** | | | | | 77.70 | 24.20 | 115.20 | 25.50 | 22.14 | | 25.06 | | 116.40 | 57.60 | 88.00 | 55.90 | 2.62 | 1.53 | 3.25 | 1.05 |
| **Kluge et al., 2014** | | | | | 77.90 | 34.70 | 99.90 | 44.50 | 21.98 | | 21.85 | | 94.00 | 40.50 | 105.00 | 48.00 | 3.11 | 0.84 | 3.07 | 1.45 |
| **Göder et al., 2015** | | | | | 87.88 | | 68.76 | | 20.50 | 5.10 | 18.60 | 4.60 | | | | | 14.10 | 6.70 | 11.00 | 4.00 |
| **Kaskie et al., 2019** | 77.00 | 59.00 | 59.00 | 34.00 | 81.69 | | 81.32 | | 21.00 | 7.00 | 19.00 | 6.00 | | | | | | | | |
| **Gerstenberg et al., 2020** | 22.60 | 20.40 | 25.00 | 17.80 | | | | | | | | | 152.20 | 67.40 | 122.50 | 45.20 | | | | |
| **Petit et al., 2022** | 107.00 | | 78.00 | | 85.00 | | 87.00 | | 21.70 | | 21.80 | | 114.00 | | 87.00 | | | | | |
| **Dan et al., 2024** | 65.00 | 49.00 | 52.00 | 41.00 | 93.00 | 42.00 | 113.00 | 33.00 | 18.10 | 7.60 | 21.90 | 5.10 | | | | | | | | |

| Study (excluded) | ST1 (SZA) | SD | ST1 (CTRL) | SD | %ST1 (SZA) | SD | %ST1 (CTRL) | SD | ST2 (SZA) | SD | ST2 (CTRL) | SD | %ST2 (SZA) | SD | %ST2 (CTRL) | SD |
|---|---|---|---|---|---|---|---|---|---|---|---|---|---|---|---|---|
| **Kupfer et al., 1971** | | | | | | | | | | | | | | | | |
| **Gillin et al., 1977** | 33.00 | 51.00 | 29.00 | 18.00 | 5.30 | 3.30 | 7.20 | 4.40 | 216.00 | 25.00 | 255.00 | 32.00 | 59.90 | 5.00 | 63.80 | 6.00 |
| **Gillin et al., 1978** | 15.00 | 3.00 | 30.00 | 7.00 | 4.30 | 1.10 | 7.60 | 5.20 | 228.00 | 14.00 | 265.00 | 23.00 | 63.30 | 2.80 | 65.80 | 3.70 |
| **Keshavan et al., 1996** | | | | | | | | | | | | | | | | |
| **Keshavan et al., 1996** | | | | | | | | | | | | | | | | |
| **Benson et al., 1996** | 48.79 | | 27.24 | | 14.60 | 7.00 | 6.10 | 2.40 | 179.13 | | 229.05 | | 53.60 | 7.80 | 51.30 | 5.40 |
| **Nishino et al., 1998** | 50.90 | 23.40 | 60.10 | 20.60 | 13.90 | 5.40 | 14.60 | 5.30 | 183.50 | 37.40 | 231.10 | 37.40 | 51.30 | 7.00 | 55.30 | 8.60 |
| **Keshavan et al., 1998** | | | | | | | | | | | | | | | | |
| **Yamashita et al., 1999** | | | | | 13.20 | | 9.90 | | | | | | 59.40 | | 50.40 | |
| **Rotenberg et al., 2000** | | | | | | | | | | | | | | | | |
| **Wetter et al., 2002** | 39.50 | | 56.00 | | | | | | 231.50 | | 202.00 | | | | | |
| **Armitage et al., 2004** | 78.01 | | 61.98 | | 19.20 | 10.20 | 12.50 | 6.70 | 246.62 | | 322.27 | | 60.70 | 12.90 | 65.00 | 9.90 |
| **Kluge et al., 2014** | 35.20 | 20.90 | 31.50 | 19.10 | 10.03 | | 6.85 | | 183.40 | 60.70 | 206.70 | 40.70 | 52.26 | | 44.97 | |
| **Kluge et al., 2014** | 29.60 | 15.10 | 20.10 | 12.40 | 8.35 | | 4.40 | | 205.10 | 45.20 | 296.60 | 49.30 | 57.87 | | 64.87 | |
| **Göder et al., 2015** | 51.44 | | 42.52 | | 12.00 | 8.30 | 11.50 | 4.20 | 222.07 | | 184.48 | | 51.80 | 8.40 | 49.90 | 7.60 |
| **Kaskie et al., 2019** | 31.12 | | 34.24 | | 8.00 | 6.00 | 8.00 | 4.00 | 206.17 | | 231.12 | | 53.00 | 8.00 | 54.00 | 6.00 |
| **Gerstenberg et al., 2020** | | | | | | | | | | | | | | | | |
| **Petit et al., 2022** | 47.00 | | 47.00 | | 12.40 | | 12.20 | | 160.00 | | 185.00 | | 41.60 | | 46.90 | |
| **Dan et al., 2024** | 34.00 | 40.00 | 31.00 | 23.00 | 6.90 | 8.20 | 6.10 | 4.70 | 259.00 | 57.00 | 262.00 | 54.00 | 23.80 | 8.60 | 20.50 | 6.50 |

| Study (excluded) | ST3 (SZA) | SD | ST3 (CTRL) | SD | %ST3 (SZA) | SD | %ST3 (CTRL) | SD | ST4 (SZA) | SD | ST4 (CTRL) | SD | %ST4 (SZA) | SD | %ST4 (CTRL) | SD | DELTAT (SZA) | SD | DELTAT (CTRL) | SD | %DELTA (SZA) | SD | %DELTA (CTRL) | SD |
|---|---|---|---|---|---|---|---|---|---|---|---|---|---|---|---|---|---|---|---|---|---|---|---|---|
| **Kupfer et al., 1971** | | | | | | | | | | | | | | | | | | | | | 9.20 | 9.70 | 10.10 | 12.00 |
| **Gillin et al., 1977** | | | | | | | | | | | | | | | | | 38.00 | 21.00 | 21.00 | 16.00 | 10.90 | 5.50 | 6.70 | 5.50 |
| **Gillin et al., 1978** | | | | | | | | | | | | | | | | | 32.00 | 9.00 | 13.00 | 4.00 | 9.70 | 2.30 | 3.50 | 1.20 |
| **Keshavan et al., 1996** | | | | | | | | | | | | | | | | | 48.40 | 26.60 | 52.20 | 35.40 | 16.90 | 13.00 | 12.80 | 8.70 |
| **Keshavan et al., 1996** | | | | | | | | | | | | | | | | | 44.00 | 29.90 | 56.20 | 37.10 | 14.20 | 11.50 | 13.90 | 8.90 |
| **Benson et al., 1996** | 39.44 | | 37.51 | | 11.80 | 6.70 | 8.40 | 4.20 | 3.01 | | 49.56 | | 0.90 | 1.70 | 11.10 | 4.70 | | | | | | | | |
| **Nishino et al., 1998** | | | | | | | | | | | | | | | | | 49.00 | 23.90 | 34.20 | 30.00 | 14.10 | 8.20 | 8.10 | 6.90 |
| **Keshavan et al., 1998** | | | | | | | | | | | | | | | | | 35.00 | 30.10 | 53.10 | 27.40 | 9.90 | 8.30 | 12.20 | 5.90 |
| **Yamashita et al., 1999** | | | | | 0.70 | | 5.10 | | | | | | 0.00 | | 0.00 | | | | | | 0.70 | | 5.10 | |
| **Rotenberg et al., 2000** | | | | | | | | | | | | | | | | | 67.32 | | 59.74 | | 16.30 | 10.10 | 14.50 | 6.80 |
| **Wetter et al., 2002** | | | | | | | | | | | | | | | | | 67.50 | | 143.00 | | 16.37 | | 30.39 | |
| **Armitage et al., 2004** | | | | | | | | | | | | | | | | | 5.28 | | 15.37 | | 1.30 | 2.30 | 3.10 | 5.30 |
| **Kluge et al., 2014** | 21.90 | 15.50 | 47.00 | 22.70 | 6.24 | | 10.23 | | 32.30 | 30.50 | 50.10 | 34.60 | 9.20 | | 10.90 | | 54.20 | 39.70 | 95.80 | 47.60 | 15.45 | | 20.84 | |
| **Kluge et al., 2014** | 24.30 | 22.60 | 22.60 | 10.40 | 6.86 | | 4.94 | | 16.60 | 23.10 | 15.80 | 18.50 | 4.68 | | 3.46 | | 41.00 | 39.30 | 38.40 | 37.00 | 11.57 | | 8.40 | |
| **Göder et al., 2015** | | | | | | | | | | | | | | | | | 67.73 | | 73.94 | | 15.80 | 9.50 | 20.00 | 7.20 |
| **Kaskie et al., 2019** | | | | | | | | | | | | | | | | | 66.13 | | 77.04 | | 17.00 | 9.00 | 18.00 | 7.00 |
| **Gerstenberg et al., 2020** | | | | | | | | | | | | | | | | | | | | | | | | |
| **Petit et al., 2022** | | | | | | | | | | | | | | | | | 94.00 | | 75.00 | | 24.20 | | 19.10 | |
| **Dan et al., 2024** | | | | | | | | | | | | | | | | | 121.00 | 47.00 | 102.00 | 31.00 | 23.80 | 8.60 | 20.50 | 6.50 |

| Study | N. SZA/Sex/(Age) | Diagnosis | Medication status | TST | SD | LAT | SD | SLEFF | SD | WAKE | SD | REMT | SD | %REM | SD | REML | SD | REMD | SD | ST1 | SD | %ST1 | SD | ST2 | SD | %ST2 | SD | ST3 | SD | %ST3 | SD | ST4 | SD | %ST4 | SD | DELTAT | SD | %DELTA | SD |
|---|---|---|---|---|---|---|---|---|---|---|---|---|---|---|---|---|---|---|---|---|---|---|---|---|---|---|---|---|---|---|---|---|---|---|---|---|---|---|---|
| **Hauri et al., 1973** | 2F (28.5) | Schizoaffective disorder | Drug-free (≥1 week) | 411.50 | | 8.00 | | | | | | 113.57 | | 27.60 | | | | | | 24.69 | | 6.00 | | 231.47 | | 56.25 | | 14.20 | | 3.45 | | 0.00 | | 0.00 | | | | | |
| **Gillin et al., 1981** | 3M, 3F (31) | 4 Chronic schizophrenia<br>**2 Schizoaffective disorder** | Drug-free (≥1 week) | 357.80 | 78.90 | 62.00 | 43.30 | 80.00 | 11.80 | | | 112.30 | 20.60 | 32.30 | 5.10 | 76.90 | 32.30 | 2.50 | 0.50 | 18.00 | 11.30 | 5.30 | 3.20 | 178.80 | 70.60 | 48.00 | 12.50 | | | | | | | | | 48.00 | 25.30 | 14.50 | 8.80 |
| **Glassman et al., 1986** | 1M (39) | Schizoaffective disorder | Lithium carbonate, chlorpromazine, triazolam, benztropine<br>(medication-induced somnambulism with REM suppression) | 508.00 | | | | | | | | 24.00 | | 4.72 | | 480.00 | | | | 4.00 | | 0.79 | | 390.00 | | 76.80 | | 10.00 | | 1.97 | | 80.00 | | 15.75 | | 90.00 | | 17.72 | |
| **Lahmeyer et al., 1988** | 2F (29) | Schizoaffective disorder | Drug-free (2 weeks) | | | | | | | | | | | | | 80.13 | | 1.32 | | | | | | | | | | | | | | | | | | | | | |
| **Keshavan et al., 1991** | 7M, 5F (31.2) | 8 Schizophrenia<br>**3 Schizoaffective disorder**<br>1 Schizotypal disorder | Drug-free (≥2 weeks)<br>2 drug-naive | | | | | | | | | | | | | 65.10 | 46.50 | | | | | | | | | | | | | | | | | | | | | 16.00 | 8.70 |
| **Keshavan et al., 1992** | 14M, 5F (30.5) | 17 Schizophrenia<br>**2 Schizoaffective disorder** | Drug-free (≥2 weeks)<br>6 drug-naive | | | | | | | | | | | | | 65.00 | 32.40 | | | | | | | | | | | | | | | | | | | | | | |
| **Douglass et al., 1993** | 6? (?) | **Schizoaffective disorder**<br>Schizophrenia | Drug-free (≥2 weeks) | 230.00 | 78.00 | 92.70 | 58.00 | 64.40 | 22.00 | 30.50 | 52.00 | 58.60 | 20.00 | 22.60 | 6.50 | 41.60 | 102.00 | 1.40 | 0.75 | 87.63 | | 38.10 | 25.00 | 108.10 | | 47.00 | 12.00 | | | | | | | | | | | | |
| **Douglass et al., 1993** | 3? (?) | Schizophrenia<br>**Schizoaffective disorder**<br>+ **narcolepsy** | Drug-free (≥2 weeks) | 274.00 | | 27.30 | | 71.70 | | 81.30 | | 36.30 | | 13.00 | | 124.70 | | 1.50 | | 140.56 | | 51.30 | | 97.27 | | 35.50 | | | | | | | | | | | | | |
| **Keshavan et al., 1994** | 8M, 11F (30.6) | 13 Schizophrenia<br>**6 Schizoaffective disorder**<br>(suicidal behavior) | Drug-free (≥2 weeks) | | | | | | | | | 96.00 | 24.55 | | | 54.82 | 27.54 | 1.69 | 0.70 | | | | | | | | | | | | | | | | | | | | |
| **Keshavan et al., 1994** | 12M, 8F (32.3) | 16 Schizophrenia<br>**4 Schizoaffective disorder**<br>(no suicidal behavior) | Drug-free (≥2 weeks) | | | | | | | | | 75.05 | 26.70 | | | 58.10 | 28.32 | 1.50 | 0.47 | | | | | | | | | | | | | | | | | | | | |
| **Keshavan et al., 1995** | 10M, 10F (30) | **8 Schizoaffective disorder**<br>12 Schizophrenia | Drug-free (≥4 weeks)<br>12 drug-naive | 315.00 | 97.00 | | | 70.29 | | | | | | | | | | | | | | | | | | | | | | | | | | | | 46.90 | 41.70 | 16.70 | 16.20 |
| **Baandrup et al., 2016** | 5M, 5F (49) | 6 Schizophrenia<br>**2 Schizoaffective disorder**<br>2 Bipolar disorder | Antipsychotics<br>+ BZD<br>+ Antidepressants<br>+ Mood stabilizers<br>+ Anticholinergics | 469.30 | 124.60 | | | 84.90 | | | | 74.15 | | 15.80 | 5.00 | 163.80 | 65.50 | | | 55.38 | | 11.80 | 7.50 | 313.96 | | 66.90 | 8.30 | 25.81 | | 5.50 | 5.90 | | | | | | | | |
| **Weinhold et al., 2022** | 9M, 9F (40.7) | 11 Schizophrenia<br>**7 Schizoaffective disorder** | Atypical antipsychotics<br>+ Antidepressants<br>+ Mood stabilizers | 442.00 | 13.00 | 32.00 | 5.00 | 86.30 | 1.80 | 67.00 | 36.00 | 68.00 | 5.00 | 15.38 | | 130.00 | 15.00 | | | 49.00 | 5.00 | 11.09 | | 176.00 | 11.00 | 39.82 | | 149.00 | 11.00 | 33.71 | | | | | | | | | |
| | | | **Study summary (no clear sleep data)** | | | | | | | | | | | | | | | | | | | | | | | | | | | | | | | | | | | | |
| **Giedke et al., 1992** | 12M, 18F (48) | **4 Schizoaffective disorder**<br>26 MDD | All patients on neuroleptics, antidepressants and BZD<br>Sleep deprivation significantly improved depressed mood | | | | | | | | | | | | | | | | | | | | | | | | | | | | | | | | | | | | |
| **Keshavan et al., 1992** | 11M, 5F (31.65) | 10 Schizophrenia<br>**4 Schizoaffective disorder**<br>1 Atypical psychosis<br>1 Psychotic affective disorder | Reduced NREM sleep<br>REM activity increased, REML reduced (drug-free ≥2 days)<br>Neuroleptic treatment did not improve sleep | | | | | | | | | | | | | | | | | | | | | | | | | | | | | | | | | | | | |

| | Meta-analysis | | | | | Heterogeneity | | | Publication Bias | | | Funnel plot asymmetry | |
|---|---|---|---|---|---|---|---|---|---|---|---|---|---|
| **SZA vs HC** | **N. of studies** | **N. of subjects** | **Mean ± SD** | **RE Model** | **p-value** | **Q** | **I²** | **p-value** | **Fail-Safe N (p-value)** | **Outlier** | **Overly influential** | **Rank correlation (p-value)** | **Regression test (p-value)** |
| **TST** | 4 | 33 vs 60 | 371.29 ± 72.7 vs 443.29 ± 28.72 | -1.43 (-1.903 to -0.949) | **0.001** | 2.797 | 0% | p=0.424 | 48 (p<0.001) | - | - | p=0.7500 | p=0.4260 |
| **LAT** | 5 | 42 vs 79 | 51.78 ± 39.47 vs 11 ± 8.09 | 1.70 (1.236 to 2.159) | **0.001** | 4.379 | 10.43% | p=0.357 | 105 (p<0.001) | - | Benson et al., 1980 | p=0.0833 | p=0.1477 |
| **WAKE** | 3 | 25 vs 47 | 46.17 ± 42.34 vs 12.83 ± 10.94 | 1.20 (0.649 to 1.746) | **0.001** | 2.262 | 6.62% | p=0.323 | 19 (p<0.001) | - | - | p=1.0000 | p=0.8293 |
| **REMT** | 4 | 33 vs 60 | 79.7 ± 29.22 vs 96.85 ± 20.05 | -0.620 (-1.058 to -0.181) | **0.006** | 1.495 | 0% | p=0.683 | 8 (p=0.002) | - | - | p=0.3333 | p=0.2678 |
| **%REM** | 5 | 42 vs 79 | 21.65 ± 5.98 vs 22.3 ± 5.02 | -0.038 (-0416 to 0.339) | 0.843 | 1.722 | 0% | p=0.787 | 0 (p=0.399) | - | - | p=0.4833 | p=0.3592 |
| **REML** | 5 | 42 vs 60 | 66.35 ± 28.33 vs 86.26 ± 34.76 | -0.528 (-0.911 to -0.145) | **0.007** | 1.369 | 0% | p=0.849 | 9 (p=0.003) | - | - | p=0.8167 | p=0.8123 |
| **ST4** | 3 | 26 vs 51 | 7.65 ± 16.17 vs 47.41 ± 37.00 | -1.20 (-1.713 to -0.693) | **0.001** | 0.166 | 0% | p=0.920 | 21 (p<0.001) | - | - | p=1.0000 | p=0.9619 |
| **%ST4** | 3 | 26 vs 51 | 1.88 ± 4.63 vs 10.53 ± 8.30 | -1.14 (-1.649 to -0.636) | **0.001** | 0.429 | 0% | p=0.807 | 19 (p<0.001) | - | - | p=1.0000 | p=0.9100 |
| **DELTAT** | 3 | 25 vs 47 | 43.16 ± 32.25 vs 74.56 ± 30.90 | -0.814 (-1.725 to 0.097) | 0.08 | 6.107 | 67.66% | p=0.047 | 8 (p<0.001) | Zarcone et al., 1995 | - | p=0.3333 | p=0.0874 |

| | Meta-analysis | | | | | Heterogeneity | | | Publication Bias | | | Funnel plot asymmetry | |
|---|---|---|---|---|---|---|---|---|---|---|---|---|---|
| **SZA vs MDD** | **N. of studies** | **N. of subjects** | **Mean ± SD** | **RE Model** | **p-value** | **Q** | **I²** | **p-value** | **Fail-Safe N (p-value)** | **Outlier** | **Overly influential** | **Rank correlation (p-value)** | **Regression test (p-value)** |
| **TST** | 7 | 53 vs 83 | 345.56 ± 67.65 vs 344.43 ± 58.48 | 0.028 (-0.827 to 0.884) | 0.948 | 21.237 | 80.04% | p=0.002 | 0 (p=0.431) | Kupfer & Foster, 1975 | Kupfer & Foster, 1975 | p=0.7726 | p=0.1983 |
| **LAT** | 7 | 56 vs 80 | 57.82 ± 43.48 vs 37.68 ± 33.80 | 0.684 (0.321 to 1.047) | **0.001** | 4.931 | 0% | p=0.553 | 31 (p<0.001) | - | - | p=1.0000 | p=0.4002 |
| **SLEFF** | 3 | 26 vs 49 | 68.52 ± 15.42 vs 67.19 ± 17.64 | 0.535 (-1.247 to 2.316) | 0.556 | 14.418 | 90.44% | p<0.001 | 0 (p=0.093) | Kupfer & Foster, 1975 | - | p=1.0000 | p=0.2274 |
| **WAKE** | 5 | 43 vs 67 | 40.08 ± 37.69 vs 42.41 ± 36.98 | -0.385 (-1.649 to 0.879) | 0.55 | 19.199 | 88.48% | p<0.001 | 0 (p=0.137) | Kupfer & Foster, 1975 | Kupfer & Foster, 1975 | p=1.0000 | **p=0.0239*** |
| **REMT** | 7 | 53 vs 83 | 72.89 ± 25.6 vs 78.23 ± 35.85 | 0.009 (-0.861 to 0.879) | 0.984 | 20.393 | 80.67% | p=0.002 | 0 (p=0.349) | Kupfer & Foster, 1975 | Kupfer & Foster, 1975 | p=0.2389 | p=0.0652 |
| **%REM** | 8 | 62 vs 89 | 21.08 ± 5.65 vs 22.71 ± 6.71 | -0.246 (-0.689 to 0.198) | 0.277 | 11.842 | 37.83% | p=0.106 | 0 (p=0.092) | Kupfer & Foster, 1975 | - | p=0.2751 | p=0.3701 |
| **REML** | 10 | 67 vs 131 | 54.39 ± 25.57 vs 45.52 ± 22.90 | 0.033 (-0.367 to 0.434) | 0.87 | 14.28 | 35.00% | p=0.113 | 0 (p=0.439) | - | - | p=1.000 | p=0.8657 |
| **REMD** | 3 | 20 vs 43 | 1.77 ± 0.71 vs 1.84 ± 0.95 | -0.150 (-0.684 to 0.384) | 0.582 | 0.152 | 0% | p=0.927 | 0 (p=0.342) | - | - | p=1.000 | p=0.7320 |
| **ST4** | 3 | 26 vs 23 | 7.65 ± 16.17 vs 14.78 ± 25.23 | -0.348 (-0.922 to 0.225) | 0.234 | 1.051 | 0% | p=0.591 | 0 (p=0.110) | - | - | p=1.000 | p=0.4381 |
| **%ST4** | 3 | 26 vs 23 | 1.88 ± 4.63 vs 3.60 ± 5.59 | -0.357 (-0.932 to 0.217) | 0.223 | 1.281 | 0% | p=0.527 | 0 (p=0.104) | - | - | p=1.000 | p=0.4131 |
| **DELTAT** | 6 | 49 vs 92 | 26.74 ± 15.60 vs 21.26 ± 12.90 | 0.351 (-0.991 to 1.693) | 0.609 | 30.314 | 91.28% | p<0.001 | 0 (p=0.184) | Kupfer & Foster, 1975 | - | p=0.7194 | **p=0.0379*** |
| **%DELTA** | 3 | 26 vs 49 | 5.42 ± 3.28 vs 2.22 ± 2.67 | 1.16 (-0.501 to 2.821) | 0.171 | 12.554 | 88.30% | p=0.002 | 9 (p<0.001) | Kupfer & Foster, 1975 | - | p=0.3333 | **p=0.0005*** |

| | Meta-analysis | | | | | Heterogeneity | | | Publication Bias | | | Funnel plot asymmetry | |
|---|---|---|---|---|---|---|---|---|---|---|---|---|---|
| **SZA vs SCZ** | **N. of studies** | **N. of subjects** | **Mean ± SD** | **RE Model** | **p-value** | **Q** | **I²** | **p-value** | **Fail-Safe N (p-value)** | **Outlier** | **Overly influential** | **Rank correlation (p-value)** | **Regression test (p-value)** |
| **TST** | 7 | 41 vs 79 | 368.62 ± 65.88 vs 369.82 ± 48.07 | -0.052 (-0.770 to 0.666) | 0.887 | 15.949 | 66.73% | p=0.014 | 0 (p=0.378) | Reich et al., 1975.1 | - | p=1.0000 | p=0.6722 |
| **LAT** | 6 | 44 vs 65 | 58.06 ± 36.12 vs 57.30 ± 36.78 | 0.210 (-0.160 to 0.581) | 0.266 | 20.661 | 0% | p=0.004 | 5 (p=0.018) | Reich et al., 1975.2 | - | p=0.3988 | **p=0.0003*** |
| **SLEFF** | 3 | 14 vs 41 | 71.80 + 14.40 vs 78.63 ± 7.47 | -1.63 (-4.714 to 1.458) | 0.301 | 14.999 | 93.61% | p<0.001 | 7 (p=0.002) | Reich et al., 1975.2 | - | p=1.0000 | **p=0.0343*** |
| **WAKE** | 5 | 31 vs 62 | 40.08 ± 32.34 vs 21.16 ± 14.97 | 0.664 (-0.375 to 1.703) | 0.21 | 14.377 | 77.93% | p=0.006 | 10 (p=0.003) | Reich et al., 1975.1 | - | p=0.4833 | p=0.8658 |
| **REMT** | 7 | 41 vs 79 | 80.4 ± 25.6 vs 83.17 ±18.53 | 0.218 (-0.845 to 1.281) | 0.688 | 24.28 | 84.39% | p<0.001 | 0 (p=0.382) | Reich et al., 1975.1 | Reich et al., 1975.1 | p=0.0690 | **p=0.0005*** |
| **%REM** | 8 | 50 vs 88 | 21.89 ± 5.36 vs 19.74 ± 2.86 | 1.06 (-0.724 vs 2.850) | 0.244 | 61.892 | 94.47% | p<0.001 | 13 (p=0.005) | Zarcone et al., 1987 | Zarcone et al., 1987 | p=0.1789 | **p<0.0001*** |
| **REML** | 8 | 50 vs 88 | 62.27 ± 24.85 vs 67.15 ± 19.65 | -0.894 (.2.461 to 0.672) | 0.263 | 41.256 | 93.60% | p<0.001 | 7 (p=0.014) | Reich et al., 1975.1 | Reich et al., 1975.1 | p=0.3988 | **p<0.0001*** |
| **REMD** | 3 | 8 vs 30 | 2.31 ± 0.20 vs 1.82 ± 0.10 | 3.12 (-0.388 to 6.635) | 0.081 | 21.365 | 89.27% | p<0.001 | 23 (p<0.001) | Benson et al., 1983 | - | p=1.0000 | p=0.0185 |
| **ST1** | 3 | 14 vs 41 | 43.48 ± 8.9 vs 37.74 ± 4.69 | 0.558 (-0.326 to 1.441) | 0.216 | 3.517 | 44.12% | p=0.172 | 1 (p=0.043) | - | - | p=0.3333 | p=0.0898 |
| **%ST1** | 3 | 14 vs 41 | 12.63 ± 5.6 vs 10.76 ± 4.70 | 0.203 (-0.411 to 0.817) | 0.516 | 0.161 | 0% | p=0.923 | 0 (p=0.289) | - | - | p=0.3333 | p=0.7012 |
| **ST2** | 4 | 23 vs 50 | 194.74 ± 18.30 vs 201.45 ± 11.41 | -0.137 (-1.836 to 1.561) | 0.874 | 26.218 | 88.52% | p<0.001 | 0 (p=0.250) | Zarcone et al., 1995 | - | p=0.7500 | p=0.5116 |
| **%ST2** | 3 | 14 vs 41 | 56.02 ± 6.20 vs 58.83 ± 6.30 | -0.198 (-0.907 to 0.510) | 0.583 | 2.372 | 19.39% | p=0.305 | 0 (p=0.327) | - | - | p=0.3333 | p=0.1563 |
| **ST4** | 3 | 26 vs 28 | 7.65 ± 16.17 vs 17.38 ± 26.08 | -0.461 (-1.007 to 0.084) | 0.097 | 0.862 | 0% | p=0.650 | 1 (p=0.049) | - | - | p=1.0000 | p=0.9210 |
| **%ST4** | 3 | 26 vs 28 | 1.88 ± 4.63 vs 4.68 ± 6.55 | -0.507 (-1.054 to 0.040) | 0.069 | 1.101 | 0% | p=0.577 | 1 (p=0.034) | - | - | p=1.0000 | p=0.7572 |
| **DELTAT** | 5 | 31 vs 62 | 38.33 ± 20.36 vs 42.39 ± 15.50 | -0.984 (-2.093 to 0.126) | 0.082 | 16.348 | 80.35% | p=0.003 | 16 (p<0.001) | - | - | **p=0.0167*** | **p=0.0001*** |
| **%DELTA** | 3 | 14 vs 41 | 9.91 ± 8.19 vs 10.25 ± 7.20 | -0.167 (-0.972 to 0.638) | 0.685 | 2.916 | 35.41% | p=0.233 | 0 (p=0.276) | - | - | p=0.3333 | p=0.0878 |

*Asymmetry found
